# Genome-wide association study of liver fat, iron, and extracellular fluid fraction in the UK Biobank

**DOI:** 10.1101/2021.10.25.21265127

**Authors:** Colm O’Dushlaine, Mary Germino, Niek Verweij, Jonas B. Nielsen, Ashish Yadav, Christian Benner, Joshua D. Backman, Nan Lin, GHS-RGC DiscovEHR Collaboration, Gonçalo R. Abecasis, Aris Baras, Manuel A. Ferreira, Luca A. Lotta, Johnathon R. Walls, Prodromos Parasoglou, Jonathan L. Marchini

## Abstract

Abdominal magnetic resonance imaging (MRI) represents a non-invasive approach allowing the extraction of clinically informative phenotypes. We developed an automated pipeline to segment liver pixels from abdominal MRI images and apply published models to approximate fat fraction, extracellular fluid fraction and iron content in 40,058 MRIs from the UK Biobank. We then conducted a genome-wide association of these traits using imputed variants (N=37,250 individuals, 11,914,698 variants) and exome sequence data (N=35,274 individuals, 8,287,315 variants). For liver fat we identified 8 novel loci in or near genes MARC1, GCKR, ADH1B, MTTP, TRIB1, GPAM, PNPLA2 and APOH. For liver iron we identified 1 novel locus between the genes ASNSD1 and SLC40A1, an iron transporter involved in hemochromatosis. For extracellular fluid fraction we identified 6 novel loci in or near genes AGMAT, NAT2, MRPL4-S1PR2, FADS1, ABO and HFE, with almost all having prior associations to obesity, liver, iron, or lipid traits.

## Introduction

Chronic liver disease is among the leading causes of morbidity and mortality, is often underdiagnosed and poses a substantial unmet clinical need^1^. Magnetic resonance imaging of the liver is able to capture liver fat and mark features of inflammation and fibrosis of the liver in a non-invasive manner and is therefore a powerful tool to study the genetic drivers of liver disease. The UK Biobank (UKB) is an ambitious research initiative aiming to characterize 500,000 individuals via extensive phenotyping together with genetic information^2^. A subset of 100,000 subjects are undergoing multiple MRI sessions of the abdomen and liver^3^, providing a rich resource to study genetics of well measured quantitative liver phenotypes, such as liver fat by proton density fat fraction (PDFF), hepatic iron content (HIC) and extracellular fluid fraction (ECF).

PDFF by MRI is considered a gold standard to quantify liver fat and has been demonstrated to be accurate when applied to MRI scans from the UKB^4^. Fatty liver is a key feature of chronic liver conditions such as non-alcoholic fatty liver disease (NAFLD) and the buildup of liver fat is an important precursor to steatohepatitis and liver fibrosis, which affects approximately 10% off middle-aged adults, and can lead to cirrhosis, hepatocellular carcinoma, and death. HIC, or hepatic iron content, marks iron concentrations in the liver. Excess iron, or iron overload, is associated with a range of liver conditions and metabolic disorders, including diabetes, high blood pressure, and cardiomyopathy^5^. Wilman and colleagues^6^ conducted a genome-wide association study (GWAS) of UKB MRI-derived liver iron among eight thousand individuals. They reported three genetic variants across HFE (2 independent variants) and TMPRSS6 that replicated in an independent dataset. ECF, or extracellular fraction, marks water accumulation and has previously shown to correlate with liver inflammation and fibrosis on histology^7^.

In this work, we present an automated workflow (**Figure 1** and **Methods**) to segment the liver from MRI images of 40,058 UKB participants and calculate PDFF, ECF and HIC by applying pre-defined mathematical models ^8-10^. We build on previous work on the genetics of liver MRI-derived traits by increasing sample size (over forty thousand samples) and analyze both rare exome and common imputed variants and report several novel associations.

**Figure 1.**
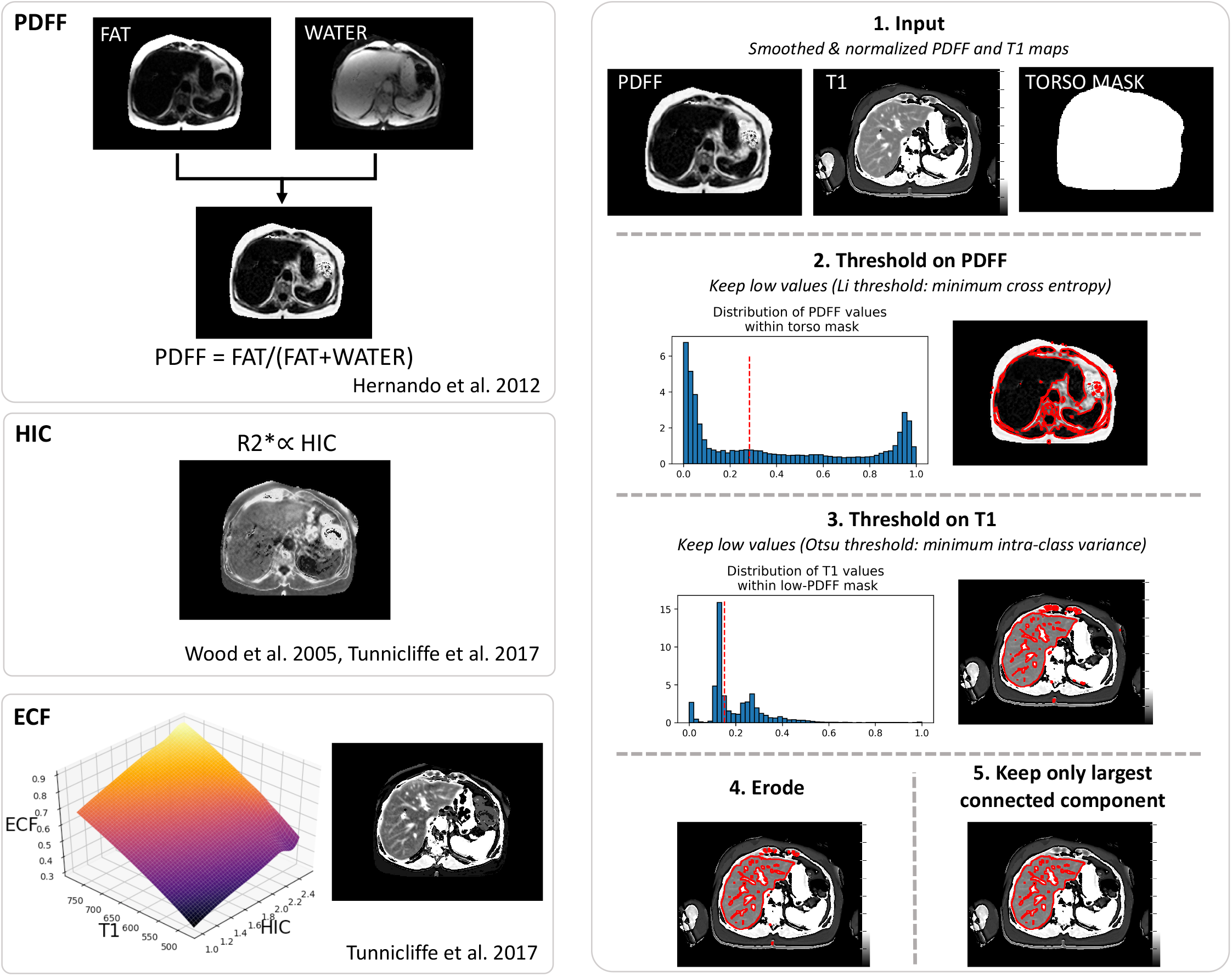
Summary of automated liver segmentation and derivation of liver image phenotypes. Three distinct phenotypes were derived from two abdominal MRIs acquisition, one for estimating fat content and the other a quantitative T1 mapping sequence: proton density fat fraction (PDFF), hepatic iron content (HIC) and extracellular fluid fraction (ECF, a proxy for liver fibrosis and inflammation). Pixels belonging to the liver were segmented using a thresholding approach, Li thresholding for PDFF maps to identify liver tissue, and Otsu thresholding for T1 maps to exclude larger vessels (see **Methods**). PDFF was estimated as the fraction of fat signal relative to total fat plus water signal. R2* was converted to HIC by a published linear model. ECF was estimated by interpolation from their published table containing grid points of a non-linear numerical model describing ECF as a function of T1 (from ShMOLLI MRI) and HIC (from IDEAL MRI), correcting for field strength.

## Results

### Imaging processing to extract liver MRI phenotypes

We developed an automated image processing pipeline to estimate liver fat, iron and extracellular fluid fraction by applying pre-defined mathematical models^8-10^ (**Figure 1** and **Methods**). We applied this pipeline to liver MRI images from 40,058 UKB participants. We compared our estimates of PDFF and cT1 phenotypes to those calculated by other groups and available directly from the UKB resource in much smaller subsets (<10,000 subjects) of participants (UKB data fields 22436, 22417). We would not expect perfect agreement due to differences in the processing pipelines. The Spearman rank correlation was 0.94 between our PDFF measure and that of “Liver_proton_density_fat_fraction (AMRA)” (UKB ID 22436). For cT1 (UKB ID 22417), the correlation was 0.88.

### Discovery analysis with imputed genetic data

We performed GWAS of PDFF, HIC and ECF using an imputed dataset of 11,914,698 variants and 37,250 individuals of European ancestry (see **Methods**). We ran two versions of the analysis the first adjusting for basic confounders (sex, age, age-squared, age*sex, top 20 principal components for ancestry, imaging center, imaging protocol) which we refer to as the baseline analysis, the second adjusting for additional confounders including body mass index (BMI), alcohol and other relevant comorbidity (BMI, BMI-squared, alcohol intake, weight loss/gain, diabetes, heart attack, angina, stroke, high blood pressure), which we refer to as the adjusted analysis. The inclusion of heritable covariates can bias effect estimates and increase false discovery rates if the variant being tested is associated with the covariate, but can also lead to an increase in power^11^.

Loci with statistically significant variants for each liver trait in the adjusted analysis are shown in **Figure 2**. The baseline analysis without adjustment for alcohol and disease confounders is summarized in **Supplementary Figures 1-3**. We used GCTA-COJO methodology^12^ to summarize the association results down to a set of approximately independent set of markers, and these are reported in **Table 1**. We used the software FINEMAP^13^ to refine these associated loci and estimate the most likely causal variants from the imputed data. Results of these analyses are shown in **Supplementary Figures 4-5**.

**Table 1.**
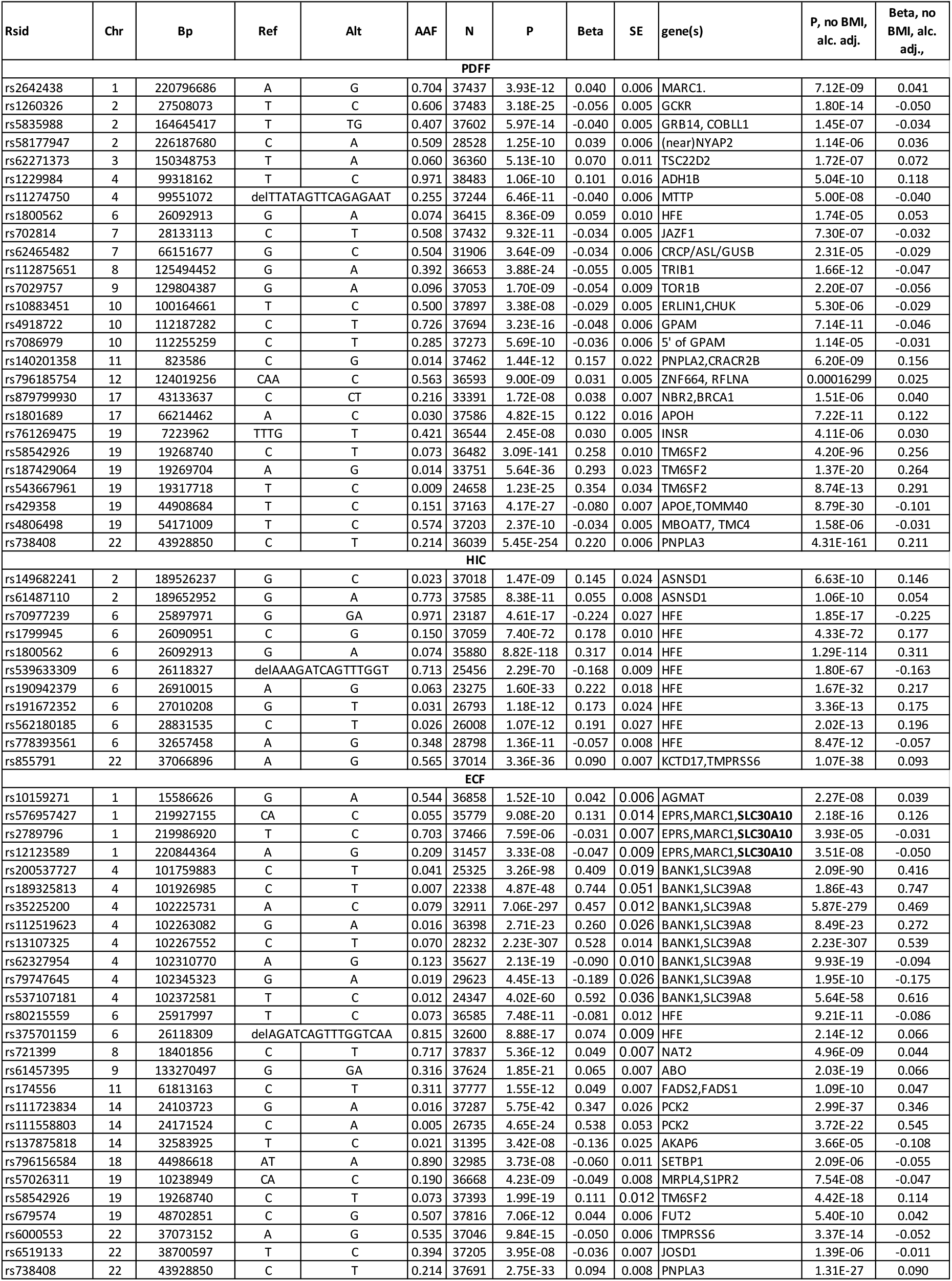
Sentinel SNPs from imputed GWAS. Independent genome-wide significant associations for PDFF, HIC, ECF. Results shown include additional covariate adjustment for BMI, alcohol, and additional covariates. Results for models without additional adjustment for BMI, alcohol, and additional covariates. Regional association plots are shown in **Supplementary Figures 5-7**. Association results at the associated loci for a carefully selected set of traits are shown in **Figure 4**.

**Figure 2.**
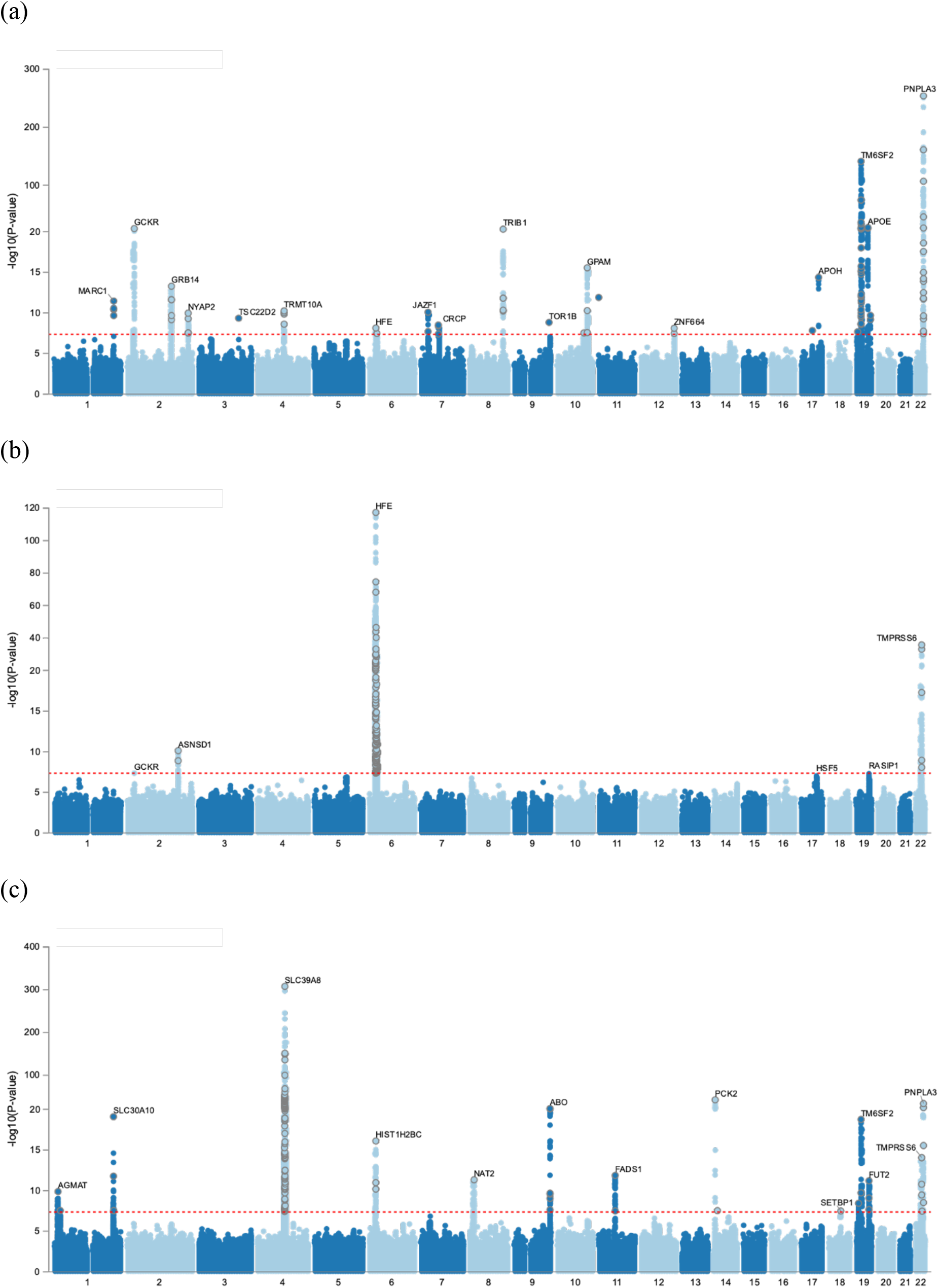
Manhattan plots for (a) PDFF, (b) HIC, (c) ECF. Results shown for GWAS of imputed data and include additional covariate adjustment for BMI and alcohol. Nearest genes are labeled.

For PDFF we identified 11 associated loci in the baseline analyses (**Table 1**). These included previously reported risk loci (PNPLA3, TM6SF2, APOE/TOMM40) for liver fat and related traits^14 15^, and 8 novel loci that highlight a central theme for lipid metabolism and in particular triglyceride generation and storage in regulating liver fat accumulation in humans. The SNP rs4918722 lies in the intronic region of GPAM, encoding an enzyme responsible for catalysis in phospholipid biosynthesis and is responsible for the first step in triglycerides synthesis. Rats overexpressing GPAM in the hepatocytes show steatosis and hepatic insulin resistance in absence of obesity or high fat diet^16^. On the other hand, GPAM knockout mice are protected against diet-induced steatosis by reducing triglyceride synthesis and storage^17^. The missense SNP rs1801689 is situated in APOH, which encodes for beta-2 glycoprotein that plays a role in various physiological processes including hemostasis and lipid metabolism such as triglyceride-rich lipoprotein clearance^18,19^. APOH is exclusively expressed in the liver. The intergenic SNP rs112875651 lies near TRIB1 (Tribbles-1), another gene involved in hepatic lipid metabolism and lipid homeostasis, the locus was first found to be associated with circulating lipid levels, primarily triglycerides levels^20^. The missense SNP rs140201358 is situated in PNPLA2, encoding for a key enzyme for intracellular hydrolysis of stored triglyceride in the liver (adipose triglyceride lipase, ATGL), and is closely related to PNPLA3. ATGL-deficient humans are presenting with lipid myopathy, in mice, generalized ATGL deficiency causes triglyceride deposition and progressive hepatic steatosis^21^.

The missense SNP rs2642438 in MARC1 was previously found to be protective for all cause cirrhosis^22^, decreased severity of NAFLD and hepatic lipid composition^23^. The missense SNP rs1229984 in the ADH1B gene, encoding alcohol dehydrogenase 1B, is a key enzyme in ethanol metabolism and reflecting alcohol-induced fatty liver. This SNP has recently been shown to modify the risk of NASH and fibrosis in adults with NAFLD regardless of alcohol consumption status^24^. The missense SNP rs1260326 in GCKR is well known to be associated with triglyceride levels^25^ and non-alcoholic fatty liver disease^26^. GCKR encodes for “glucokinase regulatory protein” which regulates glucokinase, a phosphorylating enzyme that modulates hepatic glucose metabolism and hepatic lipogenesis^27^. Our analysis identified common variant associations in and around the MTTP gene, encoding the microsomal triglyceride transfer protein, that are characterized by two causal sets in a fine-mapping analysis (**Supplementary Figure 4**).

Our analysis of PDFF that conditions on BMI, alcohol and disease variables also identified the loci GRB14-COBLL1, JAZF1, TOR1B, VKORC1L1-GUSB-ASL, NYAP2, TSC22D2, ZNF664-RFLNA, HFE (**Table 1**). Most of these loci (COBLL1, JAZF1, NYAP2, TSC22D2, ZNF664, ERLIN1, INSR (insulin receptor)) have lipid, BMI, T2D or waist-hip ratio associations in the GWAS catalog (see **URLs**). The SNP rs4806498 near MBOAT7 has been previously identified by other studies on liver fat^26,28-31^.

We note that rs62465482 in/near the gene ASL which encodes argininosuccinate lyase, and has been proposed as a superior biomarker to AST and ALT for the diagnosis of liver disease^32^. Fine mapping identified one credible interval of 629 variants spanning the interval chr7:65,719,502-67,229,875, with rs62465482 having the highest posterior probability of being causal. The nearby gene VKORC1L1 encodes an enzyme important in the vitamin K cycle. In a recent publication, Vkorc1l1 mouse knockouts displayed a considerably lower fat to body weight ratio, substantially decreased plasma leptin, and significantly underdeveloped white adipose tissue, suggesting that Vkorc1l1 promotes adipogenesis and possibly obesity and downregulation of Vkorc1l1 increases intracellular vitamin K2 level and impedes preadipocyte differentiation^33^. An additional locus implicated for PDFF is the gene HFE, a gene well-known to reflect iron levels.

Not adjusting for BMI or alcohol had a profound effect on the significance of many of the associated loci with (**Table 1, Supplementary Figure 2**), with most becoming no longer genome-wide significant. Several loci were not genome-wide significant in the base model but were in the model with additional covariates, by at least five orders of magnitude. These loci include variants in intervals spanning chr2:164645417-164811133 (COBLL1), chr2:27412596-27636484 (GCKR), chr8:125464631-125495147 (adjacent to TRIB1), chr10:112266288 (upstream of TECTB), chr19:18941011-20369092 (TM6SF2 and others), and chr22:43929868-44016312 (PNPLA3). The exception is APOE-TOMM40, with a p-value decreasing from 4.2×10^−27^ to 8.8×10^−30^ and effect estimate in standard deviation units going from -0.08 to -0.1 (not shown).

To examine the similarities and differences between SNPs associated with PDFF we carried out a clustering analysis of the association signals across a range of 52 traits. For each SNP-trait association we calculated the proportion of variance explained (PVE), either on the linear or liability scale. We rescaled the PVEs across the traits for each SNP using the maximum value, and then signed the PVEs according to direction of effect. We then applied bi-clustering to the resulting matrix of signed PVE estimates and visualized the result as heatmap in **Figure 3**. This figure highlights the widespread pleiotropy across the majority of PDFF associated variants. Notably, the majority of the PDFF associated loci do have a primary effect on PDFF, with some exceptions such as ADH1B on alcoholic liver disease, ERLIN1 on alanine transferase, APOE on c-reactive protein, TRIB1 and GCKR on triglyceride levels.

**Figure 3.**
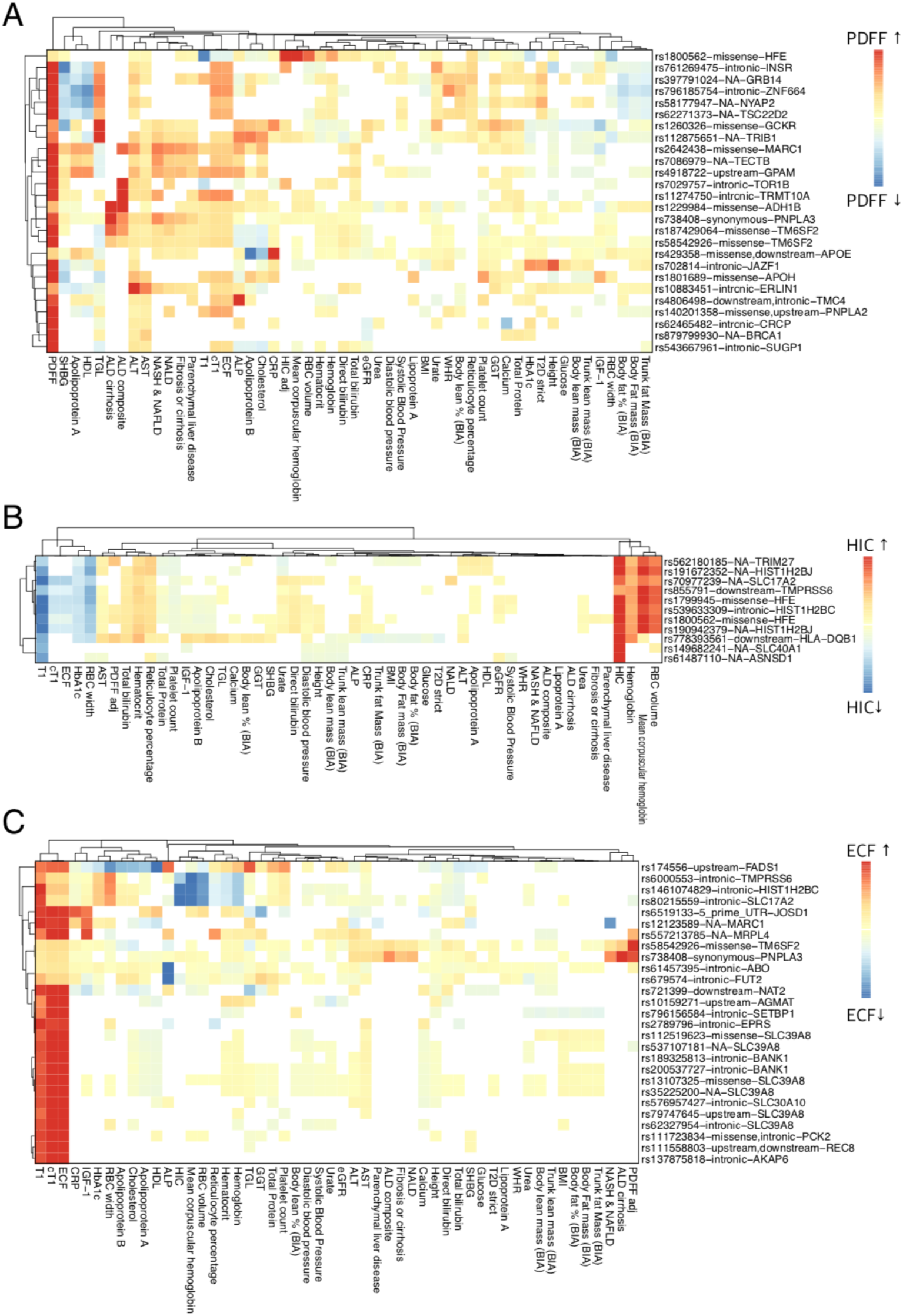
Phenome-wide association results for the associated loci. The heatmap shows the proportion of variance explained (PVE) across traits of interest by top sentinel variants from the analysis of (a) PDFF, (b) HIC and (c) ECF, signed by the direction of effect. The PVE values were normalized to have maximum at 1 and then signed depending on the direction of effect to compare association patterns between variants. Red colors indicate positive associations between the trait increasing allele and the other diseases or traits, blue colors indicate inverse associations and white colors indicate non-significant associations (P>0.005).

We also examined variants in our data that were previously reported to affect liver fat or risk of NAFLD (**Supplementary Table 1**). At the level of genome-wide significance (5×10^−8^), we replicate associations to GCKR^30,31^, NCAN^26^, MBOAT7-TMC4^28,29^ and ERLIN1^34^. Other known loci just below genome-wide significant associations but significant after multiple testing were PPP1R3B (P=2.7×10^−6^) and CHUK (P=1.3×10^−7^)^26,30^. CHUK is adjacent to ERLIN1 which is genome-wide significant in our data; fine-mapping of the ERLIN1 locus revealed most likely two credible intervals: chr7:100057584-100279430 encompassing ERLIN1 and CHUK (85 markers, mean absolute Pearson correlation among genetic variants in the credible set, computed from the same individual-level genotype that was used to generate the summary statistics (mean LD = 0.86), and chr10:99998189-100009635 encompassing DNMBP (852 markers, mean LD 0.08). Consistent with previous reports we do not see associations for HSD17B13, likely reflecting the role of the splice variant in affecting progression from steatosis to more severe liver pathology, but not in modulating fat fraction in the liver^35-38^.

For HIC, we identify 3 distinct genome-wide significant loci (**Figure 2**). The two associations in genes HFE and TMPRSS6 have been previously reported^6^. We identify one novel locus (rs149682241, between ASNSD1 and SLC40A1, **Supplementary Figures 5a-c**). The variant rs149682241 lies in the promoter region of SLC40A1, previously known as ferroportin, that has been implicated in hemochromatosis, Type 4^39^. The locus is a gene-dense region with multiple other potential causal candidate genes: a variant (rs6756571) in high LD (r^2^=0.96) with rs149682241 is a top eQTL for ORMDL1 in GTeX liver tissue (P=1.9×10^−18^), a gene approximately 150Kb from this locus. PMS1, a gene adjacent to ORMDL1 and 200Kb from the index variant was associated with ferritin levels in a GWAS of Chinese individuals^40^. Fine-mapping of this locus revealed two likely signals, chr2:189640971-189656328 (9 markers, mean LD 0.99, promoter region of ASNSD1) and chr2:189524434-189544472 (7 markers, mean LD 0.96, downstream of SLC40A1) (**Supplementary Figure 5c**). Neither ASNSD1 nor SLC40A1 have, to our knowledge, been linked by GWAS to iron or hepatic iron levels, though they have been associated with red blood cell, hematocrit and hemoglobin traits in the GWAS catalog (see **URLs**). In contrast to PDFF, not adjusting HIC for BMI, alcohol or additional ‘extra’ disease covariates generally improved significance of associated variants (**Table 1**). All loci had a primary effect on HIC and red blood cell related traits, except for the locus encompassing ASNSD1 and SLC40A1 showing more specificity for HIC and T1 alone (**Figure 3**). One marker (2:27517013:CT:C) at GCKR was significant at 5×10^−8^ but this association was not supported by other genetic variants (**Supplemental Figure 5d**).

**Figure 5.**
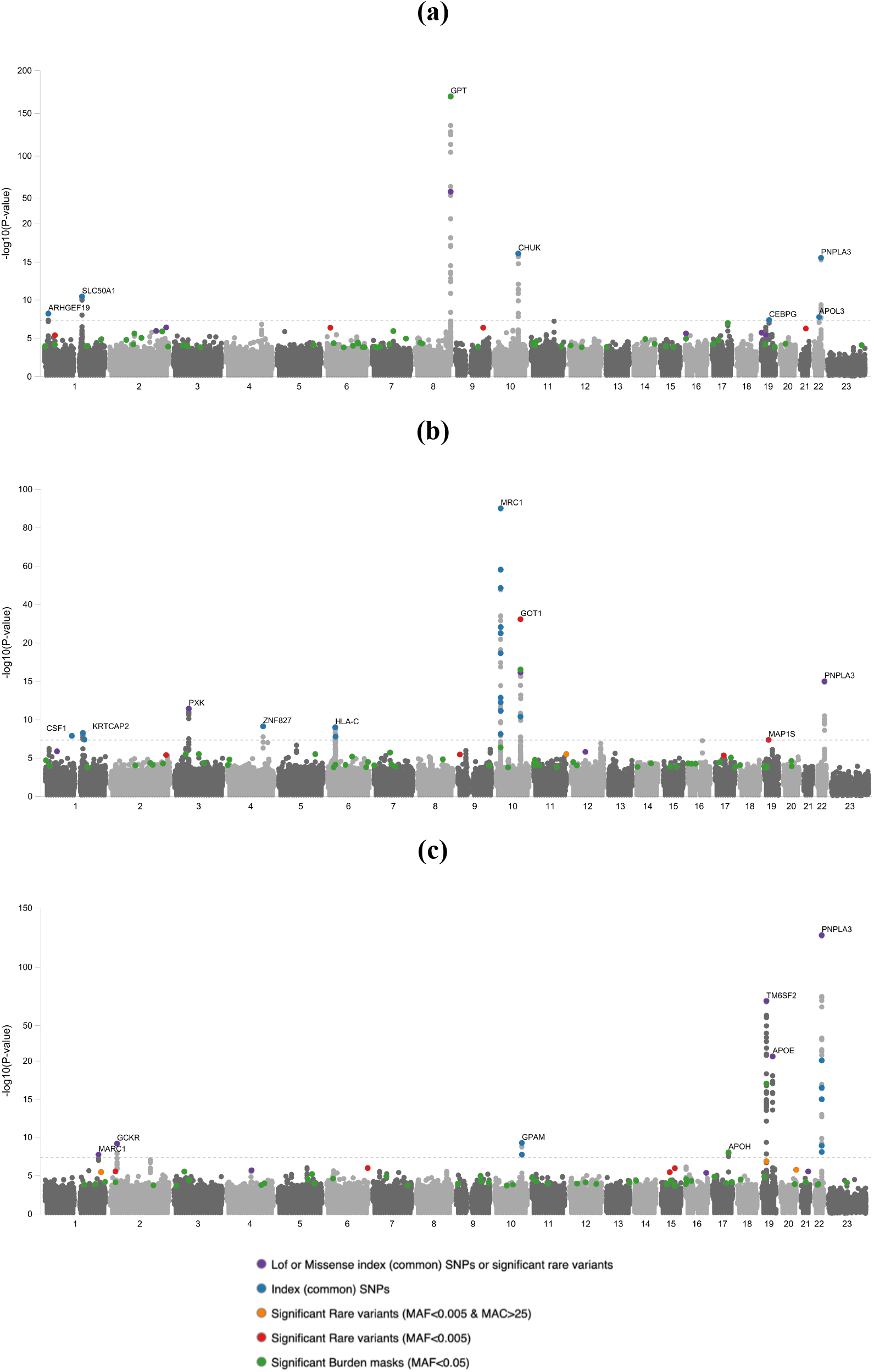
Comparisons of downsampled GWAS between (a) ALT, (b) AST, (c) PDFF.

For ECF, we identify 16 and 12 distinct loci in adjusted and baseline analyses (**Table 1, Figure 2, Supplementary Figure 6**). In our baseline analysis there are 6 novel loci in or near genes AGMAT, NAT2, MRPL4-S1PR2, FADS1, ABO and HFE. As we see for PDFF, most loci are more significant when adjusted for BMI or alcohol (**Table 1**). Almost all these loci have prior associations to obesity, liver, iron and/or lipid traits, including PNPLA3, TM6SF2, TMPRSS6, HFE, FUT2, NAT2, MRPL4-S1PR2, FADS1 and SETBP1, which are also observed in our phenome wide analyses (**Figure 3**). A locus that departs from this theme is AGMAT which has been linked to urate levels, glomerular filtration rate and alcoholic chronic pancreatitis (see GWAS catalog and URLs). Fine-mapping of this locus reveals one credible interval of 68 genetic variants (chr1:15487474-15597035, mean LD 0.96), encompassing DNAJC16 and AGMAT, top genetic variant being in the 5’ region of AGMAT.

The SNP rs721399 is near the gene NAT2 which is a phase II drug metabolizing enzyme responsible for detoxification of many commonly used hydrazine and arylamine drugs as well as common carcinogens ^41^. The observed associations in the HFE gene with ECF is expected since the encoded protein of HFE is a master regulator of iron metabolism and ECF is a measure corrected for liver iron^31^.

The genes PCK2, SLC39A8, TM6SF2, PNPLA3, TMPRSS6 and SLC30A10 were also identified by Parisinos and colleagues in a GWAS of cT1^31^. Fine mapping of the SLC30A10 association (**Supplementary Figure 6e**) reveals three credible intervals, two of which were at SLC30A10 and the third at MARC2-MARC1-HLX. SLC39A8 has been linked to schizophrenia, BMI, blood pressure cholesterol, blood manganese and, by extension, idiopathic scoliosis^42^ and loss-of-function mutations at the gene can cause undetectable serum manganese and disorders of glycosylation^43^. The top variant at this locus, rs13107325, has recently been implicated in the disruption of manganese homeostasis and intestinal barrier integrity^44^. Manganese is a cofactor for many enzymes, such as glycosyltransferases, and disruption to manganese transport by haploinsufficiency of SLC39A8 function might thus help cause a range of disorders of glycosylation. The liver has been implicated in approximately 22% of congenital disorders of glycosylation^45^, with symptoms ranging from anemia to fibrosis to hypoglycemia. These fall into liver specific and non-specific groups, see Marques-da-Silva and colleagues for details^45^.

The association at JOSD1 (rs6519133), which is only genome-wide significant in our adjusted analysis, was reported in a study of C-reactive protein (CRP)^46^. CRP is synthesized by the liver in response to inflammation, it is a general marker for inflammatory diseases and an independent predictor of coronary events^47^. The strong associations between JOSD1 (rs6519133), CRP, and ECF may point to an underlying molecular process involving inflammation or infection of the liver. Fine-mapping of the JOSD1 locus revealed one likely credible region: chr22:38525079-38745595, 240 variants, mean LD 0.9, spanning DMC1-SUN2, consistent with **Supplementary Figure 6o** and not offering a clear indication of any key gene(s) driving the signal.

### PDFF is a more powerful marker to detect genetic loci for liver fat than ALT or AST

Elevated levels of the enzymes alanine transaminase (ALT) and aspartate transaminase (AST) are indicative of (subclinical) liver damage caused by fat build up in the liver. To provide insights into the relationship between increased liver fat and elevated AST or ALT levels, we carried out GWAS experiments of AST and ALT in the same 32,726 individuals with PDFF measurements. Using a dataset of exome and array variants (PDFF values for 32,726 individuals, AST values for 31,411 individuals and ALT values for 31,499 individuals). **Figure 5** highlights top associations for each of these traits. We note 3-20x stronger associations at the loci PNPLA3, GPAM, MARC1(MTARC1), TM6SF2, APOE and GCKR (**Supplementary Table 3)**. For PNPLA3, the p-value was at least 7x stronger in PDFF compared to AST or ALT. Similarly, for TM6SF2 we see at least 10 orders of magnitude stronger p-values in PDFF. For more recently discovered candidates of interest in liver disease, GPAM and MTARC1(MARC1), we see at least 2 orders of magnitude stronger p-values for PDFF. APOE and GPAM signals appear to be specific to PDFF, and a number of signals appear to be specific for AST and ALT.

#### Polygenic risk scores predict liver disease traits in an independent dataset

We generated polygenic risk scores (PRS) from lead, COJO-independent and genome-wide significant associated genetic variants for PDFF (N=24 markers), ECF (N=23 markers) and HIC (N=7 markers) using the –score option in PLINK. We scored these variants in data from the Geisinger Health System (N=141,971 individuals), a merged dataset of exome and GWAS variants imputed into HRC with imputation quality 0.3 and above (22,258,434 total variants)). We then fitted a logistic model of the binary traits against the risk score, adjusting for on sex, age, age-squared and 4 principal components.

The PDFF PRS was associated with a range of non-alcoholic liver disease phenotypes (**Table 2**). The strongest associations occurred with the NAFLD, liver fibrosis and cirrhosis diagnoses. One standard deviation genetically determined higher PDFF was associated with 5.33 (95% CI 4.71-6.04) higher odds of NAFLD and steatohepatitis. There were also significant associations with liver cell carcinoma, Type 2 Diabetes and iron metabolism phenotypes. The ECF PRS had a similar, albeit less strong, pattern of association with the disease phenotypes. The HIC PRS exhibited the strongest association with ICD10 code E831 indicating individuals diagnosed with hemochromatosis (p-value = 7.8e-264, OR = 63.9[50.1-81.5]), other associations where with disorders of mineral metabolism and anemias.

**Table 2.** Polygenic scoring of select traits in Geisinger Health System (GHS) data. A genetic risk score was constructed from independent genome-wide significant genetic variants from PDFF, ECF and HIC GWAS and scores were derived in genetic data from GHS.

| Phenotype Description | Case N | Control N | PDFF |  |  |  | ECF |  |  |  | HIC |  |  |  |
| --- | --- | --- | --- | --- | --- | --- | --- | --- | --- | --- | --- | --- | --- | --- |
|  |  |  | Pvalue | OR | Conf2.5 | Conf97.5 | Pvalue | OR | Conf2.5 | Conf97.5 | Pvalue | OR | Conf2.5 | Conf97.5 |
| Non-Alcoholic Fatty Liver Disease and Steatohepatitis RGC Composite Definition | 5681 | 74303 | 1.53E-152 | 5.33 | 4.71 | 6.04 | 1.06E-11 | 1.42 | 1.28 | 1.57 | 0.6887 | 0.97 | 0.84 | 1.12 |
| ICD 9 : 571 - Chronic liver disease and cirrhosis | 10955 | 97683 | 9.53E-146 | 3.39 | 3.09 | 3.72 | 6.63E-14 | 1.33 | 1.24 | 1.44 | 0.4073 | 0.96 | 0.86 | 1.06 |
| Non-Alcoholic Liver Disease RGC Composite Definition | 9357 | 74303 | 9.42E-128 | 3.47 | 3.14 | 3.84 | 6.78E-12 | 1.33 | 1.23 | 1.45 | 0.4104 | 0.95 | 0.85 | 1.07 |
| ICD 10 : K758 - Other specified inflammatory liver diseases | 1435 | 117880 | 1.42E-98 | 11.77 | 9.35 | 14.79 | 2.30E-12 | 1.94 | 1.61 | 2.34 | 0.9794 | 1.00 | 0.76 | 1.33 |
| ICD 10 : K75 - Other inflammatory liver diseases | 2404 | 117880 | 1.09E-76 | 5.64 | 4.69 | 6.77 | 7.70E-09 | 1.55 | 1.33 | 1.80 | 0.4902 | 0.93 | 0.74 | 1.15 |
| Non-Alcoholic Liver Cirrhosis RGC Definition | 1166 | 74303 | 5.16E-72 | 10.49 | 8.11 | 13.55 | 0.0006 | 1.46 | 1.17 | 1.80 | 0.1704 | 1.24 | 0.91 | 1.68 |
| ICD 10 : K74 - Fibrosis and cirrhosis of liver | 1992 | 118231 | 5.78E-66 | 5.81 | 4.75 | 7.10 | 4.58E-05 | 1.41 | 1.19 | 1.66 | 0.1238 | 1.20 | 0.95 | 1.52 |
| ICD 10 : K746 - Other and unspecified cirrhosis of liver | 1740 | 118231 | 5.35E-62 | 6.16 | 4.97 | 7.63 | 0.0005 | 1.37 | 1.15 | 1.64 | 0.0508 | 1.28 | 1.00 | 1.65 |
| ICD 10 : E11 - Type 2 Diabetes RGC_T2D_new | 31178 | 96581 | 2.65E-27 | 1.43 | 1.34 | 1.52 | 5.62E-06 | 1.13 | 1.07 | 1.19 | 0.2270 | 0.96 | 0.89 | 1.03 |
| Alcoholic Liver Disease RGC Composite Definition | 623 | 74303 | 4.05E-26 | 6.78 | 4.75 | 9.65 | 0.1120 | 1.27 | 0.94 | 1.71 | 0.3483 | 0.81 | 0.52 | 1.25 |
| ICD 10 : K703 - Alcoholic cirrhosis of liver | 659 | 118508 | 1.43E-24 | 6.06 | 4.29 | 8.55 | 0.0556 | 1.33 | 0.99 | 1.76 | 0.6626 | 0.91 | 0.60 | 1.38 |
| Type 2 Diabetes by RGC EMR Phenotype Algorithm | 28155 | 68554 | 1.36E-23 | 1.44 | 1.34 | 1.55 | 6.19E-06 | 1.14 | 1.08 | 1.21 | 0.0200 | 0.91 | 0.84 | 0.98 |
| Type 2 Diabetes by RGC-modified eMERGE Network EMR Phenotype Algorithm | 22520 | 90681 | 1.18E-20 | 1.42 | 1.32 | 1.52 | 8.72E-05 | 1.13 | 1.06 | 1.19 | 0.4974 | 0.97 | 0.89 | 1.06 |
| ICD 10 : C220 - Liver cell carcinoma | 177 | 128883 | 1.56E-13 | 11.04 | 5.80 | 20.76 | 0.4410 | 1.25 | 0.70 | 2.15 | 0.0967 | 1.89 | 0.87 | 3.93 |
| ICD 10 : E831 - Disorders of iron metabolism | 972 | 89150 | 2.04E-12 | 2.85 | 2.13 | 3.81 | 6.66E-06 | 0.54 | 0.41 | 0.70 | 7.76E-246 | 63.89 | 50.07 | 81.47 |
| ICD 10 : E83 - Disorders of mineral metabolism | 7430 | 89150 | 2.19E-08 | 1.39 | 1.24 | 1.56 | 0.1009 | 0.92 | 0.84 | 1.02 | 1.89E-40 | 2.28 | 2.02 | 2.57 |
| ICD 10 : K740 - Hepatic fibrosis | 224 | 118231 | 0.0017 | 2.67 | 1.44 | 4.91 | 0.5269 | 1.18 | 0.70 | 1.92 | 0.8755 | 0.94 | 0.45 | 1.90 |
| ICD 10 : K743 - PBC | 92 | 118231 | 0.0058 | 3.74 | 1.44 | 9.43 | 0.0243 | 2.25 | 1.08 | 4.44 | 0.7426 | 0.83 | 0.25 | 2.47 |
| ICD 10 : D64 - Other anemias | 25495 | 92350 | 0.0370 | 1.08 | 1.00 | 1.15 | 0.4512 | 1.02 | 0.97 | 1.08 | 1.35E-32 | 0.61 | 0.56 | 0.66 |
| ICD 10 : D649 - Anemia, unspecified | 25098 | 92350 | 0.0404 | 1.08 | 1.00 | 1.15 | 0.4706 | 1.02 | 0.96 | 1.08 | 1.26E-32 | 0.61 | 0.56 | 0.66 |
| ICD 10 : K754 - Autoimmune hepatitis | 168 | 117880 | 0.1427 | 1.72 | 0.83 | 3.51 | 0.0605 | 1.69 | 0.96 | 2.89 | 0.6362 | 0.82 | 0.35 | 1.84 |
| ICD 10 : D509 - Iron deficiency anemia, unspecified | 11924 | 108801 | 0.1580 | 0.94 | 0.85 | 1.03 | 0.1040 | 1.06 | 0.99 | 1.14 | 1.95E-19 | 0.61 | 0.54 | 0.68 |
| Atopy RGC Composite Strict Definition | 15038 | 49105 | 0.1588 | 1.07 | 0.98 | 1.17 | 0.3989 | 1.03 | 0.96 | 1.11 | 0.0739 | 1.10 | 0.99 | 1.21 |
| Coronary Artery Disease by RGC EMR Phenotype Algorithm | 20054 | 104372 | 0.3725 | 1.04 | 0.96 | 1.12 | 0.7887 | 0.99 | 0.93 | 1.06 | 0.0013 | 0.86 | 0.79 | 0.94 |
| ICD 10 : B18 - Viral hepatitis | 2603 | 128217 | 0.3800 | 0.92 | 0.76 | 1.11 | 0.6600 | 0.97 | 0.83 | 1.13 | 0.3902 | 0.91 | 0.73 | 1.13 |
| ICD 10 : M62 - Other disorders of muscle | 13209 | 101995 | 0.3984 | 1.04 | 0.95 | 1.13 | 0.0310 | 1.08 | 1.01 | 1.16 | 0.7766 | 0.99 | 0.89 | 1.09 |
| ICD 10 : B19 - Viral hepatitis | 1481 | 128154 | 0.4008 | 0.90 | 0.70 | 1.15 | 0.8081 | 1.03 | 0.84 | 1.25 | 0.5827 | 0.92 | 0.69 | 1.22 |
| ICD 10 : D50 - Iron deficiency anemia | 14192 | 108801 | 0.5987 | 0.98 | 0.90 | 1.06 | 0.1367 | 1.05 | 0.98 | 1.13 | 1.63E-19 | 0.63 | 0.57 | 0.70 |

**Table 3.** Exome-wide significant rare variants in the exome dataset.

| Trait | Chr | Pos | Ref | Alt | rsID | P | Effect | MAF | MAC | variantEffect | variantEffectGene |
| --- | --- | --- | --- | --- | --- | --- | --- | --- | --- | --- | --- |
| ECF | 1 | 219928157 | G | A | rs188273166 | 5.06E-21 | 1.089 | 8.08E-04 | 57 | missense | SLC30A10 |
| ECF | 14 | 24096930 | C | G | rs61752842 | 1.60E-09 | 0.314 | 3.99E-03 | 280 | stop_gained | PCK2 |
| ECF | 14 | 24100214 | G | T | rs138881435 | 1.40E-10 | 0.428 | 2.44E-03 | 172 | splice_donor,intronic | PCK2,NRL |
| ECF | 14 | 24322263 | C | T | rs112742471 | 2.18E-08 | 0.296 | 3.80E-03 | 265 | downstream,intronic | LTB4R,ADCY4 |
| ECF | 16 | 71641052 | C | T | rs13337162 | 9.12E-08 | -1.131 | 2.41E-04 | 17 | downstream,3_prime_UTR | PHLPP2,MARVELD3 |
| ECF | 16 | 71669349 | A | G | rs11075896 | 6.54E-08 | -1.179 | 2.27E-04 | 16 | synonymous | PHLPP2 |
| ECF | 16 | 71748444 | T | C | rs28487278 | 6.54E-08 | -1.179 | 2.27E-04 | 16 | upstream | AP1G1 |
| ECF | 16 | 71753809 | T | C | rs9933587 | 6.54E-08 | -1.179 | 2.27E-04 | 16 | intronic | AP1G1 |
| ECF | 16 | 71756129 | T | C | rs34113755 | 6.54E-08 | -1.179 | 2.27E-04 | 16 | synonymous | AP1G1 |
| ECF | 16 | 71789541 | A | T | rs7191105 | 6.54E-08 | -1.179 | 2.27E-04 | 16 | intronic | AP1G1 |
| PDFF | 1 | 228224628 | A | G | rs202097101 | 3.15E-07 | 1.636 | 7.09E-05 | 5 | synonymous | OBSCN |
| PDFF | 2 | 21006019 | CA | C | rs982371659 | 4.24E-07 | 1.619 | 7.09E-05 | 5 | frameshift | APOB |
| PDFF | 6 | 166324496 | C | T | rs750742856 | 2.33E-07 | 1.51 | 8.50E-05 | 6 | intronic | SFT2D1 |
| PDFF | 19 | 19121300 | A | G | rs200744015 | 1.88E-07 | 0.204 | 4.82E-03 | 340 | splice_region | TMEM161A |
| PDFF | 19 | 19271164 | G | A | rs144821371 | 2.05E-10 | 0.296 | 3.40E-03 | 240 | intronic | TM6SF2 |
| PDFF | 19 | 19348800 | T | G | rs188840061 | 1.99E-10 | 0.282 | 3.77E-03 | 266 | intronic | MAU2 |

### Analysis of rare variants from exome data

For the exome data, we tested single variants and performed rare variant burden tests for all traits tested, using a significance threshold of P<=4.3×10^−7 48,49^ (**Supplementary Methods**). Exome-wide significant results for rare variants are shown in **Table 3**. Among these single markers, we identify 2 different loss-of-function mutations at PCK2 (rs61752842 and rs138881435, for ECF) and one at APOB (rs982371659, for PDFF). The enrichment of inactivating mutations at APOB associating to liver fat was also identified by a recent report^35^ and PCK2 was reported by Parisinos and colleagues in a GWAS of cT1^31^. APOB is reported to associate to several lipid traits in the GWAS (see **URLs**). We identify a missense mutation, rs188273166, at SLC30A10 for ECF (also reported by Parisinos and colleagues^31^) and a splice region association, rs200744015, at TMEM161A for PDFF. The gene TMEM161A, is located adjacent to TM6SF2 and unlikely to reflect an independent locus, supported by fine-mapping of common variants that revealed 2 credible intervals near TM6SF2: one variant - rs58542926 - at TM6SF2, and an interval of two variants chr19:19269704-19285807 in the 5’ region of TM6SF2 and overlapping SUGP1 (mean LD 0.99) (**Supplementary Figure 4b**). Similarly, MAU2 appears to be part of the TM6SF2 associated region. Finally, we note associations for variants at PHLPP2 and AP1G1 for ECF and OBSCN and SFT2D1 for PDFF. PHLPP2 has been implicated in BMI^50^ and AP1G1 has recently been implicated in HDL levels^51^. OBSCN does not appear to have an obvious connection to liver fat, though RNA-seq has implicated OBSCN among 1,185 genes with significant differences between subcutaneous and visceral fat^52^. A recent study of glucose-induced changes in gene expression in pancreatic islets, specifically gene expression in individuals with normal glucose tolerance versus individuals with hyperglycemia, highlighted increased expression of SFT2D1 between groups that was negatively correlated with insulin secretion^53^. Cross trait analyses using these variants are shown in **Supplemental Figure 7**.

Among gene-based burden tests, we see significant associations to PDFF for the genes APOH and APOB, also identified in the exome (APOB) and GWAS (APOH) single marker analysis. For ECF, we see associations for PCK2, SLC39A8, SLC30A10 and BDH2. SLC30A10 is a manganese transporter and has recently been implicated in liver health^54^. Autosomal mutations in SLC30A10 are linked to hypermanganesemia with dystonia, polycythemia, and cirrhosis (HMDPC)^55^ and mutant zebrafish models developed steatosis, liver fibrosis, and polycythemia accompanied by increased epo expression^56^. BDH2 is downregulated in hepatocellular carcinoma^57^ and has a role in iron homeostasis and affinity for ketone bodies^58-60^. However, it is likely the BDH2 signal is reflecting associations to ECF at the nearby gene SLC39A8 – fine-mapping revealed 6 credible intervals from chr4:99158072-105359633, four of which (4:102267552_C_T, rs112519623, rs79747645, chr4:102310770-102347606 (13 markers, mean LD 0.99) spanned SLC39A8, with two additional broad regions chr4:99163742-105342002 (888 markers, mean LD 0.07) and chr4:99318162-104767470 (132 markers, mean LD 0.58). Results for rare variant burden tests are shown in **Table 4**. Cross trait analyses using these genetic variants are shown in **Supplemental Figure 8**.

**Table 4.** Exome-wide significant rare variant burden tests in the exome dataset.

| ECF | Chr | Start | End | geneName | EnsemblGeneId | Mask | P | Effect | MAF | MAC |
| --- | --- | --- | --- | --- | --- | --- | --- | --- | --- | --- |
| ECF | 1 | 219915448 | 219928440 | SLC30A10 | ENSG00000196660 | M3.1 | 2.96E-11 | 0.671 | 0.00106 | 75 |
| ECF | 1 | 219915448 | 219928440 | SLC30A10 | ENSG00000196660 | M3.5 | 2.96E-11 | 0.671 | 0.00106 | 75 |
| ECF | 4 | 102253421 | 102344662 | SLC39A8 | ENSG00000138821 | M3.5 | 9.56E-27 | 0.283 | 0.016 | 1138 |
| ECF | 4 | 103079701 | 103096254 | BDH2 | ENSG00000164039 | M3.1 | 3.99E-08 | 0.19 | 0.00943 | 663 |
| ECF | 14 | 24094068 | 24103964 | PCK2 | ENSG00000100889 | M3.1 | 5.23E-25 | 0.287 | 0.014 | 1011 |
| ECF | 14 | 24094068 | 24103964 | PCK2 | ENSG00000100889 | M1.1 | 3.78E-22 | 0.325 | 0.00975 | 687 |
| PDFF | 2 | 21001729 | 21043945 | APOB | ENSG00000084674 | M1.001 | 7.43E-12 | 0.694 | 0.000709 | 50 |
| PDFF | 2 | 21001729 | 21043945 | APOB | ENSG00000084674 | M1.01 | 7.43E-12 | 0.694 | 0.000709 | 50 |
| PDFF | 2 | 21001729 | 21043945 | APOB | ENSG00000084674 | M1.1 | 7.43E-12 | 0.694 | 0.000709 | 50 |
| PDFF | 2 | 21001729 | 21043945 | APOB | ENSG00000084674 | M3.001 | 6.15E-09 | 0.291 | 0.00292 | 206 |
| PDFF | 17 | 66212132 | 66229379 | APOH | ENSG00000091583 | M3.5 | 3.51E-12 | 0.109 | 0.031 | 2128 |

For several loci of interest with rare or common variant associations (SLC30A10, PCK2, TMEM161A, APOB, BDH2), we compared ECF and PDFF images between random selections of carrier and non-carrier groups but did not observe clear visual differences between them (data not shown).

## Discussion

To gain more insights into the genetics of liver fat, iron and inflammation, we characterized liver MRI images from the UKB and conducted genetic association studies. We extracted biologically meaningful, quantitative traits – fat fraction, fluid fraction and iron content – from thousands of liver MRI images. Genome- and exome-wide analyses was performed on these traits, confirming previously published associations and identify several new ones that provide insights into genetic factors underlying fat, iron content and inflammation in the liver. These analyses identified 11 genetic loci for liver fat by PDFF, 3 genetic loci for iron overload by HIC and 16 genetic loci for liver inflammation by ECF. These results permit several conclusions.

First, through genetic associations we confirm previously hypothesized biological mechanisms that contribute to liver diseases. For liver fat we identified 8 novel loci in or near genes containing MARC1, GCKR, ADH1B, MTTP, TRIB1, GPAM, PNPLA2 and APOH, that highlight a central theme for lipid metabolism and in particular triglyceride generation and storage in regulating liver fat accumulation in humans. For example, the MTTP locus where loss of function MTTP mutations cause autosomal recessive forms of abetalipoproteinemia (MIM: 200100), while loss of function APOB mutations cause co-dominantly inherited forms of familial hypobetalipoproteinemia type 1^61^. Genetic loss of function of these genes or the pharmacological inhibition of their gene products ^62^ results in the inability to assemble and secrete liver-synthesized apolipoprotein B-containing lipid particles, resulting in liver fat accumulation and damage. A separate mechanism contributing to liver fat accumulation was illustrated by loci containing genes that are involved in fat distribution and insulin resistance due to implicating impaired peripheral adipose storage^63-67^. These include the TRIB1, GRB14/COBLL1, PNPLA2 and INSR loci. These associations suggest that individuals who carry alleles associated with an impaired ability to store fat in peripheral adipose compartments, develop more substantial ectopic fat deposition in the liver ^68^. Furthermore, by studying the ECF phenotype we provide insights into factors leading to liver inflammation such as excess liver fat, as illustrated by the PNPLA3 locus and metal accumulation, as illustrated by the HFE locus.

Second, precisely measuring the genetic analyses of quantitative liver imaging traits improves our understanding of the common genetic basis of liver disease. The strength of our study lies in the precise measurements of liver MRI data. Our approach was superior to previous efforts. For example Parisinos and colleagues ^31^ conducted a GWAS on cT1, a method to grade the severity of steatohepatitis and liver fibrosis, across only fifteen thousand individuals, reporting six independent genome-wide significant associations – four at SLC30A10 and one at TM6SF2 and PNPLA3. Haas and colleagues^35^ performed a GWAS of liver fat in UKB by training a deep learning model on publicly available liver fat estimates for 4,511 UKB individuals, produced by Perspectum Diagnostics^4^, and estimating fat fraction in remaining individuals with imaging data. Some of the effect sizes that we observed for liver fat loci were larger compared to Haas et al, suggesting our approach more precisely measured liver fat from the MRI (**Supplementary Table 1)**. To this notion, liver-fat related loci such as those implicating *PNPLA3* and *GPAM*, analyses of liver imaging phenotypes (particularly PDFF) had several orders of magnitude stronger statistical evidence of association than analyses of proxy traits such as liver transaminases. These observations, together with the finding of an association between liver imaging PRSs and liver disease outcomes, and the novel associations reported in this study suggest that the expansion of genetic data on imaging derived liver phenotypes will be a valuable tool to better understand the causes of liver disease. To date, only ∼40% of the planned UKB liver MRIs have been released. As this sample size increases to 100,000 extraction of liver phenotypes will continue to shed new light on the genetic factors underlying the pathophysiology of liver disease.

Third, through exome sequencing and rare variant association analyses, we confirm candidate genes that were identified in the common variant analyses such as SLC30A10 and PCSK2, illustrating the strength of complementing genome-wide analysis of common variants with exome-wide rare variant analyses.

Altogether, by applying new sophisticated machine learning methods to analyze liver MRI and combining it with genetic analyses, we were able identify biological insights into liver fat, hepatic iron accumulation and liver inflammatory mechanisms. These data provide new opportunities to study their role in disease and drug development.

## Data Availability

Individual-level imaging, genetic and phenotypic data are available to approved researchers of UK Biobank.

## Acknowledgements

This research has been conducted using the UK Biobank Resource (Project 26041).

## Ethics Statement

Ethical approval for the UK Biobank was previously obtained from the North West Centre for Research Ethics Committee (11/NW/0382). The work described herein was approved by UK Biobank under application number 26041. Approval for DiscovEHR analyses was provided by the Geisinger Health System Institutional Review Board under project number 2006-0258. Informed consent was obtained for all study participants.

### Online Methods

Hepatic iron content (HIC) can be derived from MRI relaxation time techniques. Specifically, iron shortens T1, T2 and T2* relaxation times measured by MRI, darkening images when iron is present^9^. T1 relaxation time also reflects extracellular fluid fraction and is related to fibrosis and inflammation^31^. Banerjee et al. 2014 ^7^ have previously reported how cT1 (T1 measurements corrected for iron) correlates positively with hepatic fibrosis. To better calculate ECF, a proxy for fibrosis, Tunnicliffe and coworkers^10^ developed a sophisticated model of the liver, accounting for blood, interstitium, two intracellular spaces, semisolid and liquid using volumes and other factors to describe ECF as a function of T1 and HIC.

### MRI sequences

Most UKB participants selected for liver MRI underwent two acquisitions, one for estimating fat content and the other a quantitative T1 mapping sequence. For the former, approximately 10,000 subjects were imaged under a Dixon gradient echo protocol; in 2016, the acquisition protocol for measurement of fat fraction was updated to the IDEAL sequence (Iterative Decomposition of water and fat with Echo Asymmetry and Least-squares estimation). Data from this acquisition are provided as a series of complex-valued 2D images per subject. The in-plane pixel size is 2.5×2.5 mm; slice thickness is 6 mm. The latter protocol, “ShMOLLI” (Shortened Modified Look-Locker Inversion recovery), has been consistent throughout the study. Data for this acquisition are provided as one real-valued 2D pre-computed T1 map per subject. The in-plane pixel size is 1.15×1.15 mm; slice thickness is 8 mm. Both MRI datasets were acquired at the same 2D cross-section per subject, intended to be through the porta hepatis. All images were acquired on a Siemens MAGNETOM Aera 1.5T clinical MRI scanner.

### Parameter Estimation

Parametric maps (pixel-wise parameter estimates) were generated for each trait, per subject, from images obtained from UKB. Signal magnitudes of fat and water, and relaxation rate 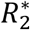 were estimated from the earlier Dixon protocol via the 3-point Dixon method^69^ using the 2^nd^, 3^rd^, and 4^th^ echoes, as done by Mojathed et al.^70^. Here, we briefly recapitulate the exposition of the Dixon 3-pt technique published by Ma ^71^. Let *S* be the complex value of pixel at co-ordinates (*x,y*) in a gradient echo image, *W* and *F* be the water and fat signal amplitudes respectively, then the general model is given by:

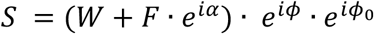

where *α* is the phase angle between fat and water signals, *ϕ* is the error phase due to magnetic field inhomogeneity, and *ϕ*_0_ is error phase due to system imperfections. Note that the parameters *S,W, F* ,*ϕ* ,*ϕ*0 are dependent on the co-ordinates (*x,y*). The phase angle *α* is a user defined parameter as part of the imaging protocol. The signal intensities *S*_0_, *S*_1_, and *S*_2_ at each pixel for echoes acquired, respectively, at 0°,180°, and 360° phase shifts (comprising a Dixon 3-point acquisition) can thus be written:

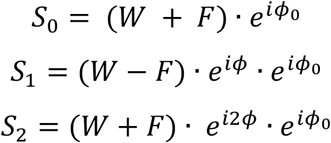

*ϕ* can be estimated as:

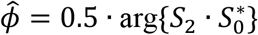

From these, the following expressions for water (W) and fat (F) amplitudes in each pixel can be derived and used for estimation:

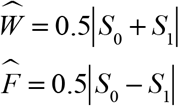

An estimate for 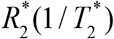, which is needed to compute HIC, can be obtained by fitting a decaying exponential to the magnitudes of the in-phase echoes using curve_fit from the scipy.optimize Python package.

The IDEAL sequence images were processed using the mixed magnitude/complex fitting method of Hernando et al. 2012, which is based on iterative least squares^8^. In this approach the signal model for image *s*_*n*_at echo *n*is given by:

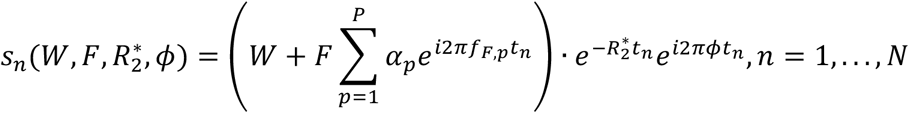

where:

- *W*and *F*are water and fat signal amplitudes, respectively
- 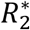 is the 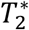 decay rate
- *ϕ*is phase error due to magnetic field inhomogeneity
- The fat signal is assumed to be comprised of *P* =6spectral peaks at frequencies *f*_*F*_ = [−249.093, −223.545, −172.449, −130.2948, −31.2963, 31.935] Hz with relative amplitudes *α*= [0.087, 0.693, 0.128, 0.004, 0.039, 0.048]
- *N* =6echos were acquired at echo times *t* = [1.2, 3.2, 5.2, 7.2, 9.2, 11.2]

Hernando et al.^8^ give the following expression for “mixed” (combined magnitude/complex) estimation of the desired parameters from measured signal *s*_(,3456_ for each of *N* echos:

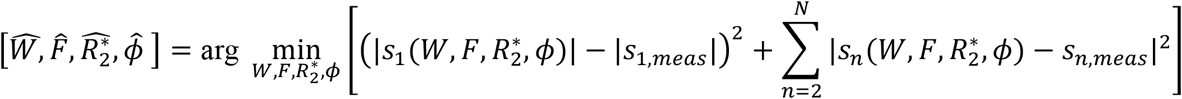

These estimates 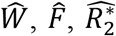 and 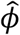 can be obtained via non-linear least squares fitting (e.g., as implemented in Python’s scipy.optimize package).

PDFF was estimated as the fraction of fat signal relative to total fat plus water signal.

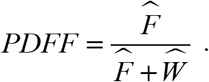

R2* was converted to HIC by a published linear model^9^

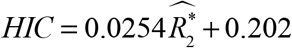

The implementation was validated using a publicly available phantom dataset containing vials of varying concentrations of fat^72^.

Tunnicliffe and colleagues^10^, developed a multi-compartment model of the liver to simulate the effects of presence of iron and fibrosis on shortened-MOLLI (ShMOLLI) T1 measurements. This model consists of the blood, interstitium, and two intracellular spaces, semisolid and liquid using volumes, relaxation rates and exchange rates previously reported in the literature. We used interpolation applied to the published results (Table 2 in Tunnicliffe and colleagues^10^) to estimate ECF from the T1 and HIC values at each pixel, correcting for field strength (**Figure 1**). ECF is used as a proxy to fibrosis and inflammation.

### Automated Liver Segmentation

Pixels belonging to the liver were automatically identified using a multi-thresholding approach across the PDFF and T1 parametric maps for each subject. After Gaussian smoothing, low pixel values in the PDFF map were identified by Li thresholding; these pixels comprise the liver, as well as other relatively low-fat regions such as the spleen. The corresponding subset of pixels in the T1 map were then subjected to Otsu thresholding, with the lower-intensity pixels retained in the liver region of interest. This step effectively excludes larger vessels and some other non-liver regions. Further refinement of the region was accomplished by morphological erosion, and finally, removal of all but the largest connected component in the resulting segmentation. To obtain a summary measure of each trait per subject, all pixels within the liver were averaged for each parametric map (**Figure 1**).

### Image processing quality control

Quality issues encountered in this dataset included mis-positioning of plane of imaging (such that little to no liver was included in the field of view); poor model fits resulting from signal loss, magnetic field inhomogeneities and/or other phase errors (especially in larger subjects); and fat/water swapping (convergence to conjugate solution because of phase wrapping) (**Supplementary Figure 9**).

A “quality control region of interest (QC ROI)” was defined as a circular region of fixed diameter, positioned according to bounding box and centroid of torso mask, based on expected positioning of liver within field-of-view. From this ROI, two metrics were used to filter images for quality (**Supplementary Figure 10**). We removed images with poor model fit and/or ROI placement, 4.4% of images (**Supplementary Figure 10**). We removed second scans for individuals with multiple scans, leaving 40,058 subjects/images. Demographic characteristics for this set of individuals, compared to the rest of participants are shown in **Supplementary Table 4**.

### Image processing computational resources

All image processing was performed with in-house Python implementations and standard libraries (scipy, numpy, scikit-image, etc.) in a parallelized computing environment. Initial work was done using an on-premises high-performance computing cluster, and later work was carried out on a cloud-based high-performance computing cluster. Computation time was approximately 2 minutes per subject.

### Relationships across derived phenotypes

We observed modest correlations (for traits deconfounded with ‘extra’ covariates, see section on trait deconfounding and genetic analysis) between PDFF and ECF (Spearman rank correlation=0.35) and between PDFF and HIC (Spearman correlation=0.34). The correlation between ECF and HIC was weaker at 0.02 (**Supplementary Figure 11**).

### UK Biobank data

A detailed description of the UKB study design has been published previously^2^ and consists of over 500,000 individuals between the ages 40-69^73^. A subset of individuals underwent detailed imaging across multiple modalities, including abdominal MRI, between years 2014 and 2019^3^. Raw liver imaging data was downloaded from UKB data fields 20203, 20204, 20254. Array and imputed genetics data was downloaded from UKB data fields 22418 and 22828 respectively. Sample preparation, exome sequencing, QC and genotype calling were done at the Regeneron Genetics Center as previously described^74 75^.

### Trait deconfounding and genetic analysis

All traits were deconfounded by residualizing the traits with the following covariates: sex, age, age-squared, top 20 principal components for ancestry, age*sex, imaging center, imaging protocol. Additional covariates (referred to as ‘extra’ here), were BMI, BMI^2^, 7 binary alcohol variables (daily, 1-2 times per week, 3-4 times per week, 1-3 times per month, special occasions, previous, current), 2 binary weight gain variables (weight gain in last year, weight loss in last year) and 5 binary disease variables (diabetes, heart attack, angina, stroke, high blood pressure). **Supplementary Figure 12** shows, for the most significant covariates, the distribution of significance of covariate effects across the three traits, in addition to the pairwise correlations between all traits and covariates. GWAS and ExWAS were performed using a linear mixed model in REGENIE^76^ (see **URLs**). We included in step 1 of REGENIE (prediction of trait based on genetic data) 211,683 variants that were directly genotyped, had a minor allele frequency (MAF) >5%, <1% genotype missingness, Hardy-Weinberg equilibrium test P-value>10^−15^, and after linkage-disequilibrium (LD) pruning (r^2^<0.5). Our analysis was applied to the European subset of the data, defined as individuals predicted to be European by applying a linear model trained based on PC estimates from HapMap3 and projecting these onto our data, as described previously^77^. We performed genome-wide association scans (GWAS) on each of our liver traits (PDFF, HIC, ECF), testing against imputed array data (N=37,250 individuals, 11,914,698 variants) and exome sequence data (N=35,274 individuals, 8,287,315 variants). Imputed UKB variants were filtered based on minor allele frequency (MAF≥0.5%) and Hardy-Weinberg (P<=10^−15^).

### Rare variant burden tests

Rare variant burden tests were carried out as previously described{Van Hout, 2019 #44; {Backman, 2021 #97}}. For each gene region defined (Ensembl 2, GRCh38), genotype information from multiple rare coding variants was collapsed into a single burden genotype, such that individuals who were: (i) homozygous reference (Ref) for all variants in that gene were considered homozygous (RefRef); (ii) heterozygous for at least one variant in that gene were considered heterozygous (RefAlt); (iii) and only individuals that carried two copies of the alternative allele (Alt) of the same variant were considered homozygous for the alternative allele (AltAlt). We did not phase rare variants, and so compound heterozygotes, if present, were considered heterozygous (RefAlt). We did this separately across four classes of variants 3: (i) predicted loss of function (pLoF), which we refer to as an “M1” burden mask; (ii) pLoF or missense (“M2”); (iii) pLoF or missense variants predicted to be deleterious by 5/5 prediction algorithms (“M3”); (iv) pLoF or missense variants predicted to be deleterious by 1/5 prediction algorithms (“M4”). The five missense deleterious algorithms used were SIFT ^78^, PolyPhen2 (HDIV), PolyPhen2 (HVAR)^79^, LRT^80^, and MutationTaster^81^. For each gene, and for each of these four groups, we considered five separate burden masks, based on the frequency of the alternative allele of the variants that were screened in that group: <5%, <1%,, <0.1%, <0.01%, <0.001% and singletons only. In main text and tables we use a shorthand notation for each mask, for example, M1.01 denotes the “M1” burden mask with the <0.1% allele frequency bin. Each burden mask was then tested for association with the same approach used for individual variants. In presenting results, for single variants we used a maximum minor allele frequency of 0.005 and minimum minor allele count of 5 to define our rare variant set. For burden tests, we allowed a maximum minor allele frequency of up to 0.05 and minimum minor allele count of down to 1.

### Conditional Analysis of Identified Loci

We identified all independent signals reaching genome-wide significance in our study with an approximate conditional and joint analysis (COJO) implemented in GCTA^12^. We used a subset of 10,000 unrelated UKB participants as a reference population.

FINEMAP^13^ implements a statistical algorithm for fine-mapping causal variants in genomic regions associated with complex traits and diseases. FINEMAP is computationally efficient by using summary statistics from genome-wide association studies and robust by applying a shotgun stochastic search algorithm. We ran FINEMAP under default settings with the option to allow for 30 causal variants.

### URLs

GWAS catalog: https://www.ebi.ac.uk/gwas/

REGENIE: https://github.com/rgcgithub/regenie

## Notes

### Competing Interest Statement

All authors own stock or stock options in Regeneron Pharmaceuticals

### Funding Statement

UK Biobank Exome Sequencing Consortium (AbbVie, Alnylam Pharmaceuticals, AstraZeneca, Biogen, Bristol-Myers Squibb, Pfizer, Regeneron and Takeda) for generation the whole exome sequencing data. All authors are employees of Regeneron Pharmaceuticals.

