## Supplementary Material for "Genome-wide association study of liver fat, iron, and extracellular fluid fraction in the UK Biobank"

#### List of investigators from the DiscovEHR cohort

All authors are listed in alphabetical order.

Lance J. Adams[1], Jackie Blank[1], Dale Bodian[1], Derek Boris[1], Adam Buchanan[1], David J. Carey[1], Ryan D. Colonie[1], F. Daniel Davis[1], Dustin N. Hartzel[1], Melissa Kelly[1], H. Lester Kirchner[1], Joseph B. Leader[1], David H. Ledbetter, Ph.D.[1], J. Neil Manus[1], Christa L. Martin[1], Raghu P. Metpally[1], Michelle Meyer[1], Tooraj Mirshahi[1], Matthew Oetjens[1], Thomas Nate Person[1], Christopher Still[1], Natasha Strande[1], Amy Sturm[1], Jen Wagner[1], Marc Williams[1]

Affiliations:
1. Geisinger, Danville, PA, USA

### Supplementary Figures

(a)

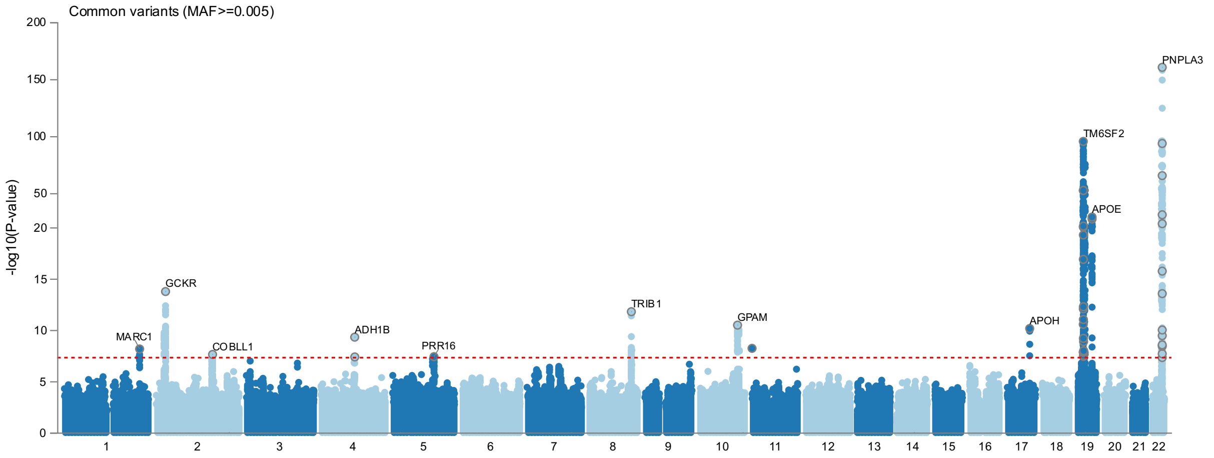

(b)

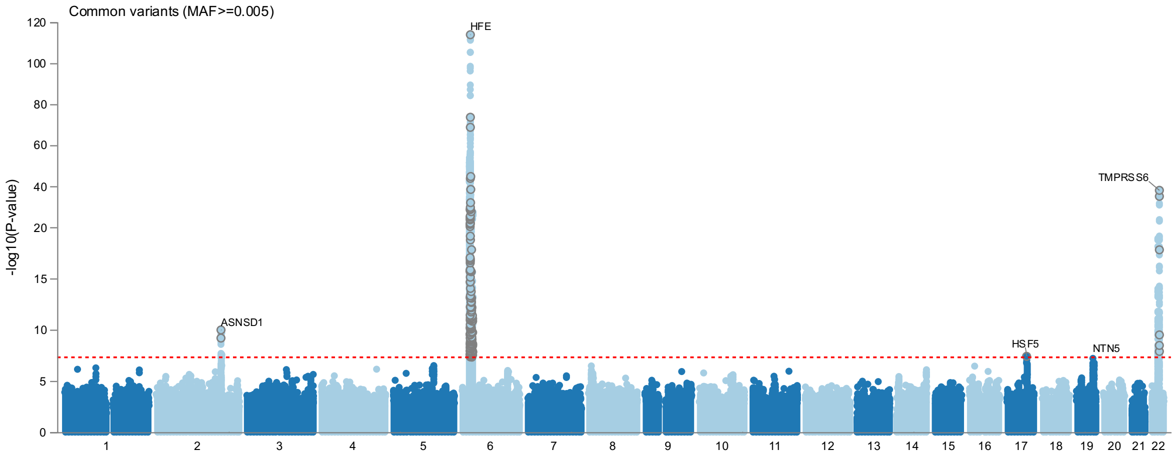

(c)

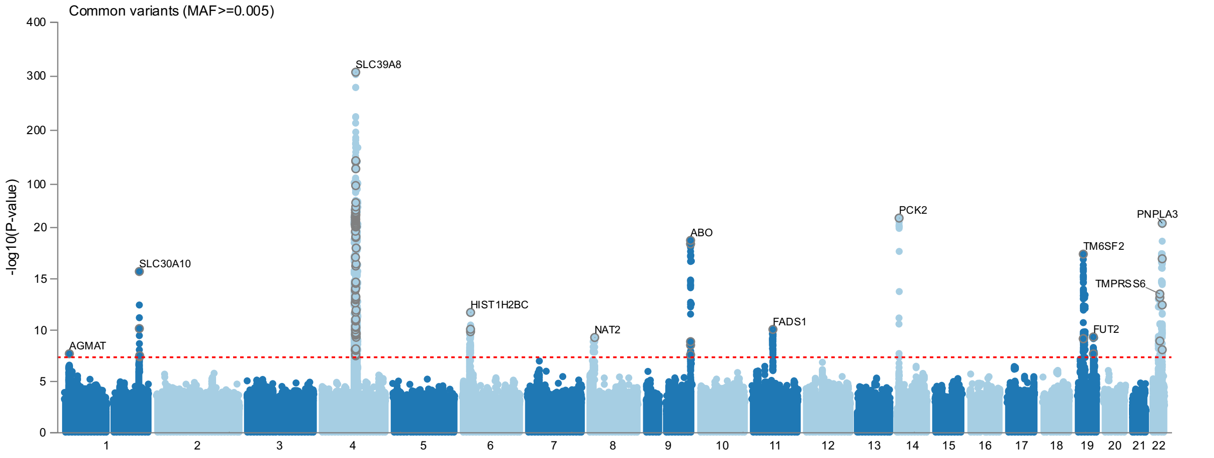

#### Supplementary Figure 1. Manhattan plots without BMI and alcohol covariate adjustment.

(a) PDFF, (b) HIC, (c) ECF. Results shown are for GWAS of imputed data.

(a)

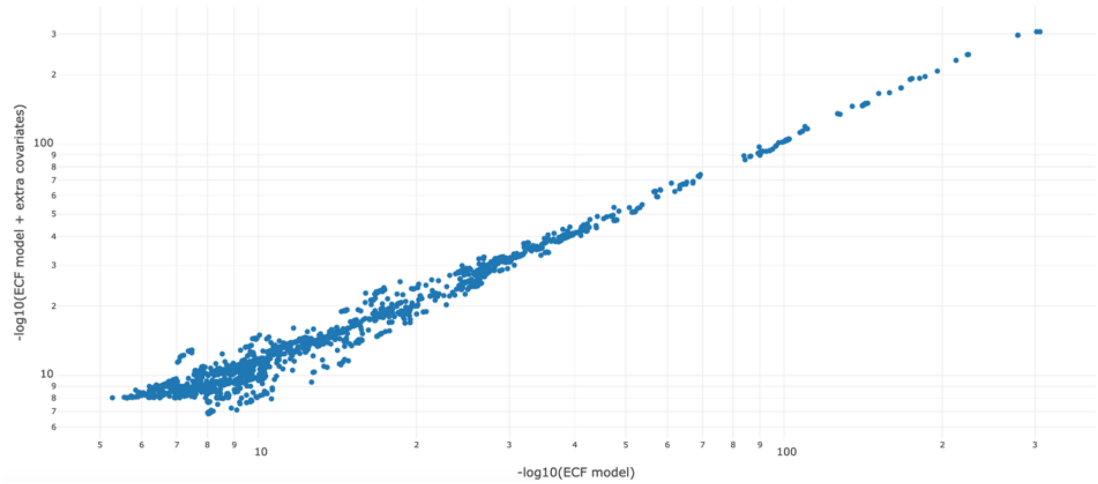

(b)

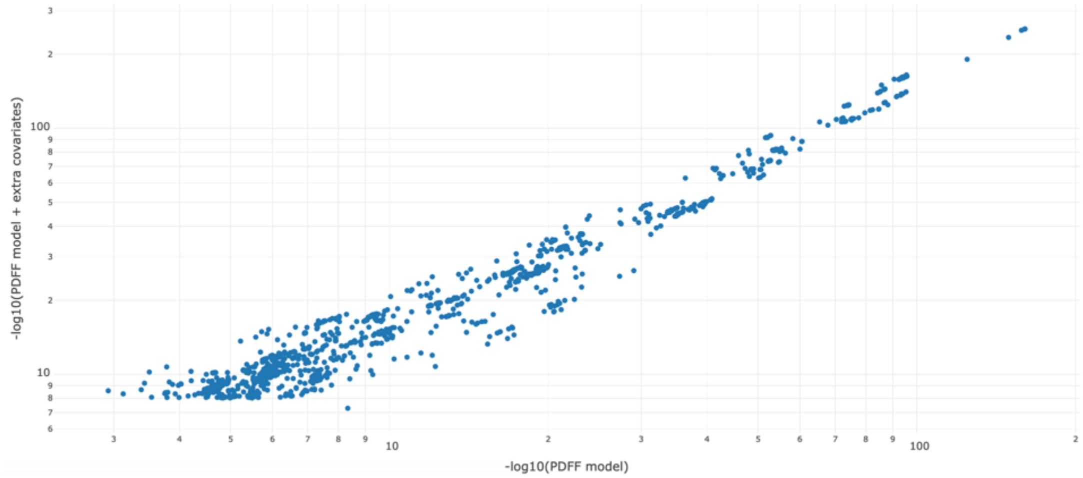

(c)

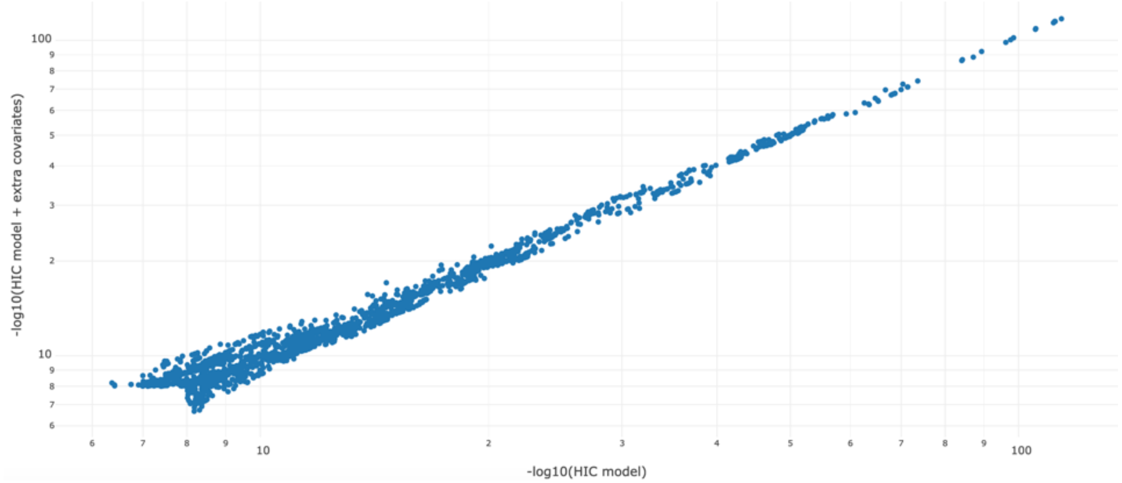

Supplementary Figure 2. Comparisons of models with and without additional covariate adjustment.
(a) ECF, (b) PDFF, (c) HIC. Results shown for GWAS of imputed data for variants with P<1e$^{-8}$ for either model.

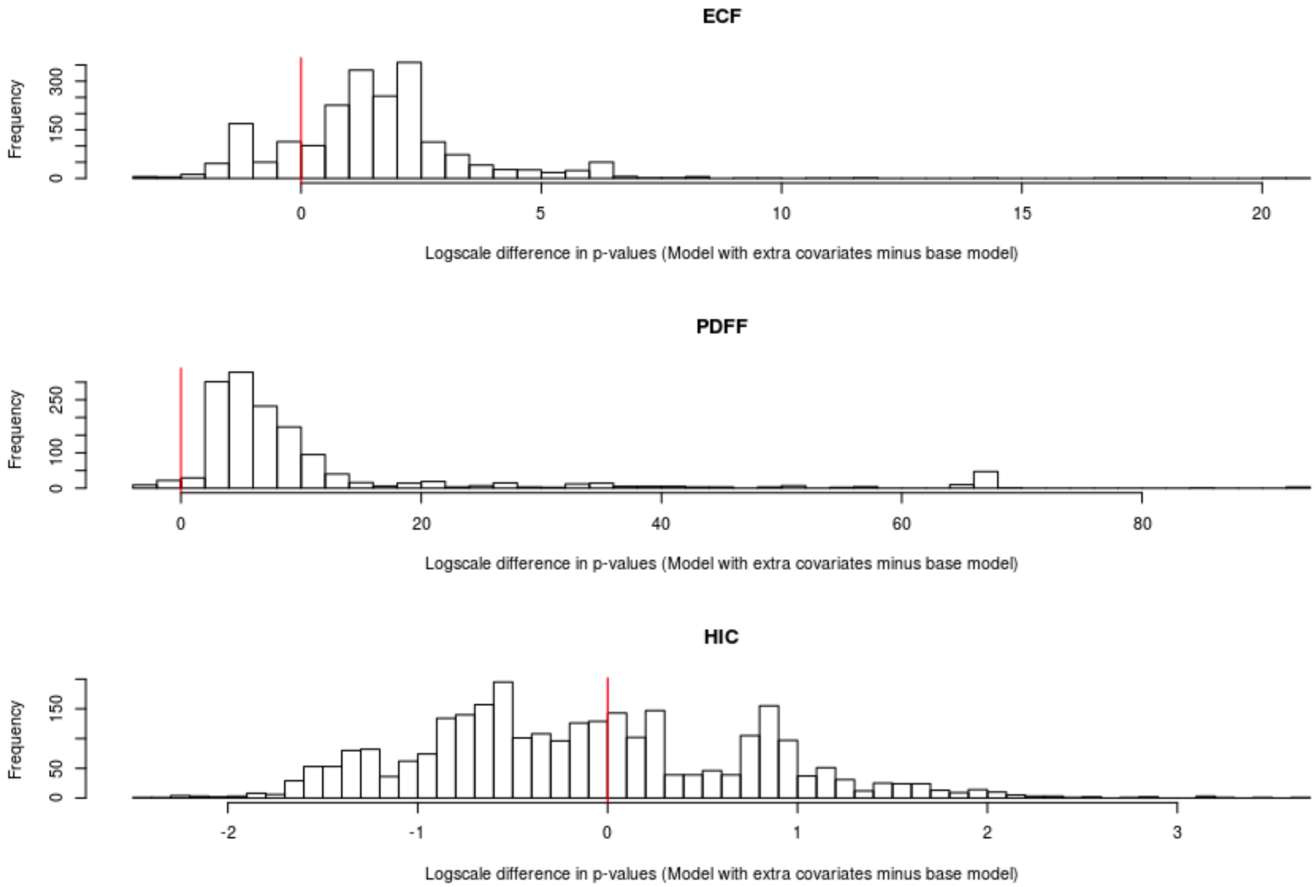

Supplementary Figure 3. Difference in p-values between models adjusted for additional covariates compared to base models.

A vertical red line is shown at 0, where no difference exists in -log10(P) from the two models.

(a) PNPLA3 (PDFF)

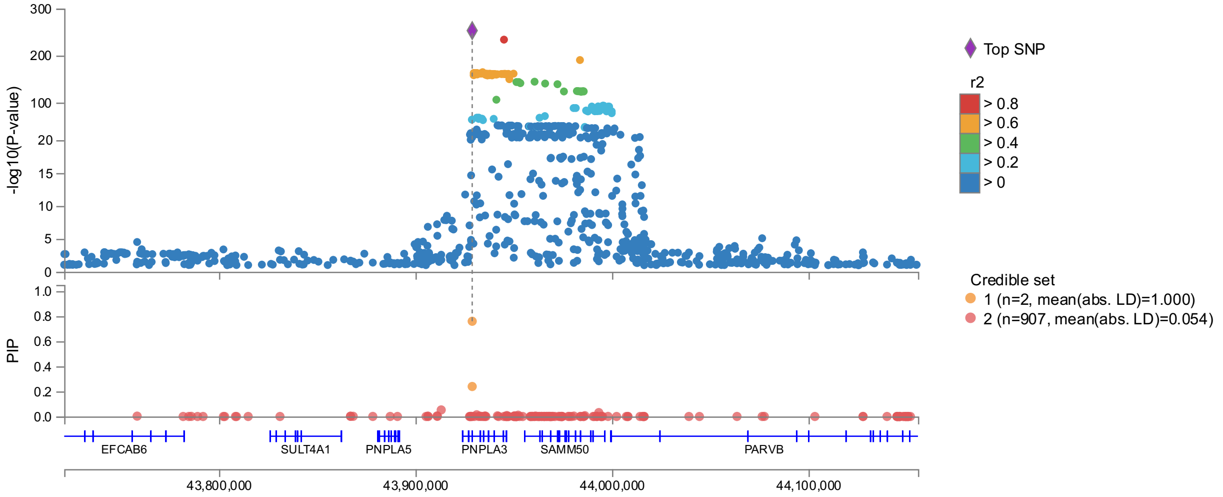

(b) TM6SF2 (PDFF)

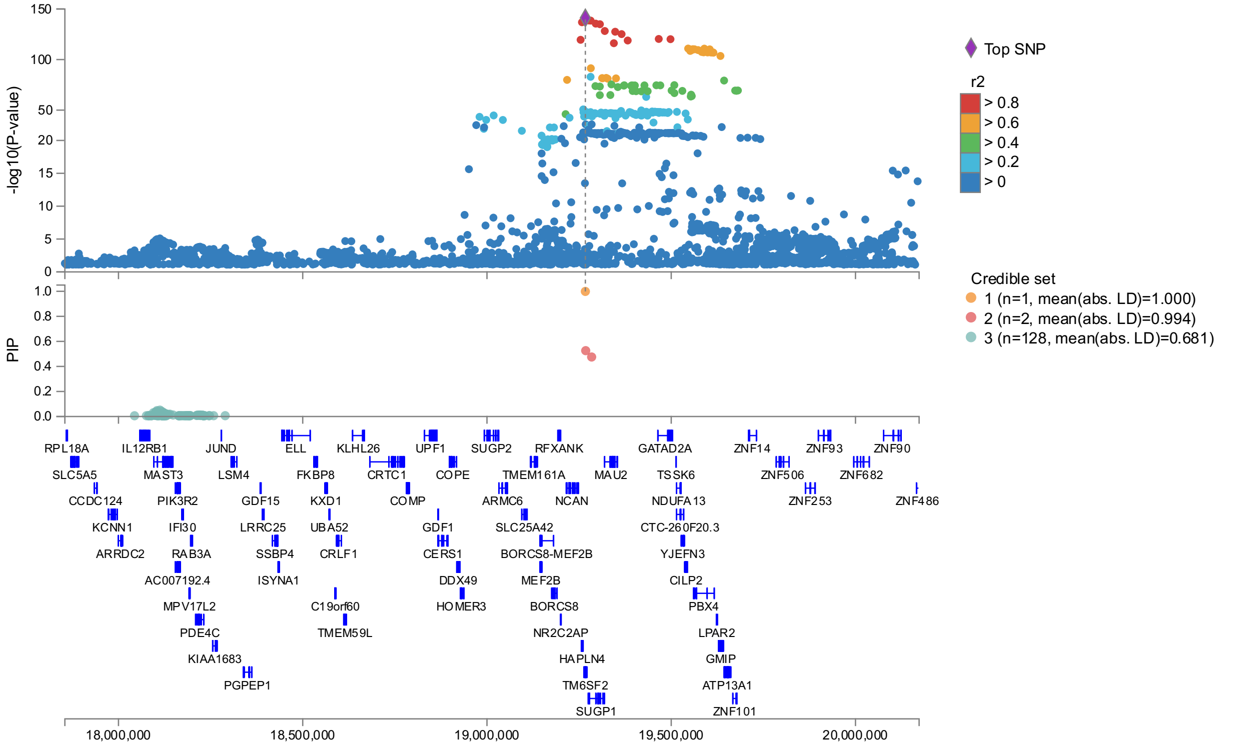

(c) APOE (PDFF)

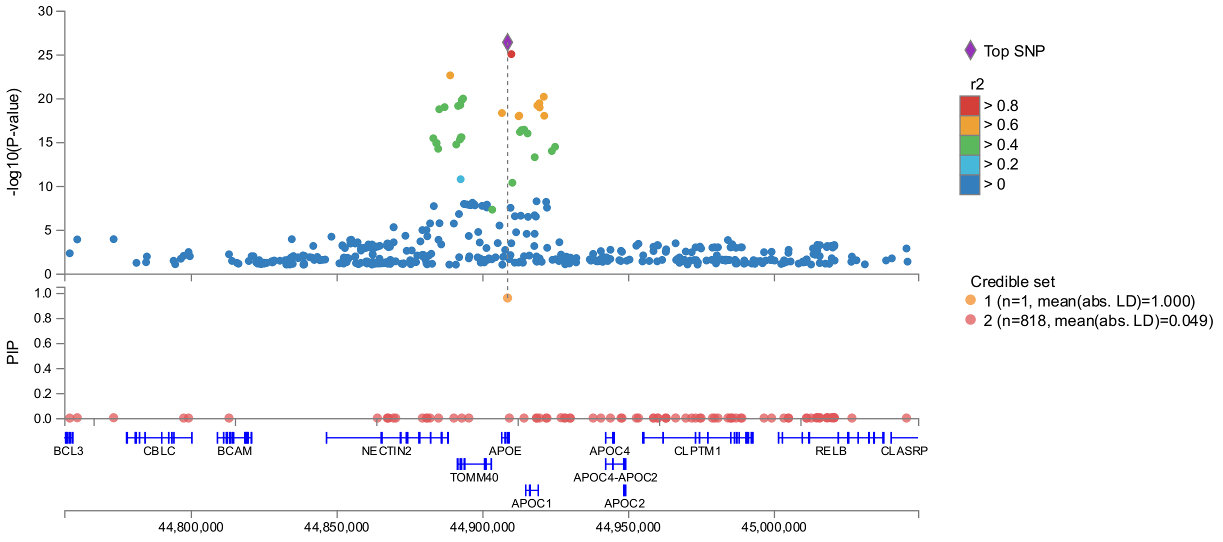

(d) TRIB1 (PDFF)

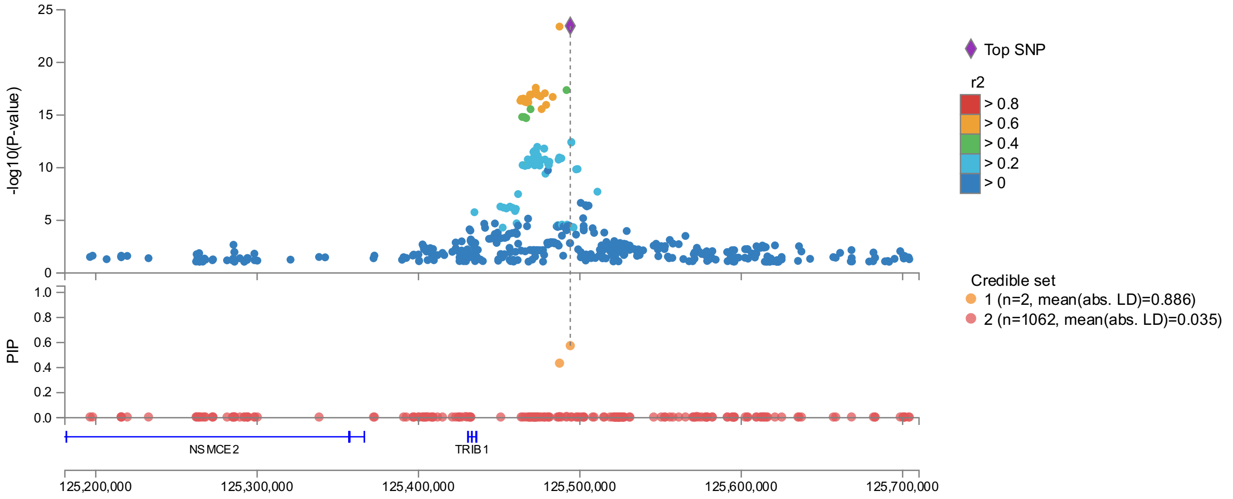

(e) GCKR (PDFF)

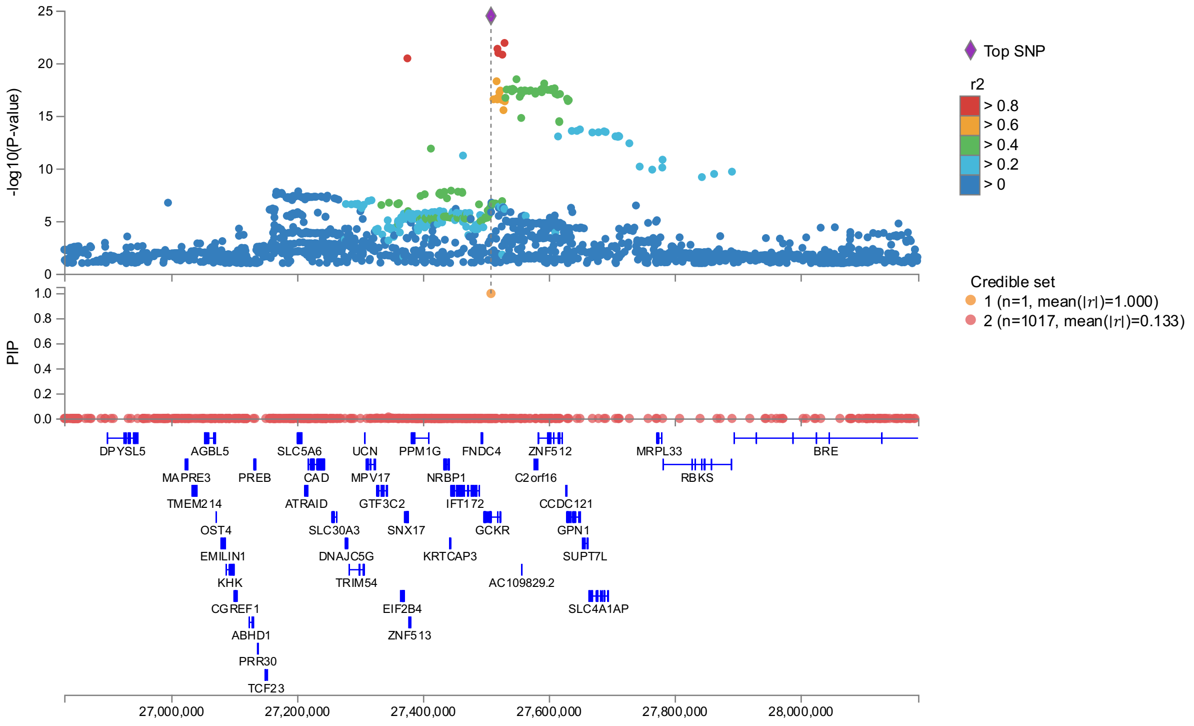

(f) GPAM (PDFF)

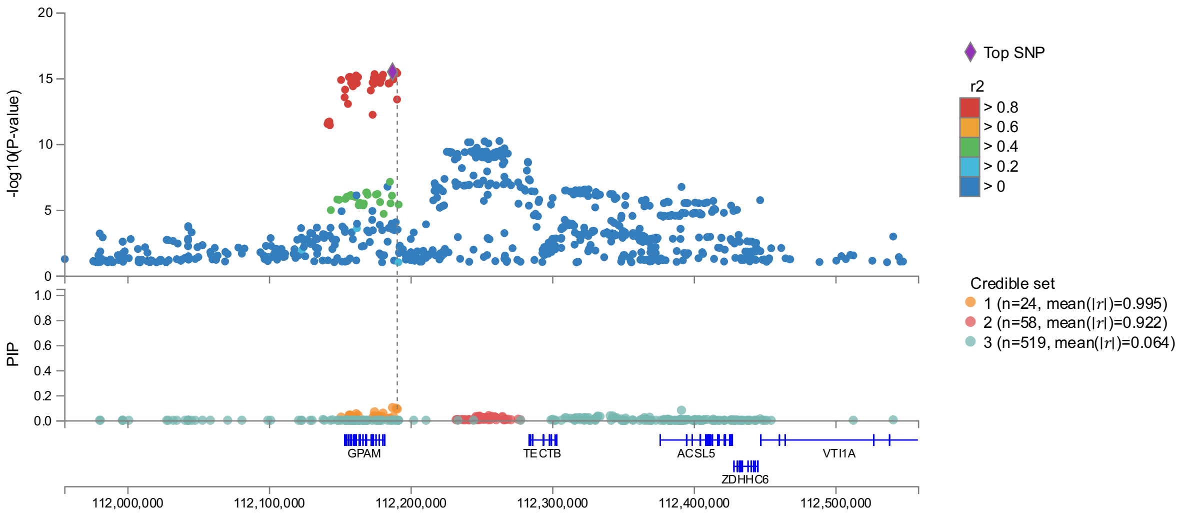

(g) APOH (PDFF)

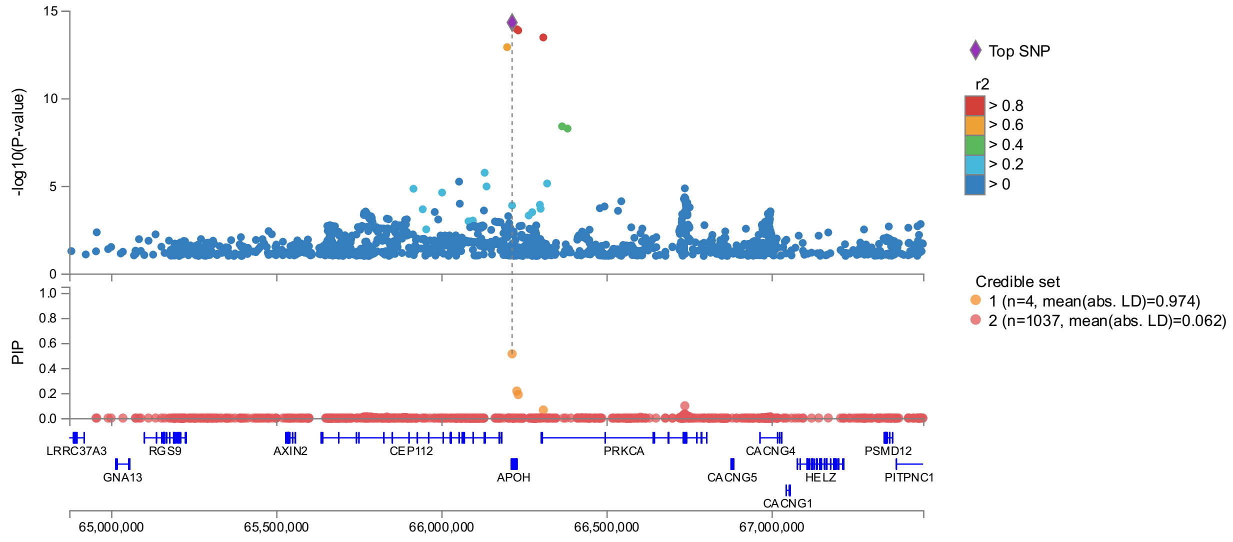

(h) COBLL1 (PDFF)

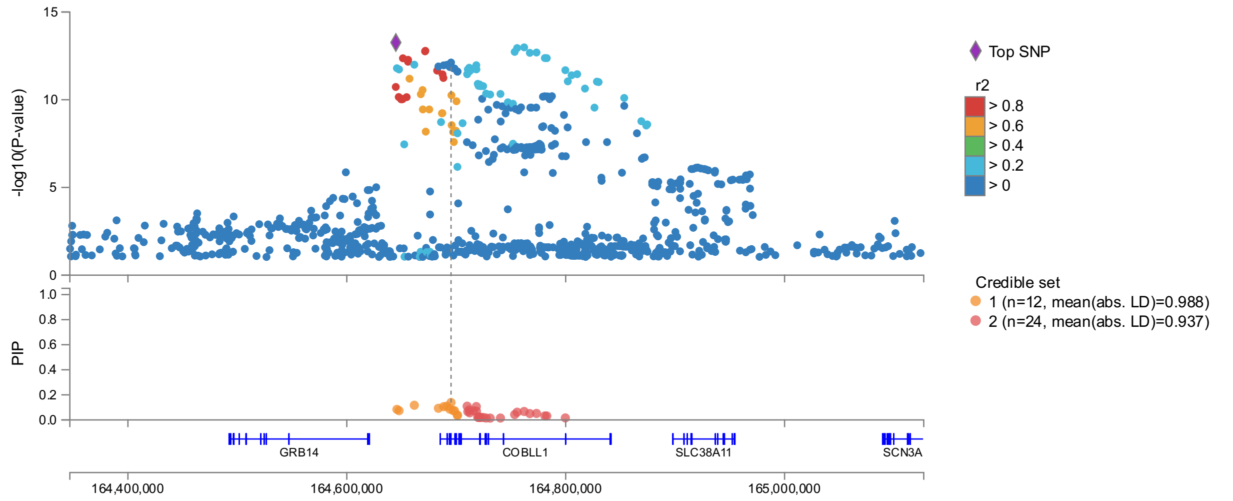

(i) MARC1 (PDFF)

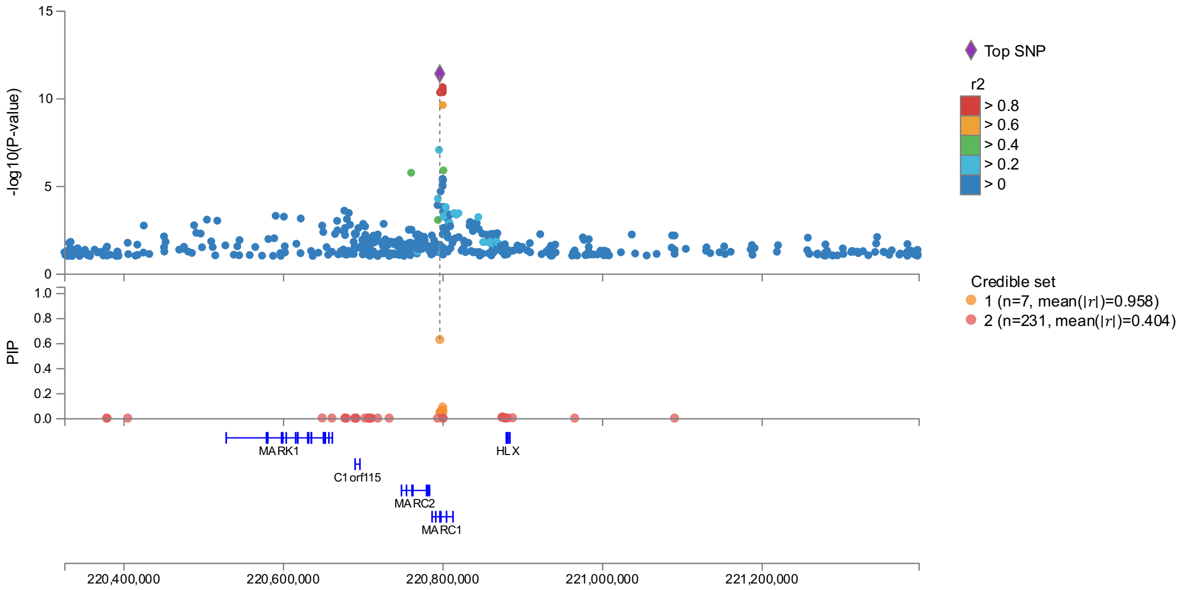

(j) PNPLA2 (PDFF)

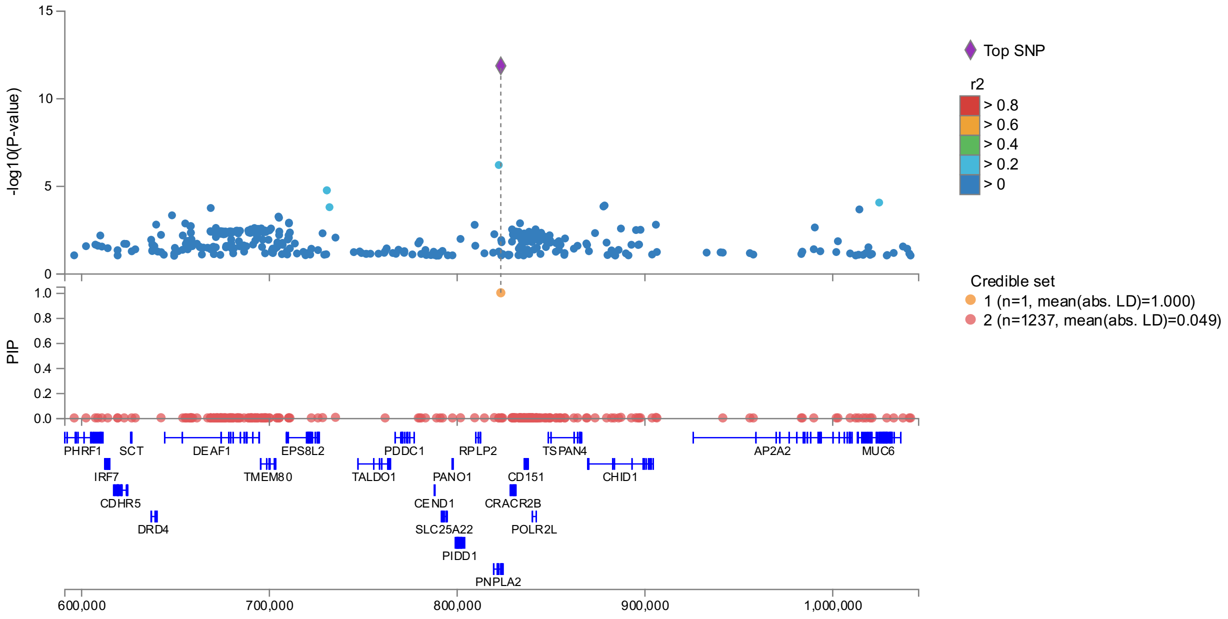

(k) JAZFA (PDFF)

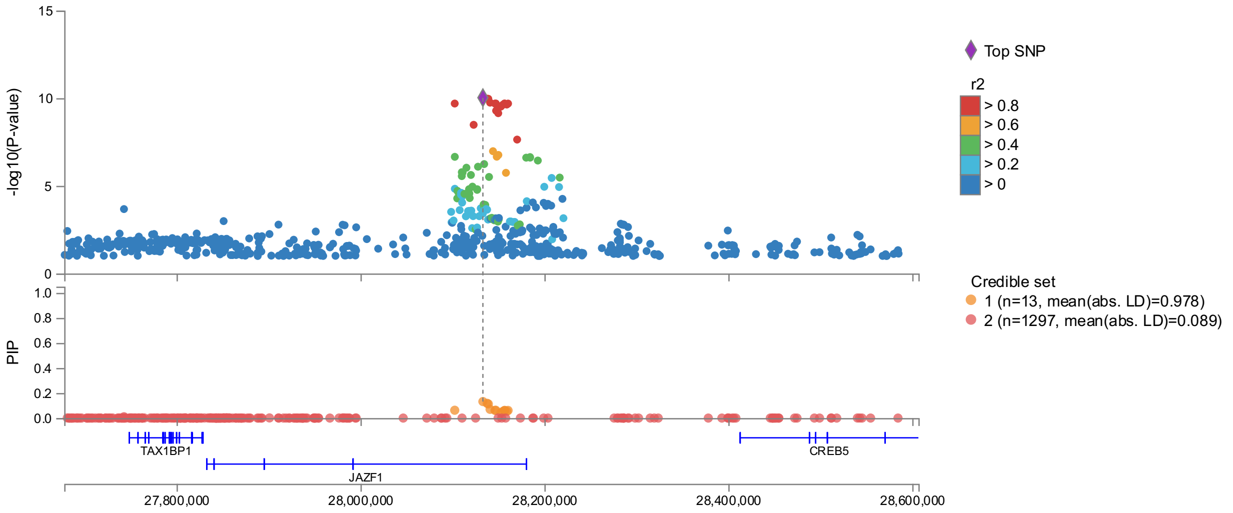

(l) TOR1B (PDFF)

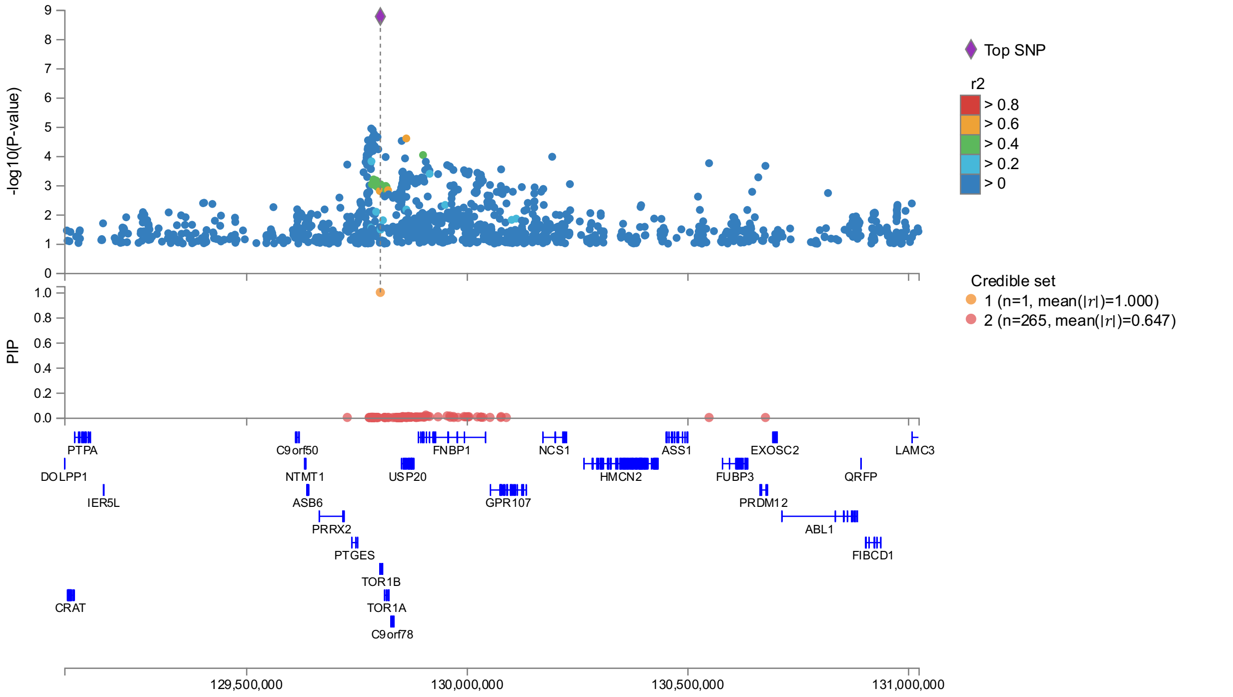

#### (m) ADH1B (PDFF) (interval 2 is under interval 3)

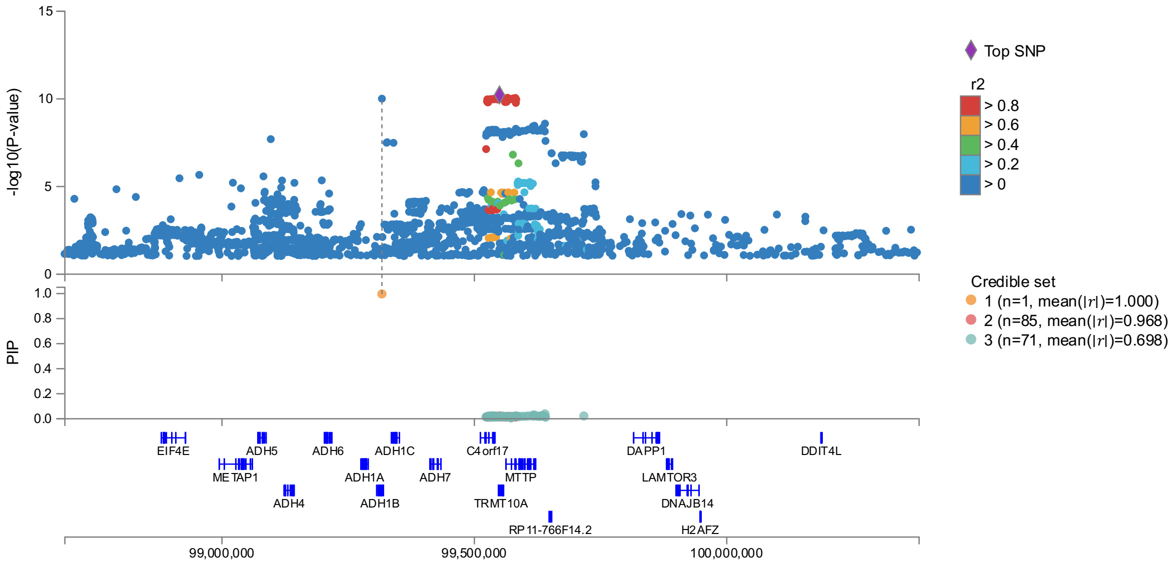

#### (n) ASL (PDFF)

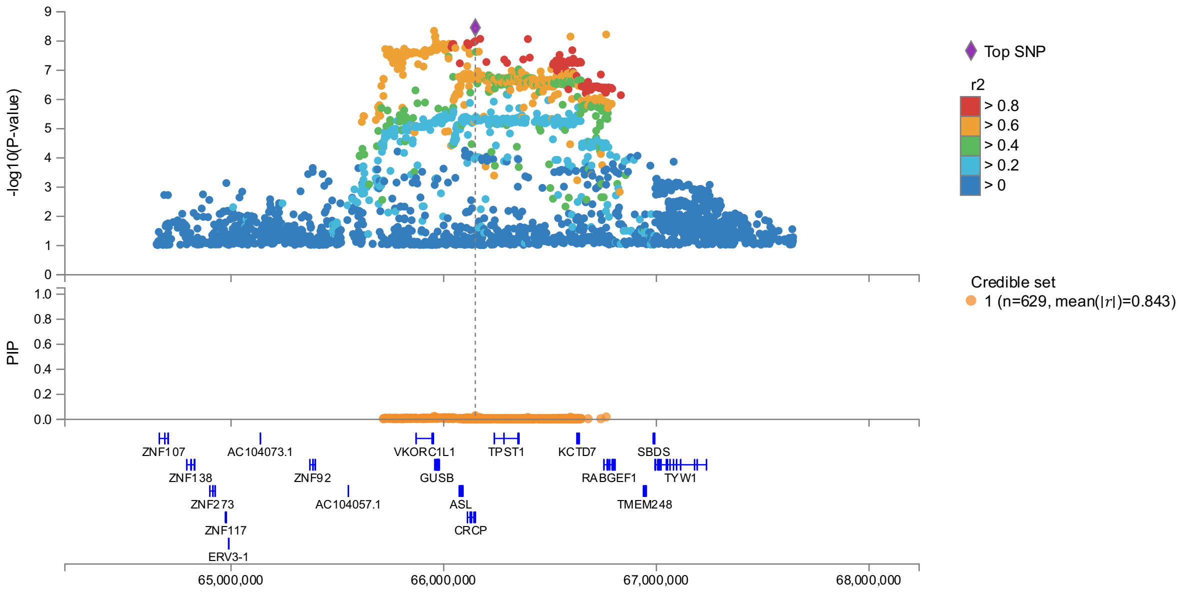

(o) TMC4 (PDFF)

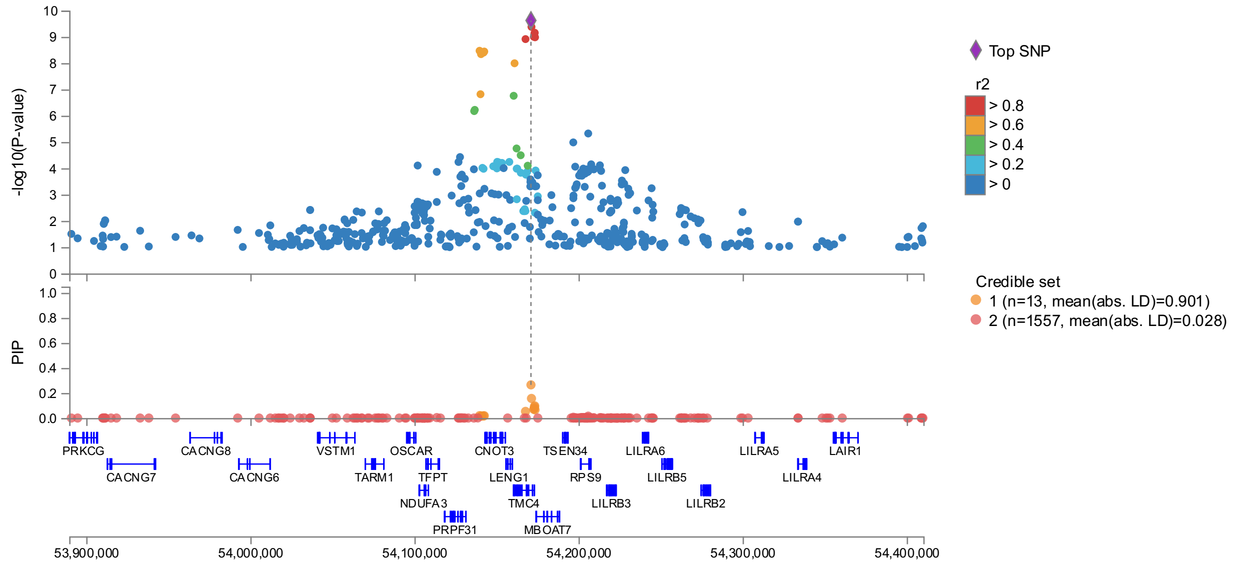

(p) NYAP2 (PDFF)

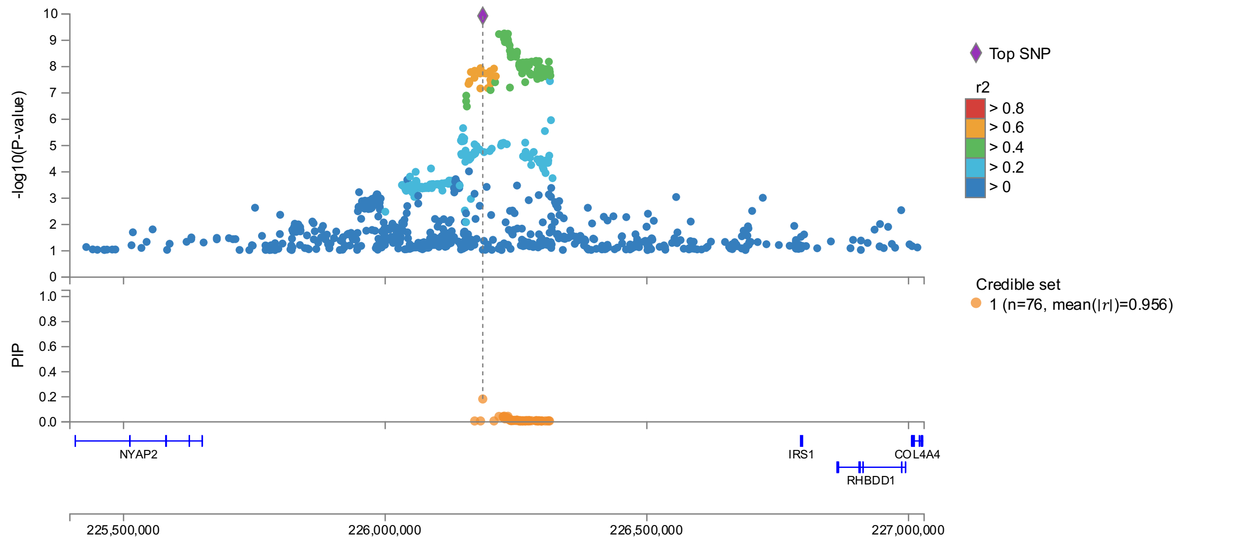

(q) TSC22D2 (PDFF)

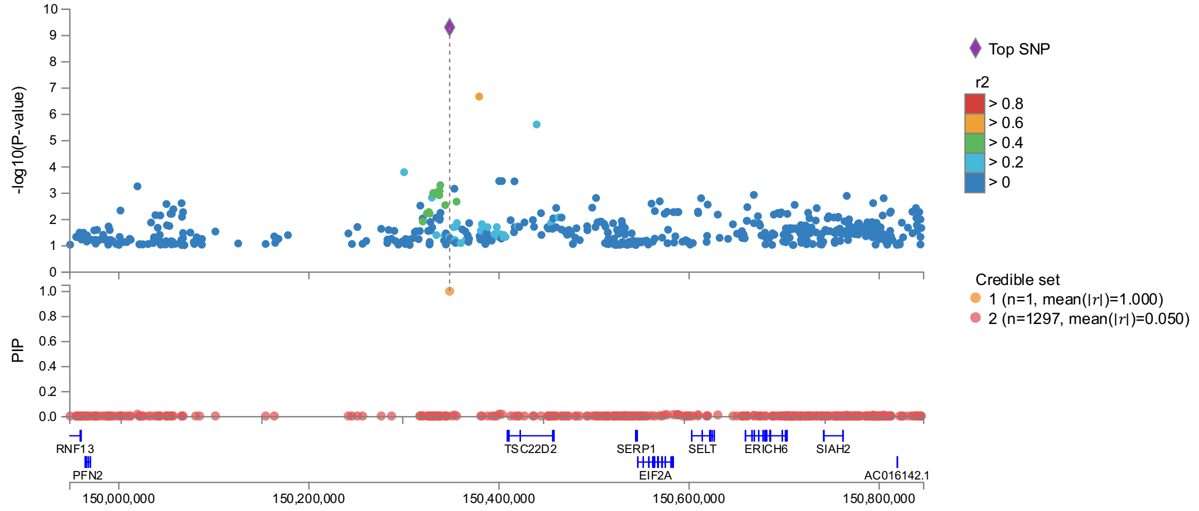

(r) ZNF664 (PDFF)

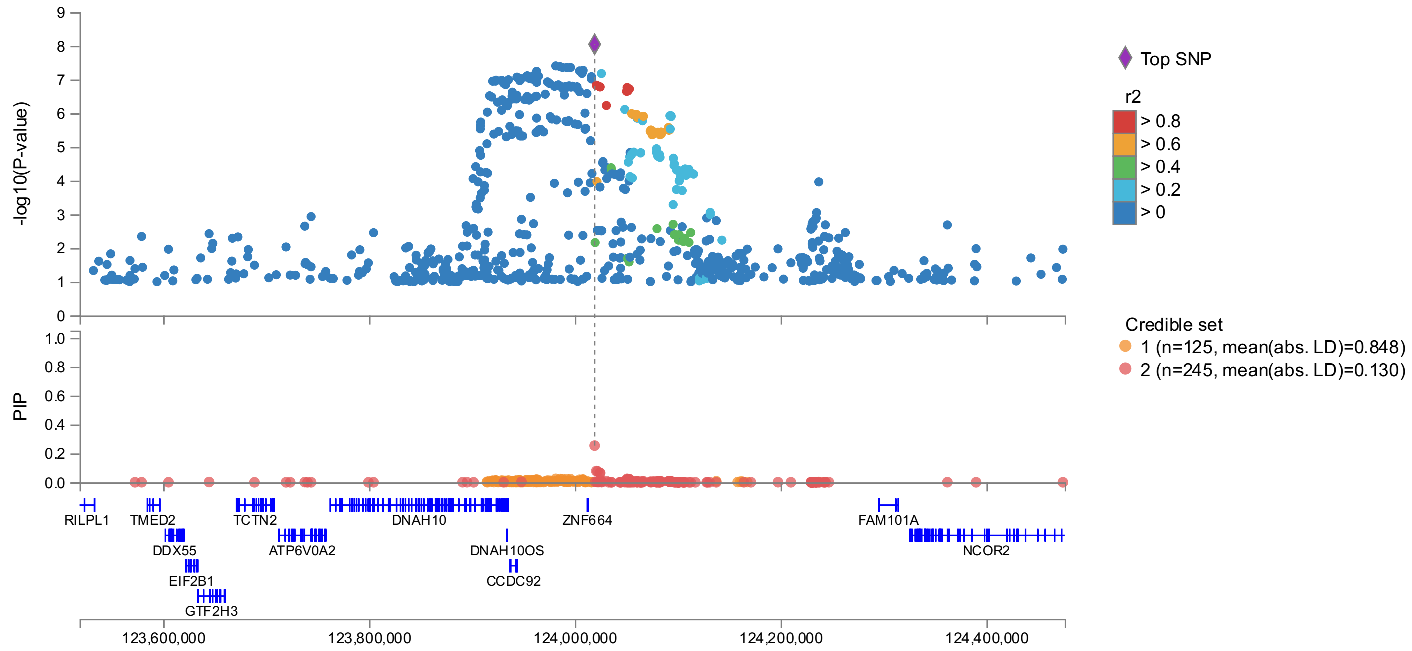

(s) HFE (PDFF)

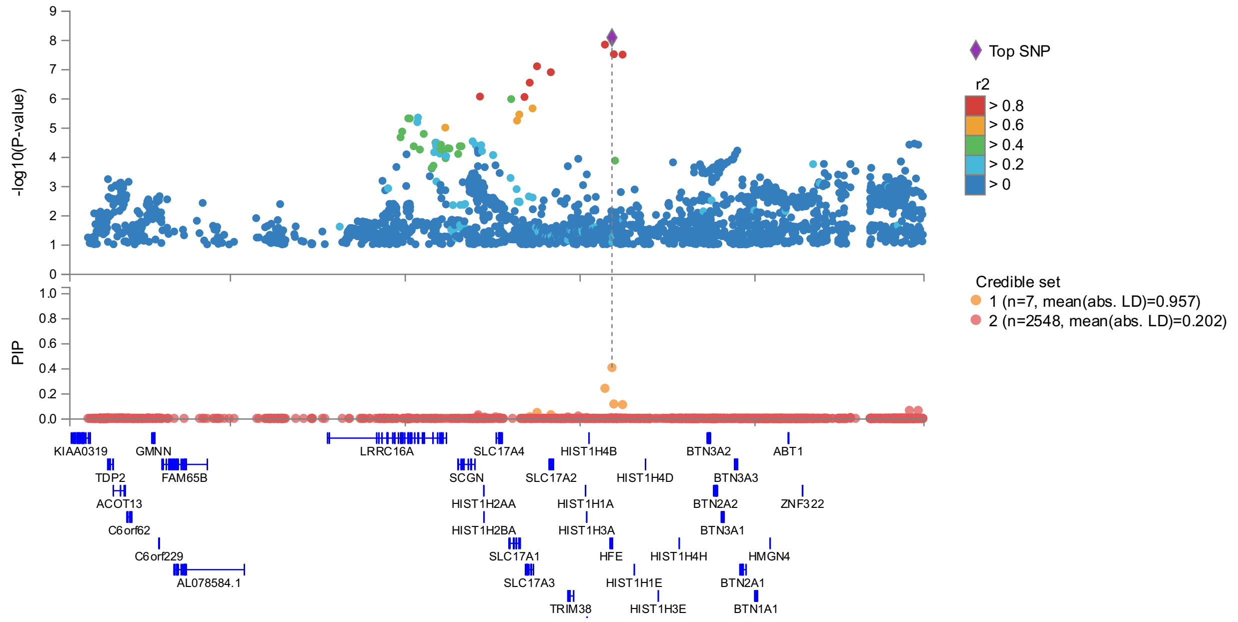

(t) BRCA1 (PDFF)

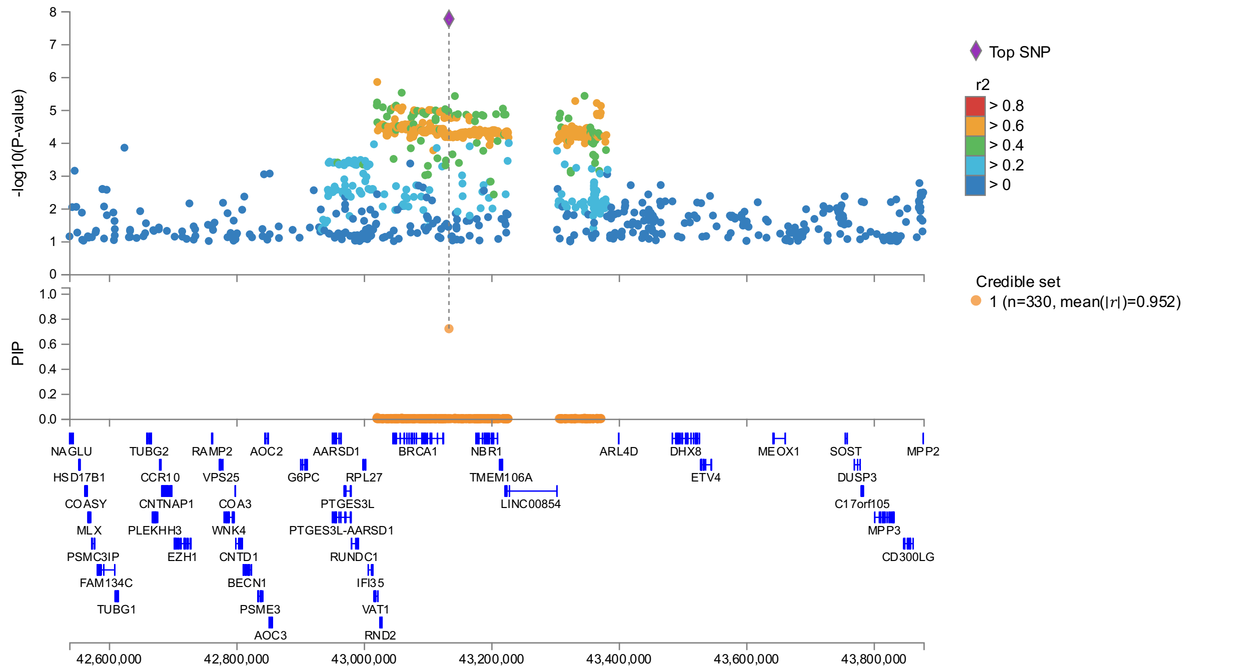

(u) INSR (PDFF)

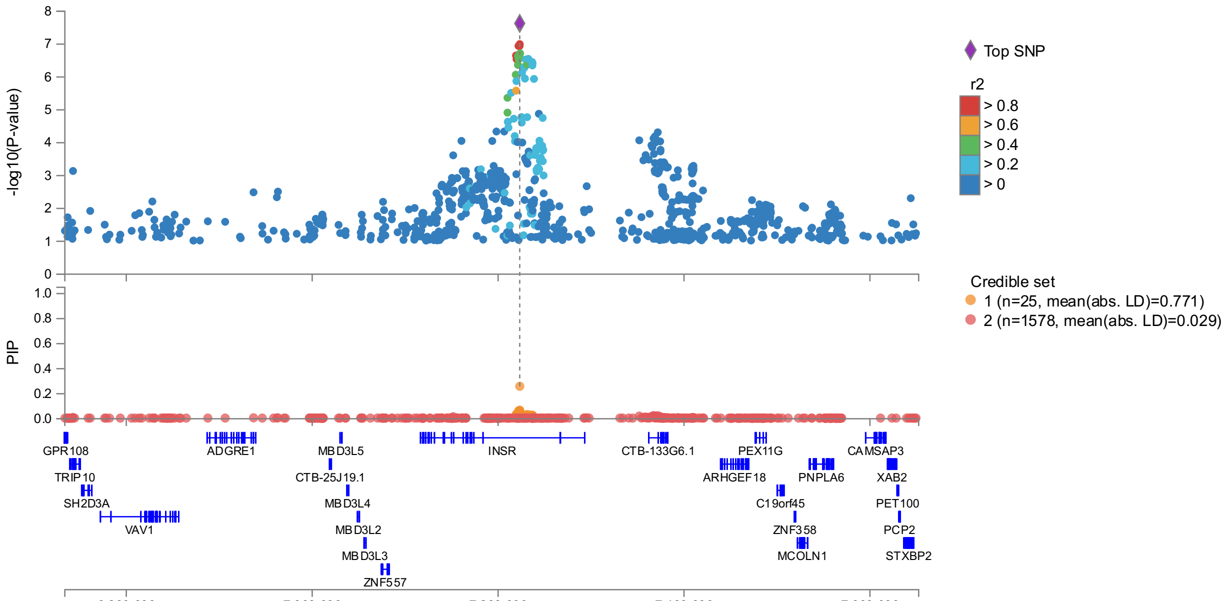

(v) ERLIN1 (PDFF)

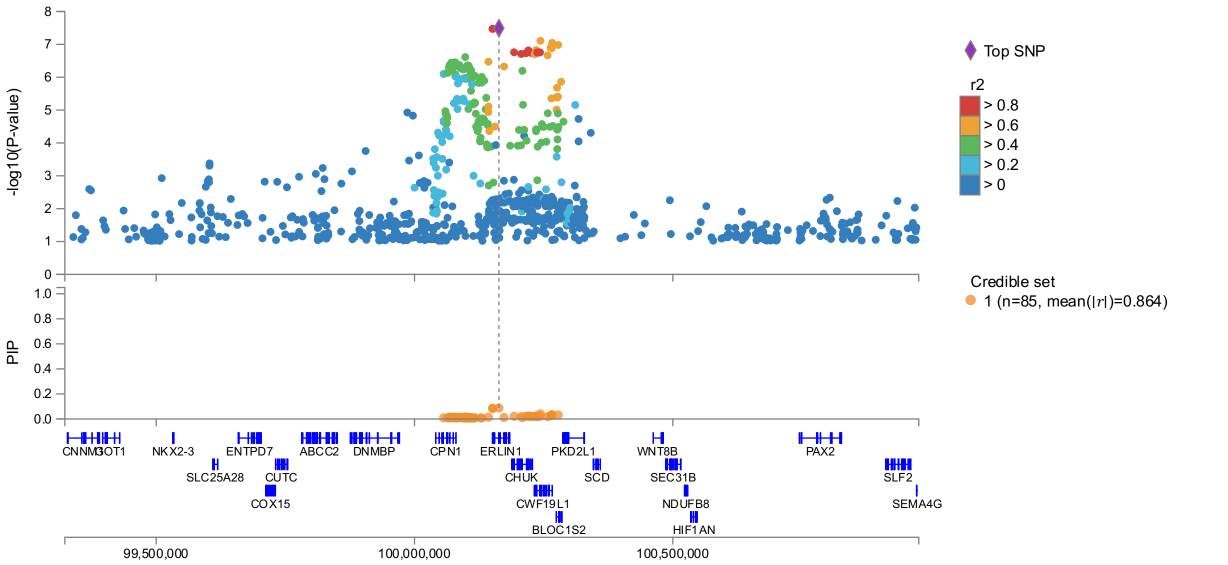

#### Supplementary Figure 4. Regional association plots for PDFF.

Results shown for imputed GWAS analysis and include additional covariate adjustment for BMI and alcohol. FINEMAP estimated credible set results are shown beneath each regional association plot. (a) PNPLA3, (b) TM6SF2, (c) APOE, (d) TRIB1, (e) GCKR, (f) GPAM, (g) APOH, (h) COBLL1, (i) MARC1, (j) PNPLA2, (k) JAZF1, (l) TOR1B, (m) ADH1B, (n) ASL, (o) TMC4, (p) NYAP2, (q) TSC22D2, (r) ZNF664, (s) HFE, (t) BRCA1, (u) INSR, (v) ERLIN1

(a) HFE (HIC)

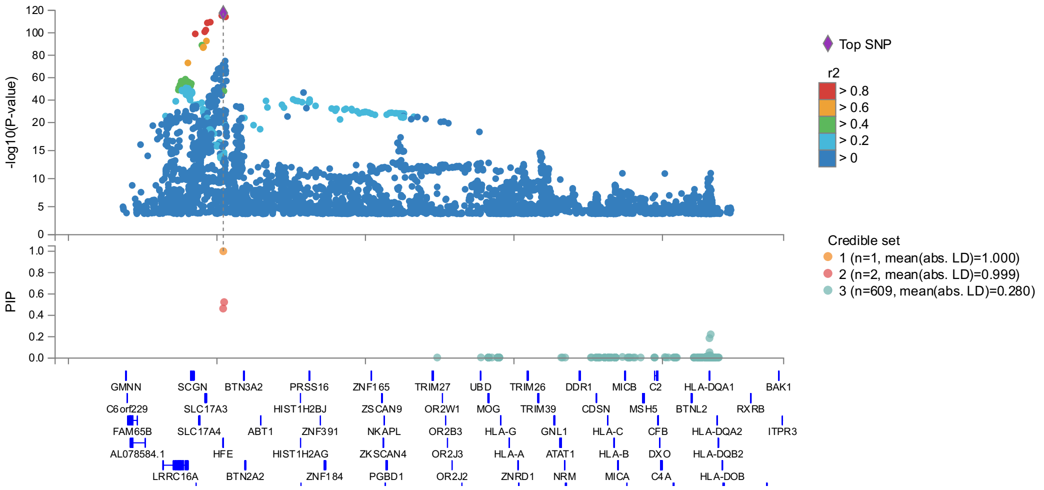

(b) TMPRSS6 (HIC)

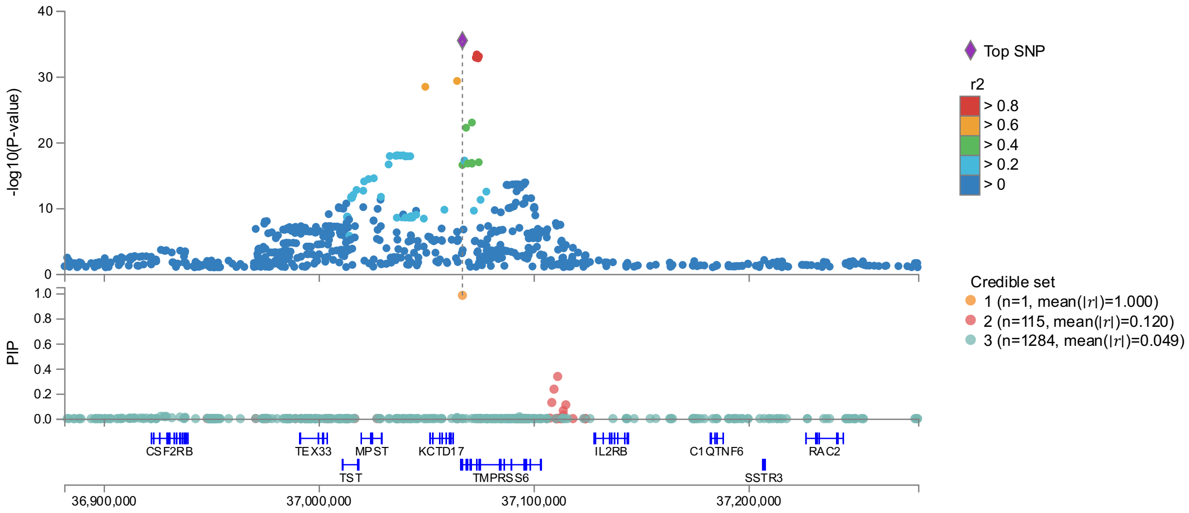

#### (c) ASNSD1 (HIC)

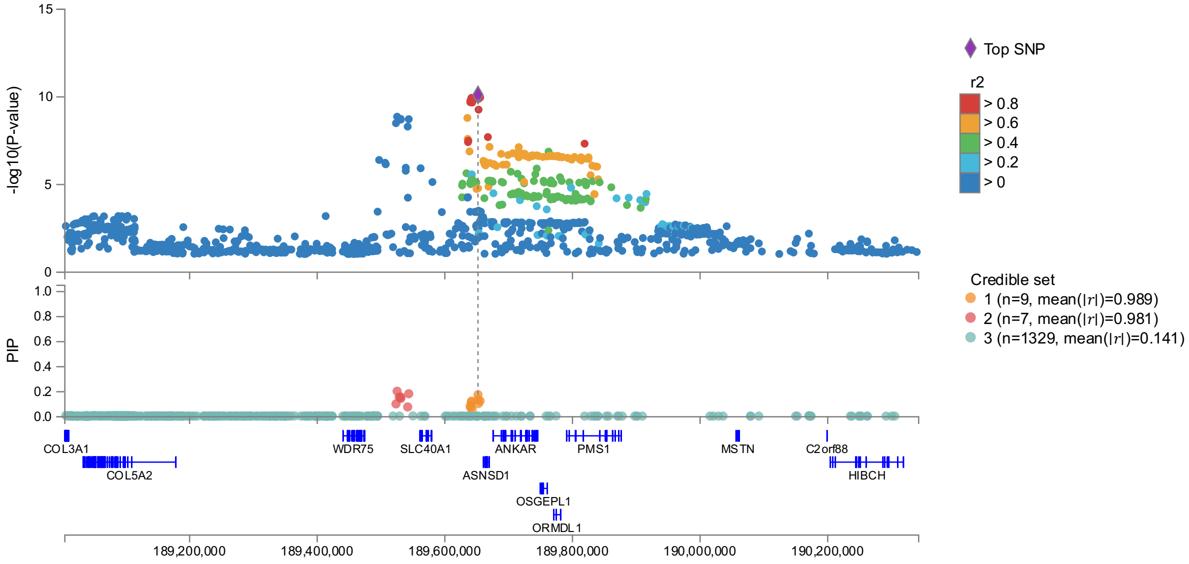

#### (d) GCKR (HIC)

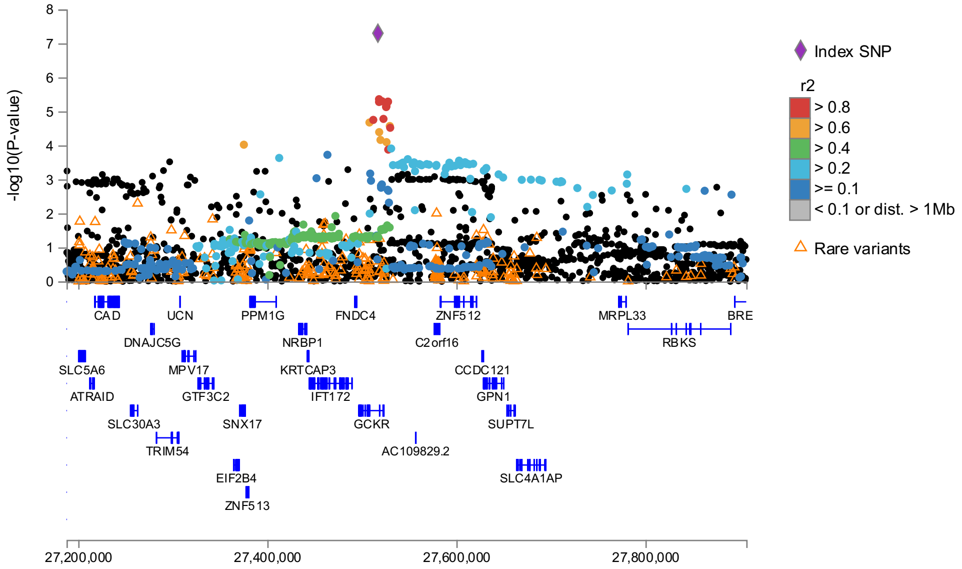

(e) HSF5 (HIC)

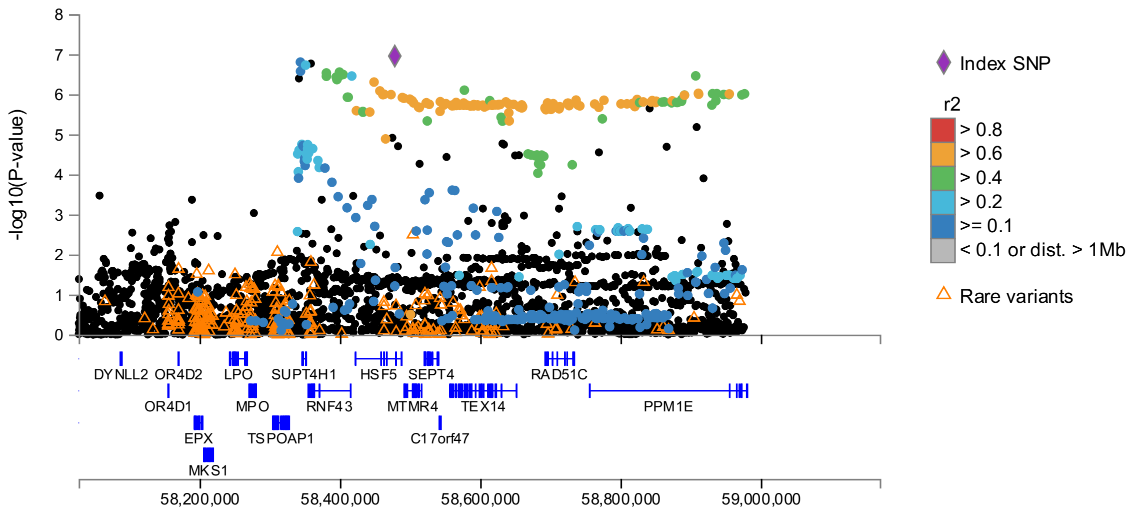

(f) RASIP1 (HIC)

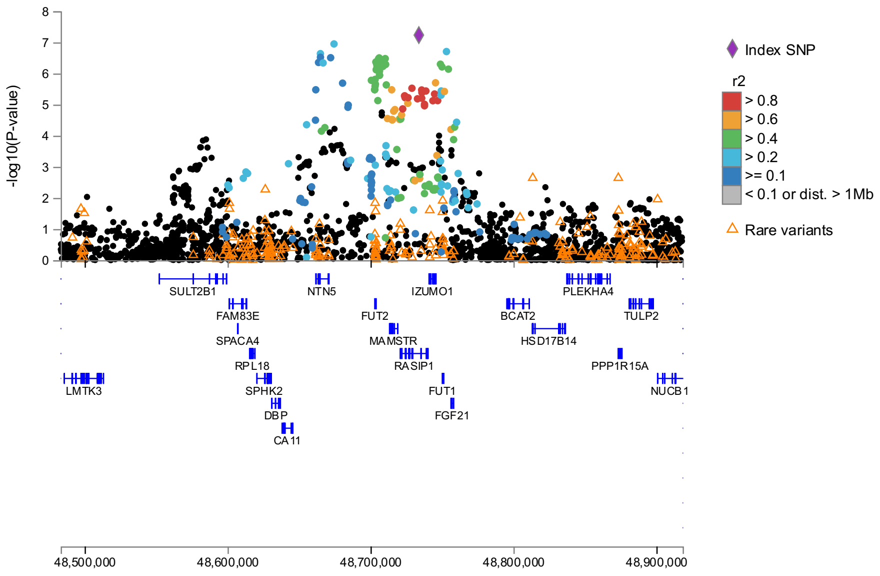

Supplementary Figure 5. **Regional association plots for HIC.**
Results shown for imputed GWAS analysis and include additional covariate adjustment for BMI and alcohol. FINEMAP estimated credible set results are shown beneath each regional association plot. (a) HFE, (b) TMPRSS6, (c) ASNSD1, (d) GCKR, (e) HSF5, (f) RASIP1

(a) SLC39A8 (ECF)

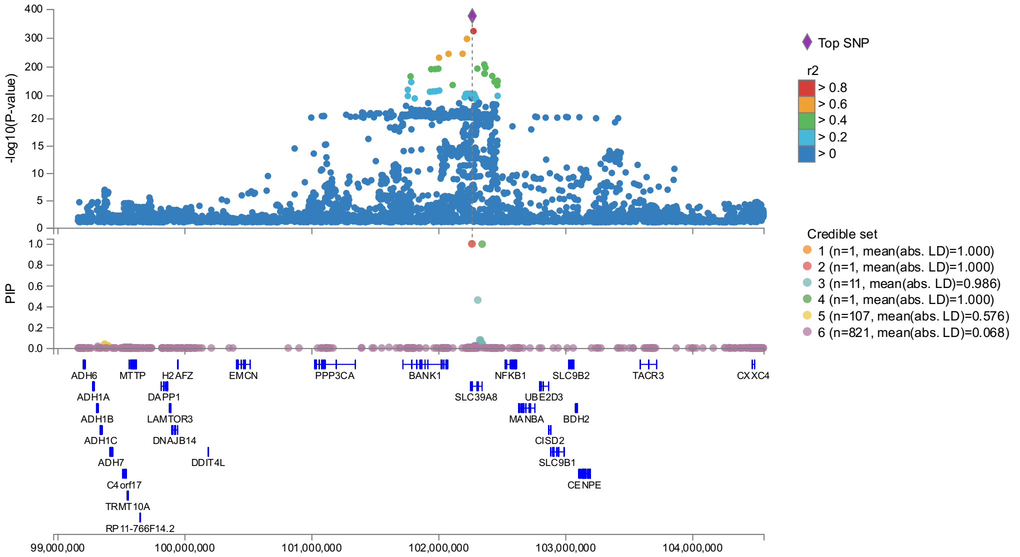

(b) PCK2 (ECF)

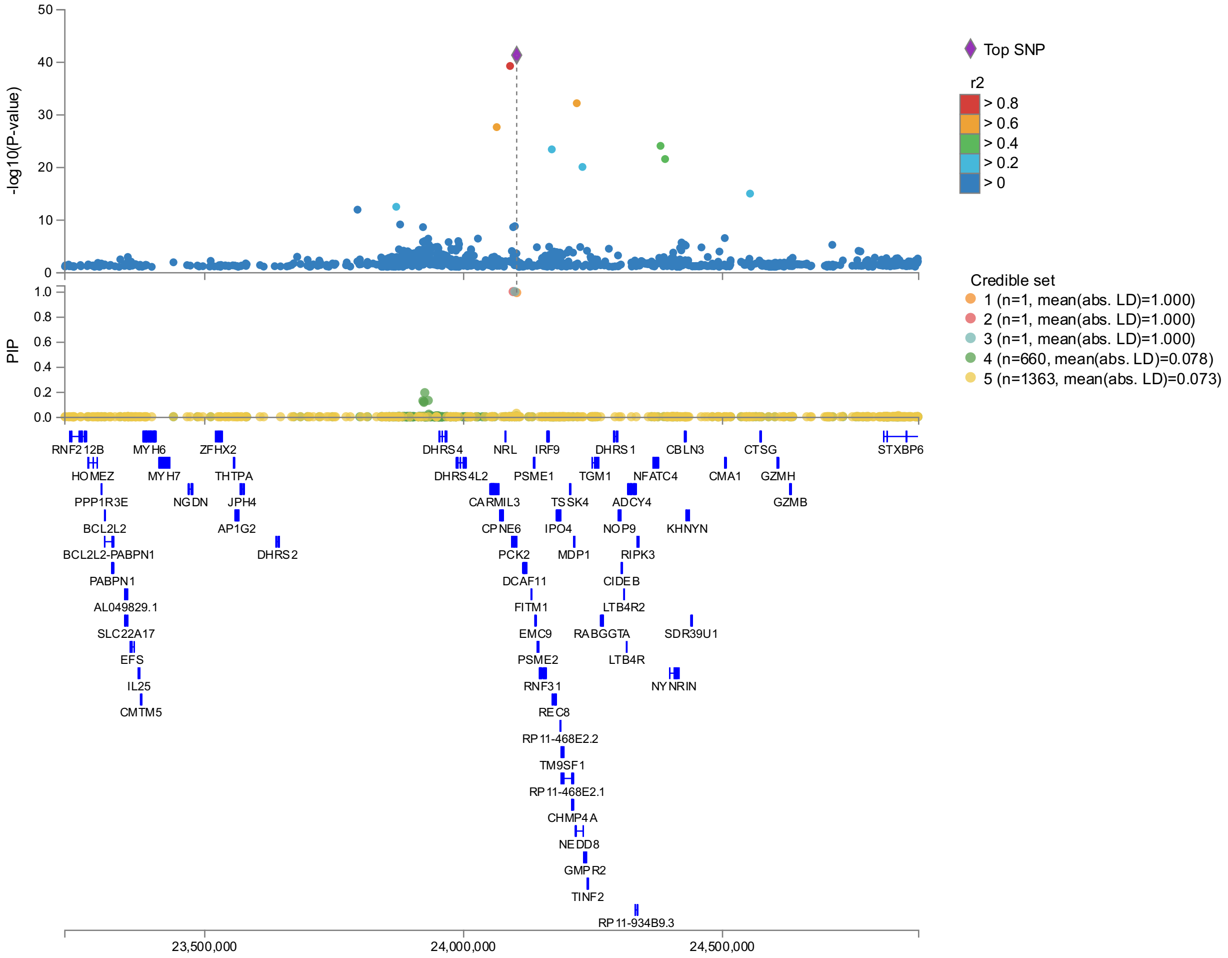

(c) PNPLA3 (ECF)

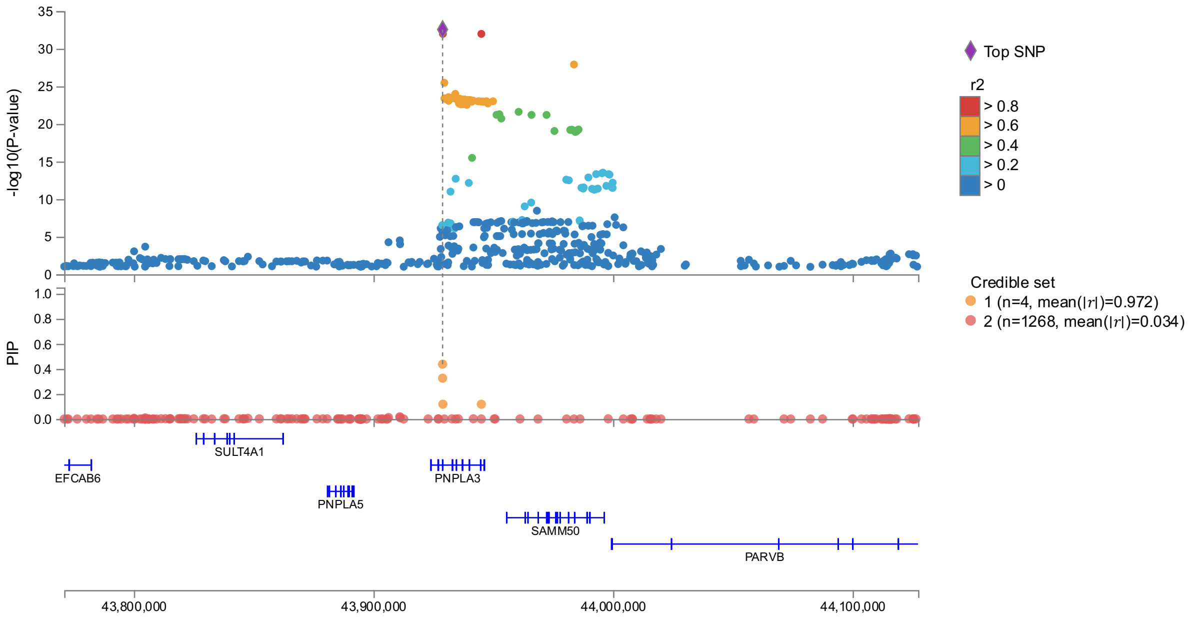

(d) ABO (ECF)

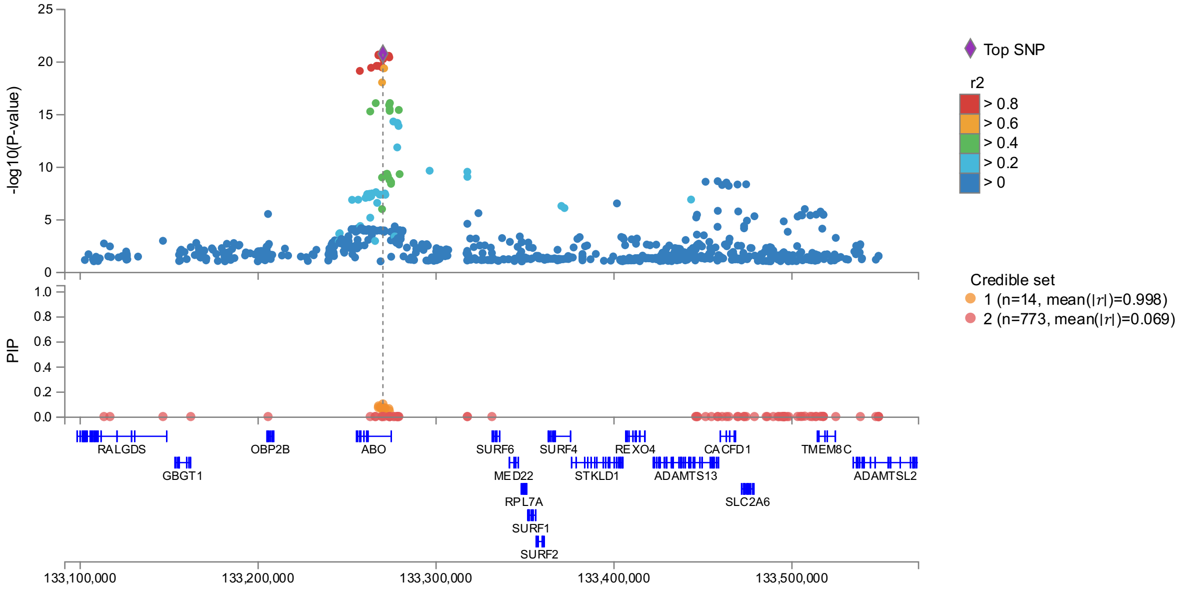

#### (e) SLC30A10 (ECF)

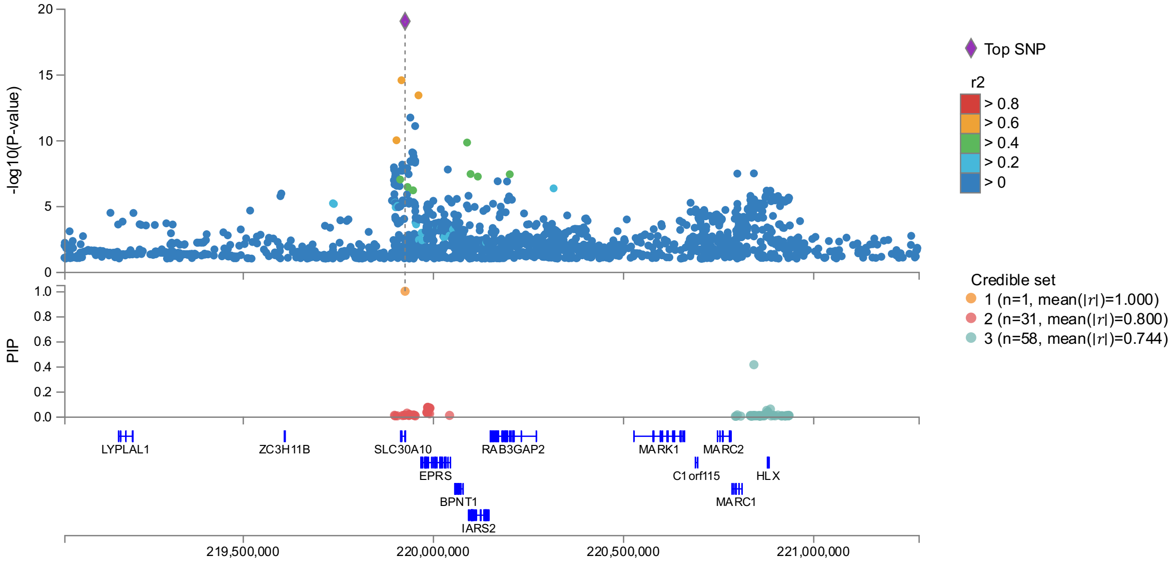

#### (f) TM6SF2 (ECF)

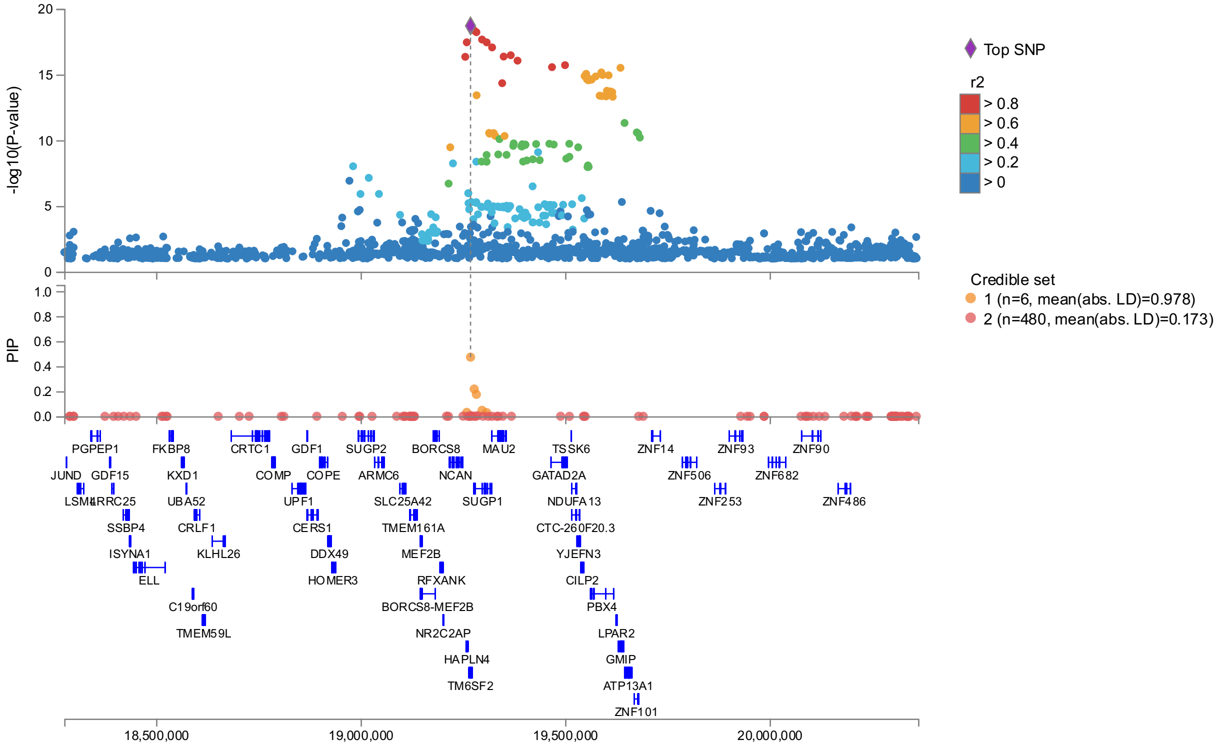

#### (g) HFE (ECF)

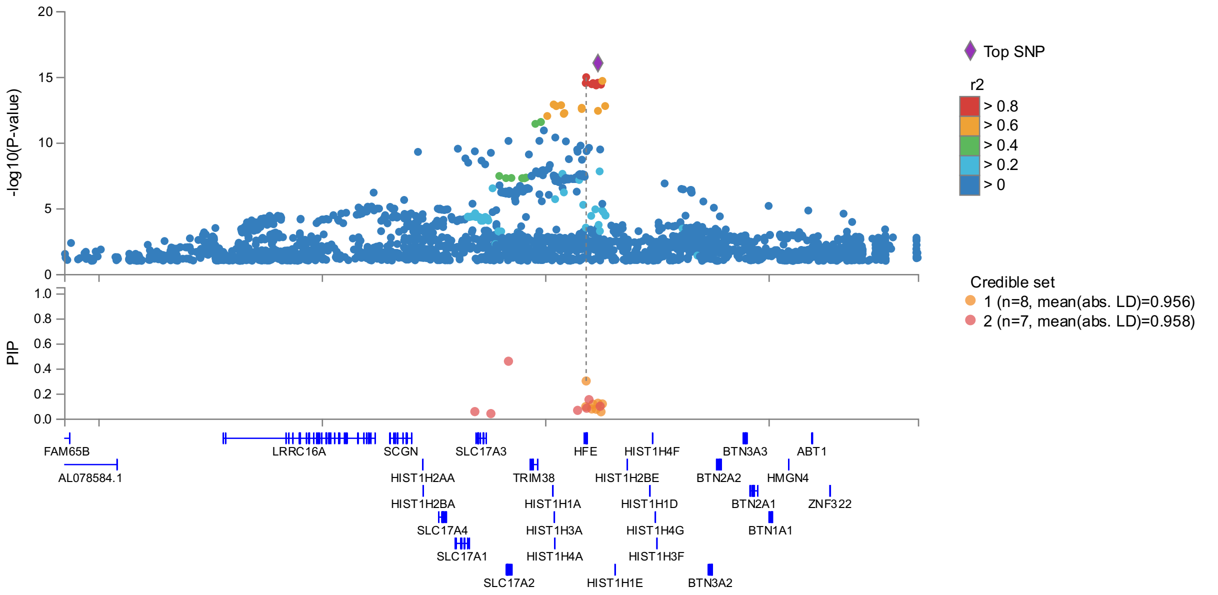

#### (h) TMPRSS6 (ECF)

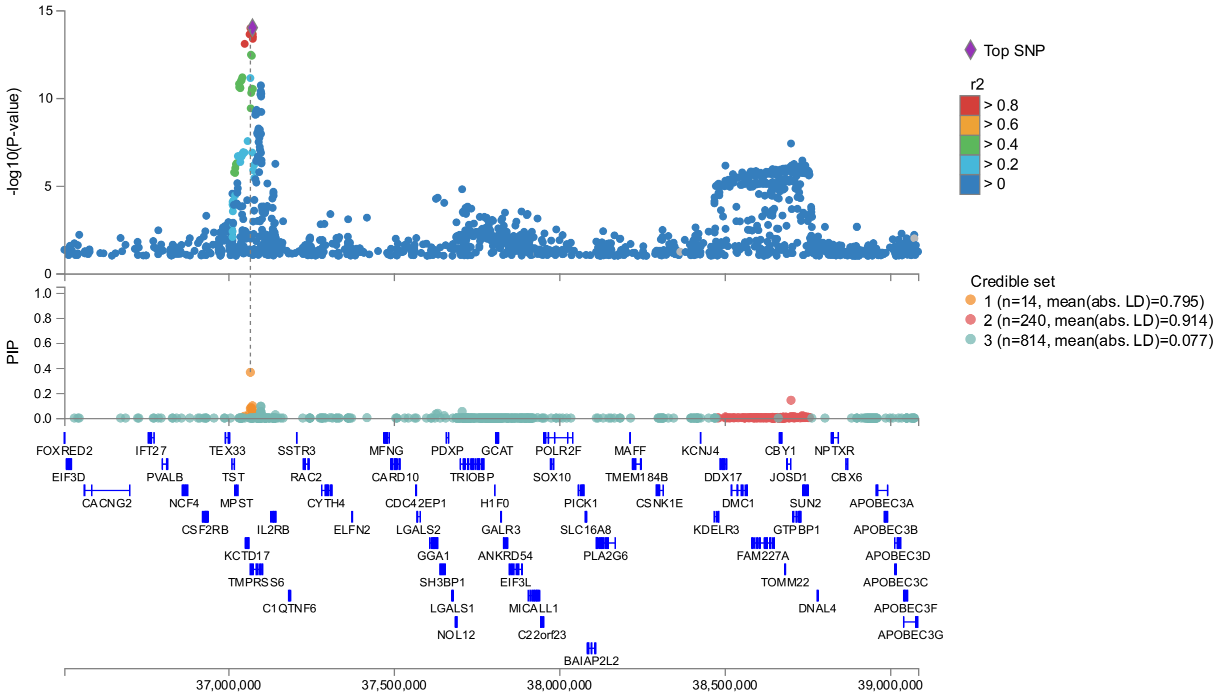

(i) FADS1 (ECF)

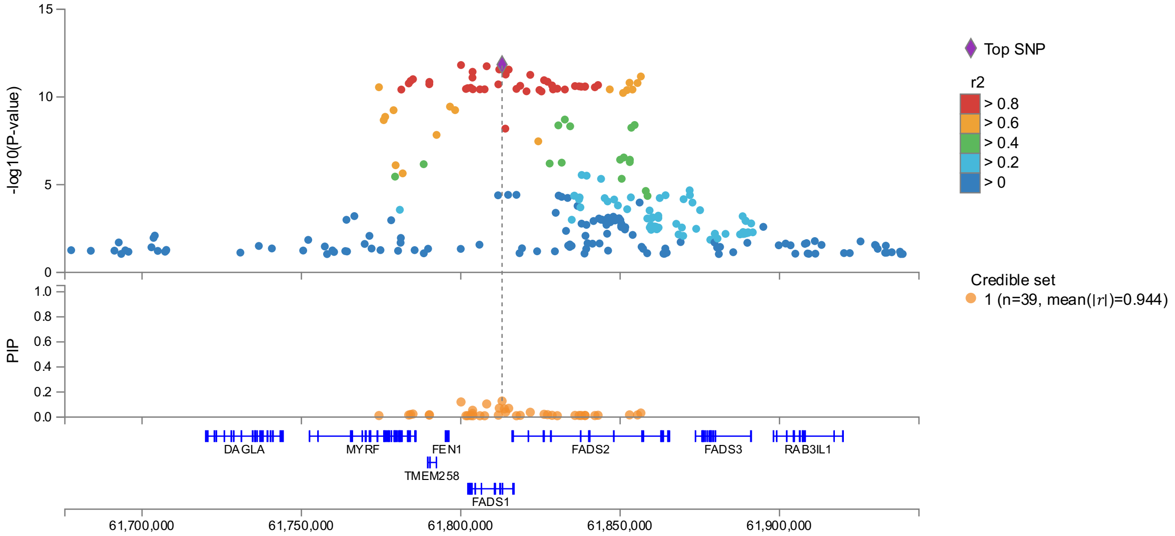

(j) FUT2 (ECF)

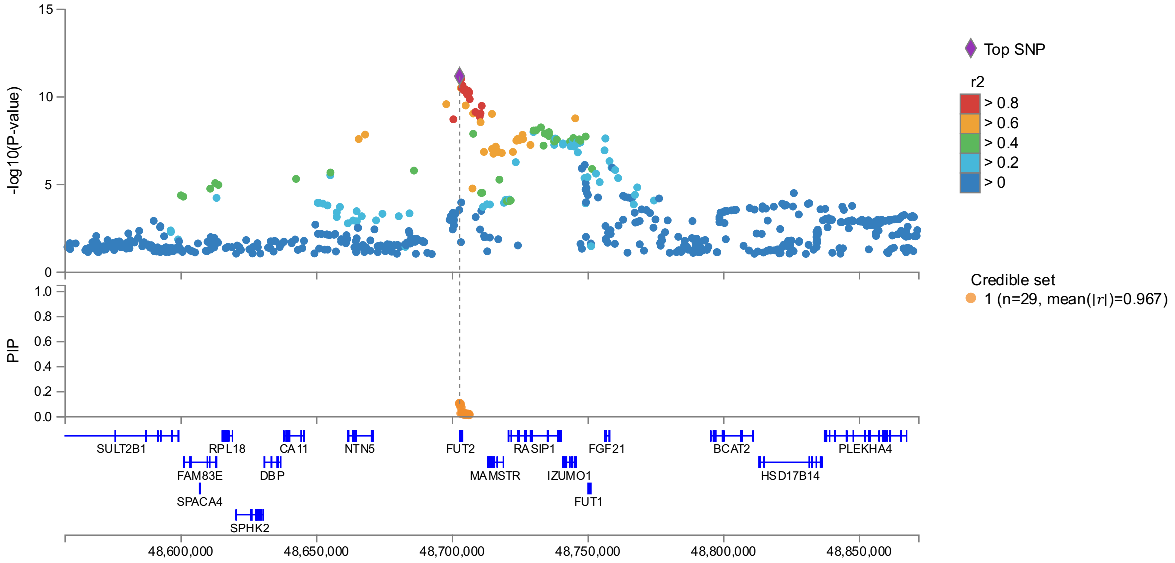

(k) NAT2 (ECF)

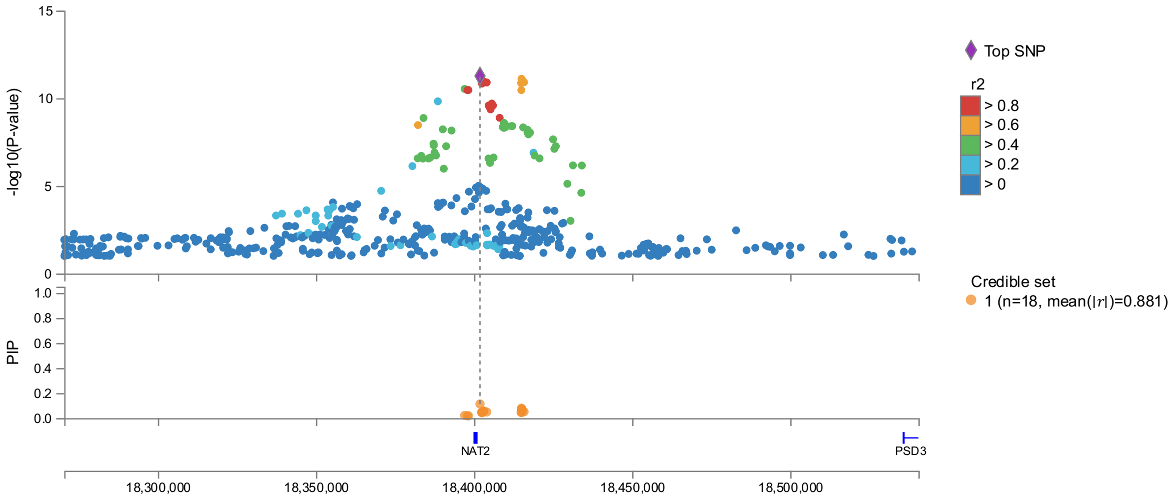

(l) AGMAT (ECF)

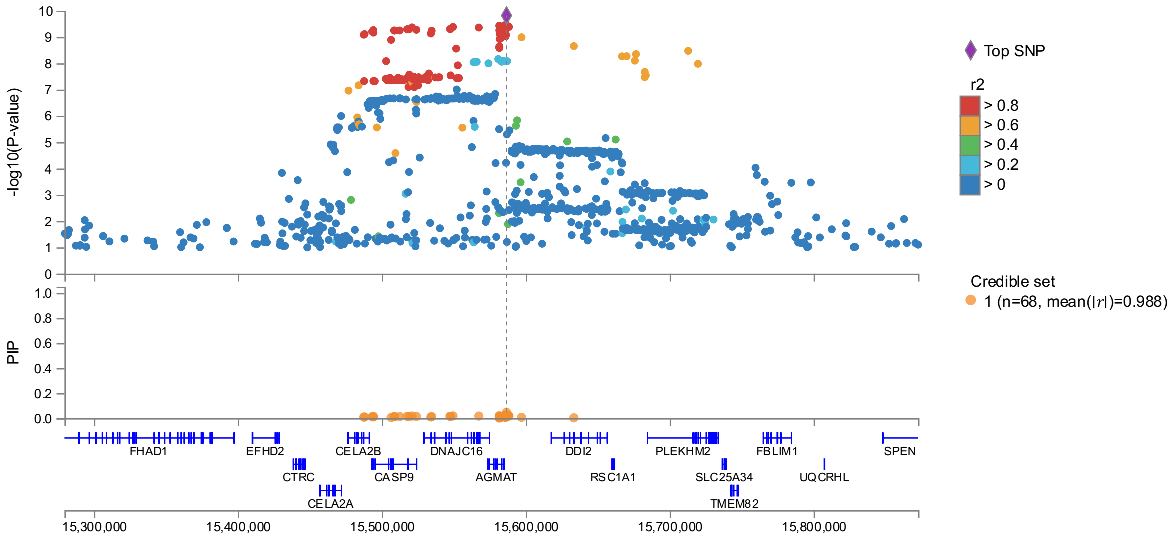

(m) MRPL4 (ECF)

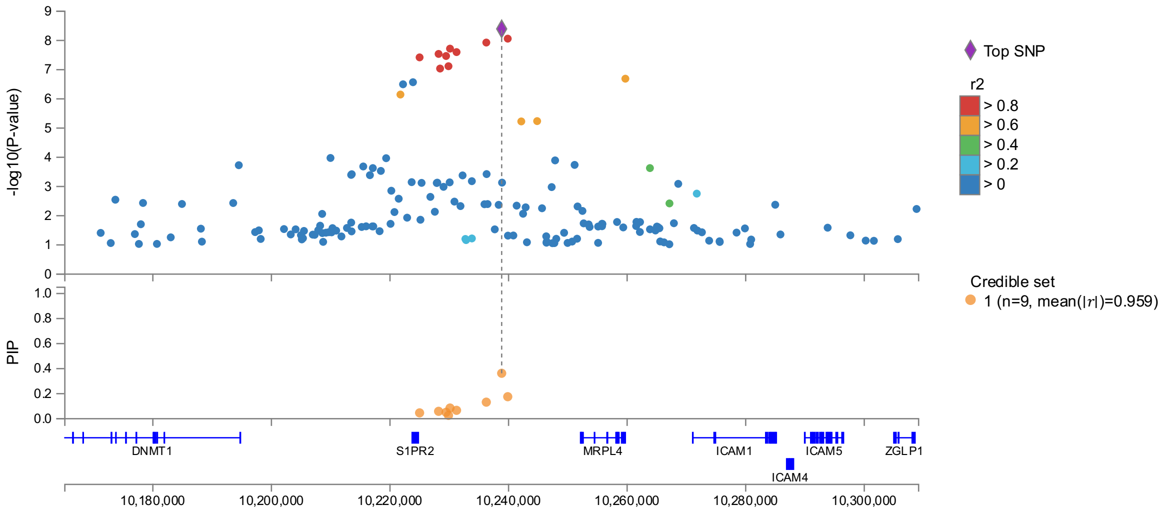

(n) SETBL1 (ECF)

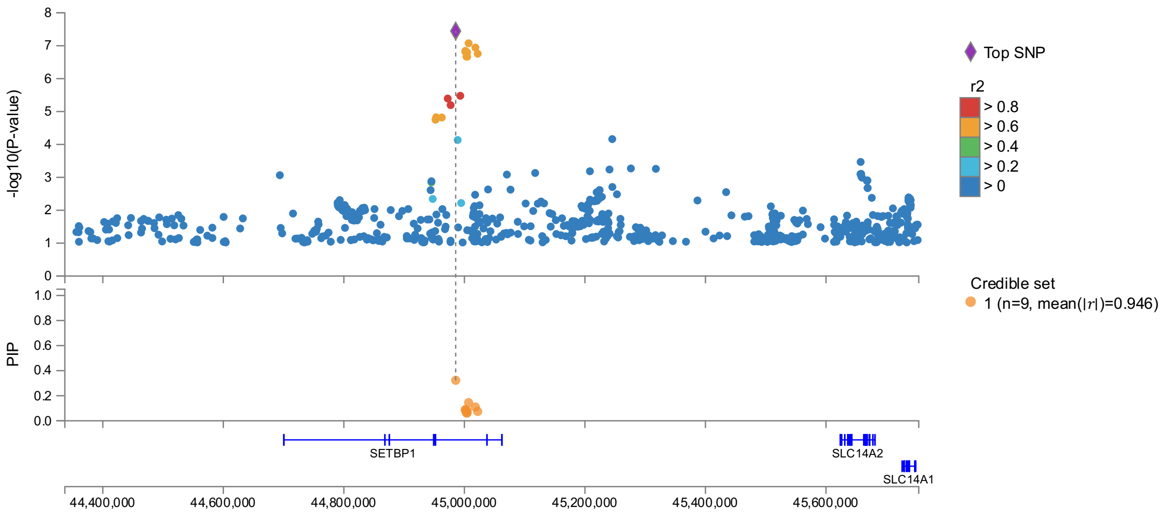

(o) JOSD1 (ECF)

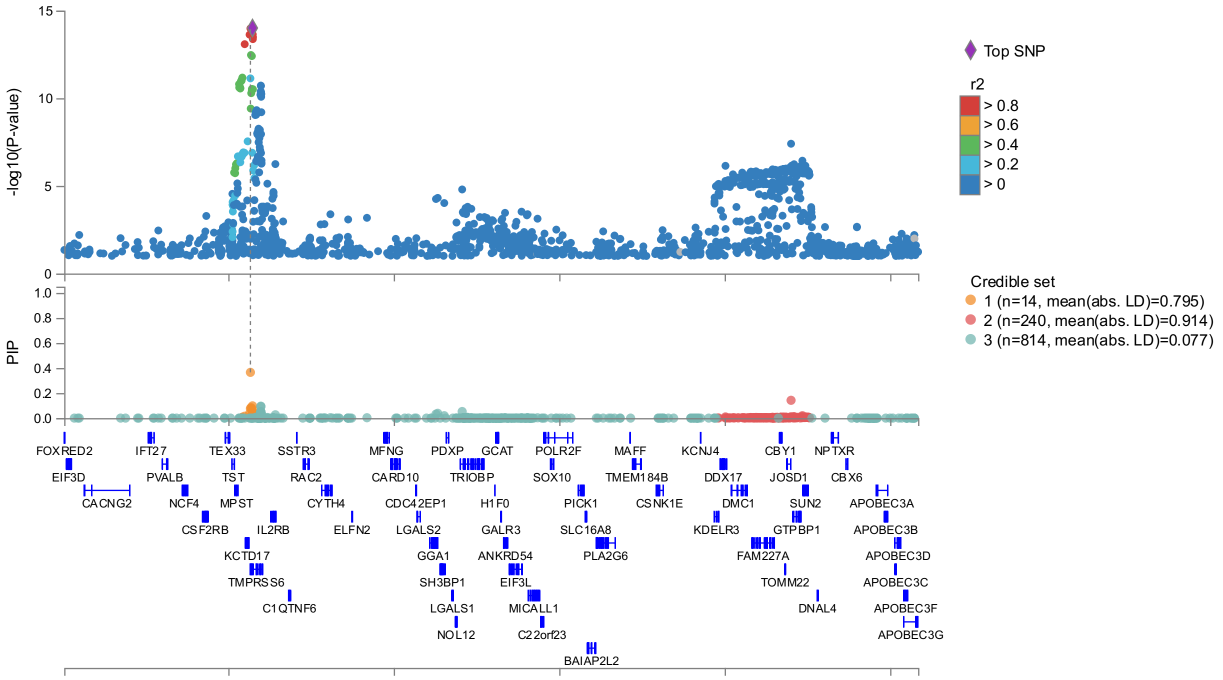

(p) AKAP6 (ECF)

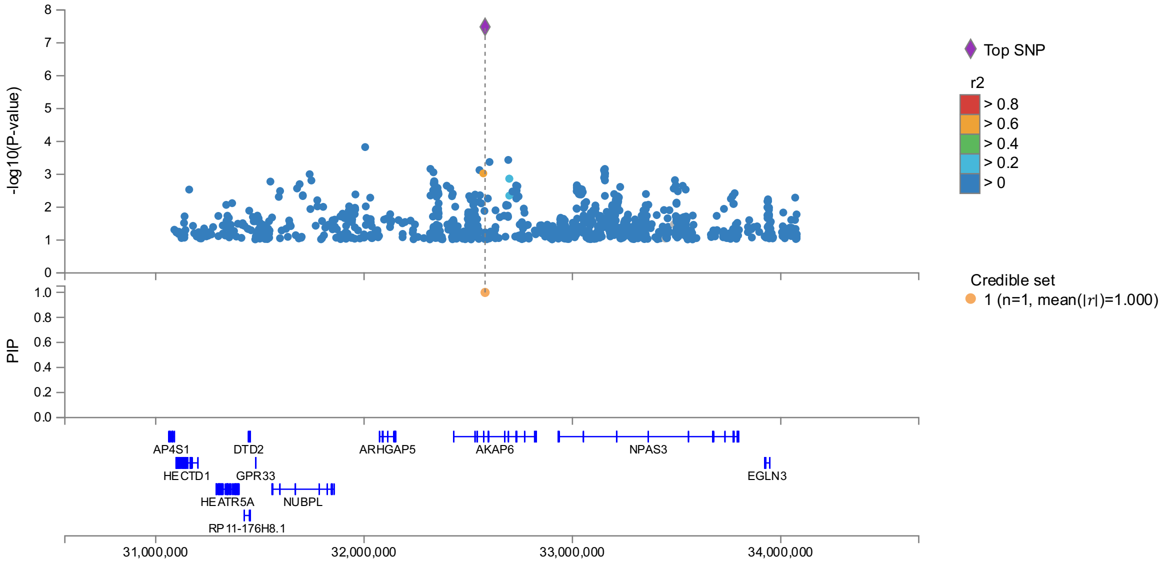

#### Supplementary Figure 6. Regional association plots for ECF.

Results shown for imputed GWAS analysis and include additional covariate adjustment for BMI and alcohol. FINEMAP estimated credible set results are shown beneath each regional association plot. (a) SLC39A8, (b), PCK2, (c) PNPLA3, (d) ABO, (e) SLC30A10, (f) TM6SF2, (g) HFE, (h) TMPRSS6, (i) FADS1, (j) FUT2, (k) NAT2, (l) AGMAT, (m) MRPL4, (n) SETBP1, (o) JOSD1, (p) AKAP6

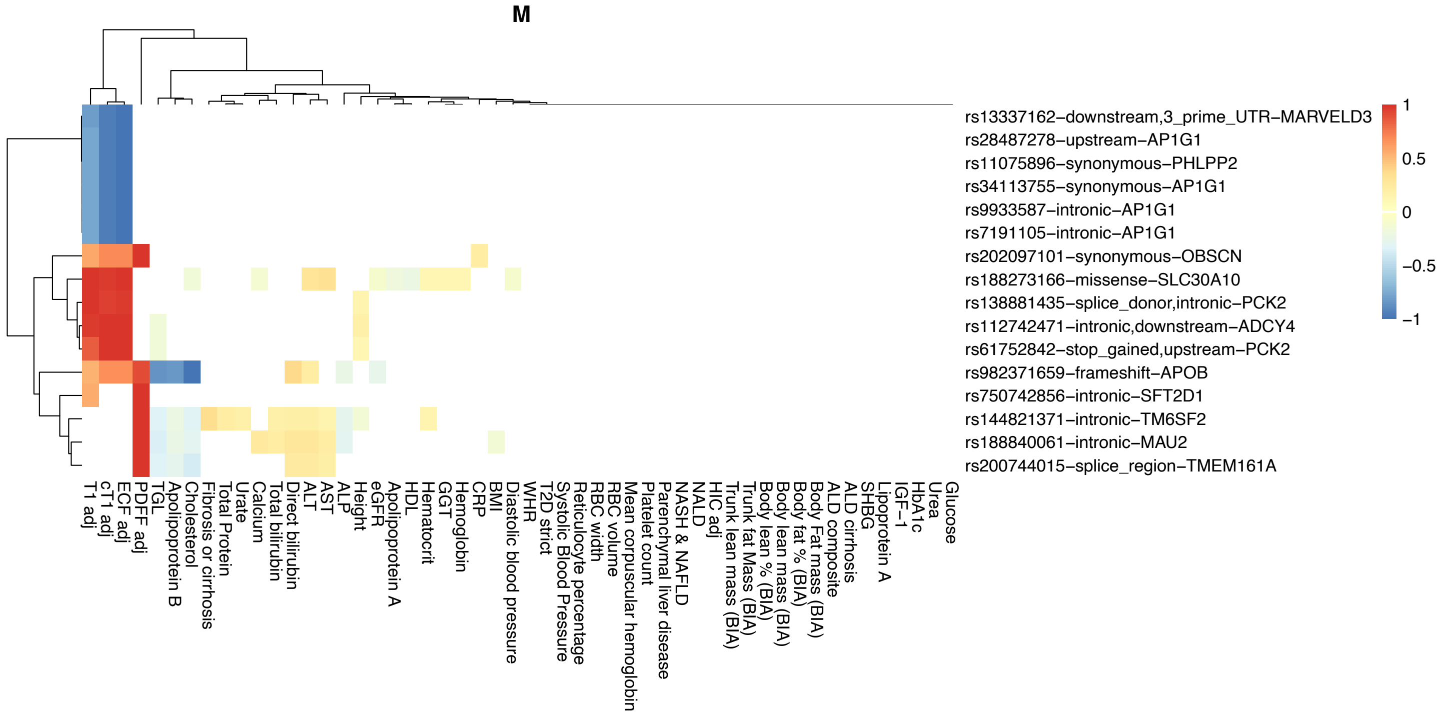

Supplementary Figure 7. Phenome-wide association results for the associated rare variants from exome data.

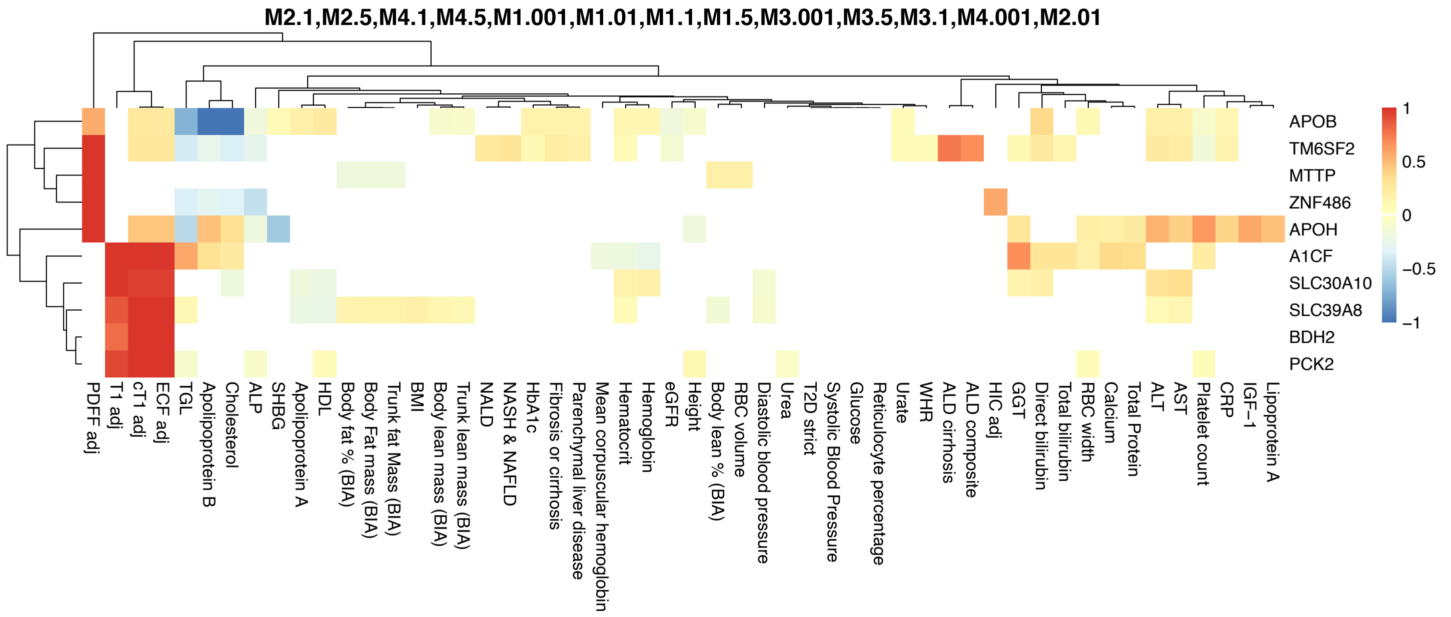

Supplementary Figure 8. Phenome-wide association results for the associated rare variant masks form exome data.

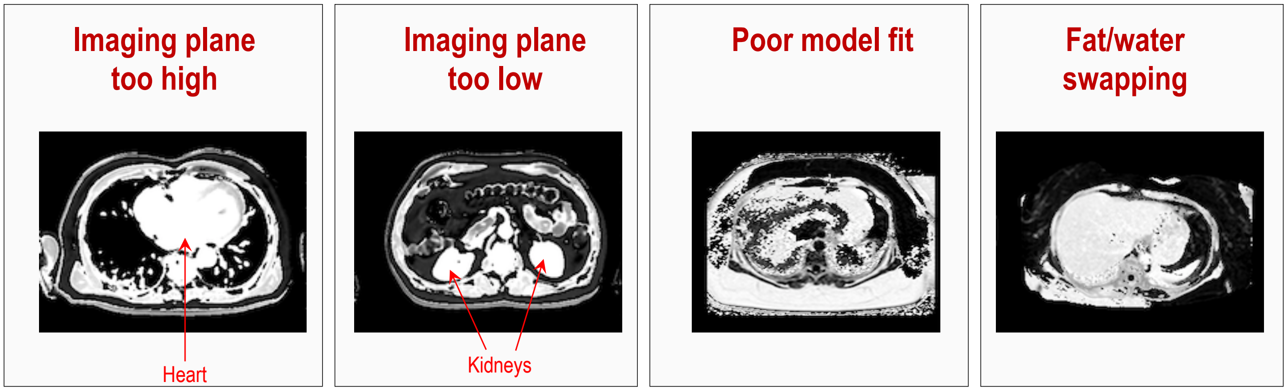

Supplementary Figure 9. Image QC examples

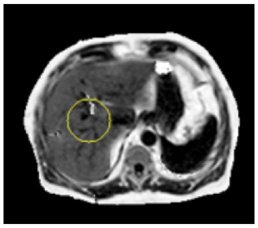

| Description | Rationale | Cut-off Threshold | % failures in test dataset |
|---|---|---|---|
| % of QC ROI overlapping with liver ROI | Mis-segmented liver overlaps less with QC ROI | 10% | 2.2% |
| Final value of cost function from PDFF parameter estimation, averaged inside QC ROI, normalized to mean magnitude | High cost function value indicates poor fit of water/fat separation model | 0.2 | 2.4% |

Supplementary Figure 10. Example of a region of interest.

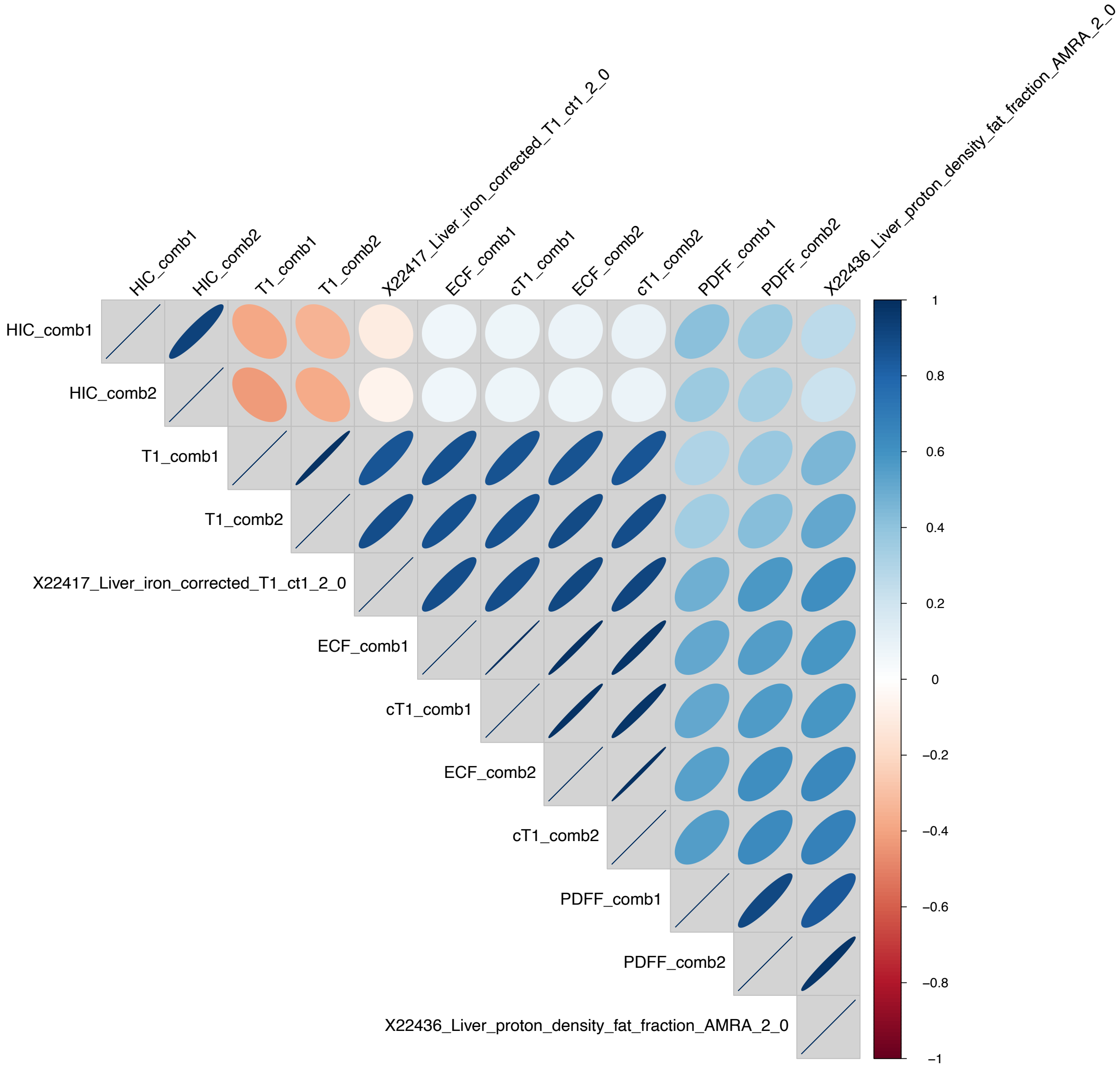

#### Supplementary Figure 11. Correlations between liver IDPs.

Correlations between all, internal and external (UKB liver iron corrected cT1, showcase id 22417 and liver PDFF (AMRA), showcase id 22436. “comb2” reflects additional deconfounding of the trait for alcohol, BMI, weight gain and various diseases, see online methods.

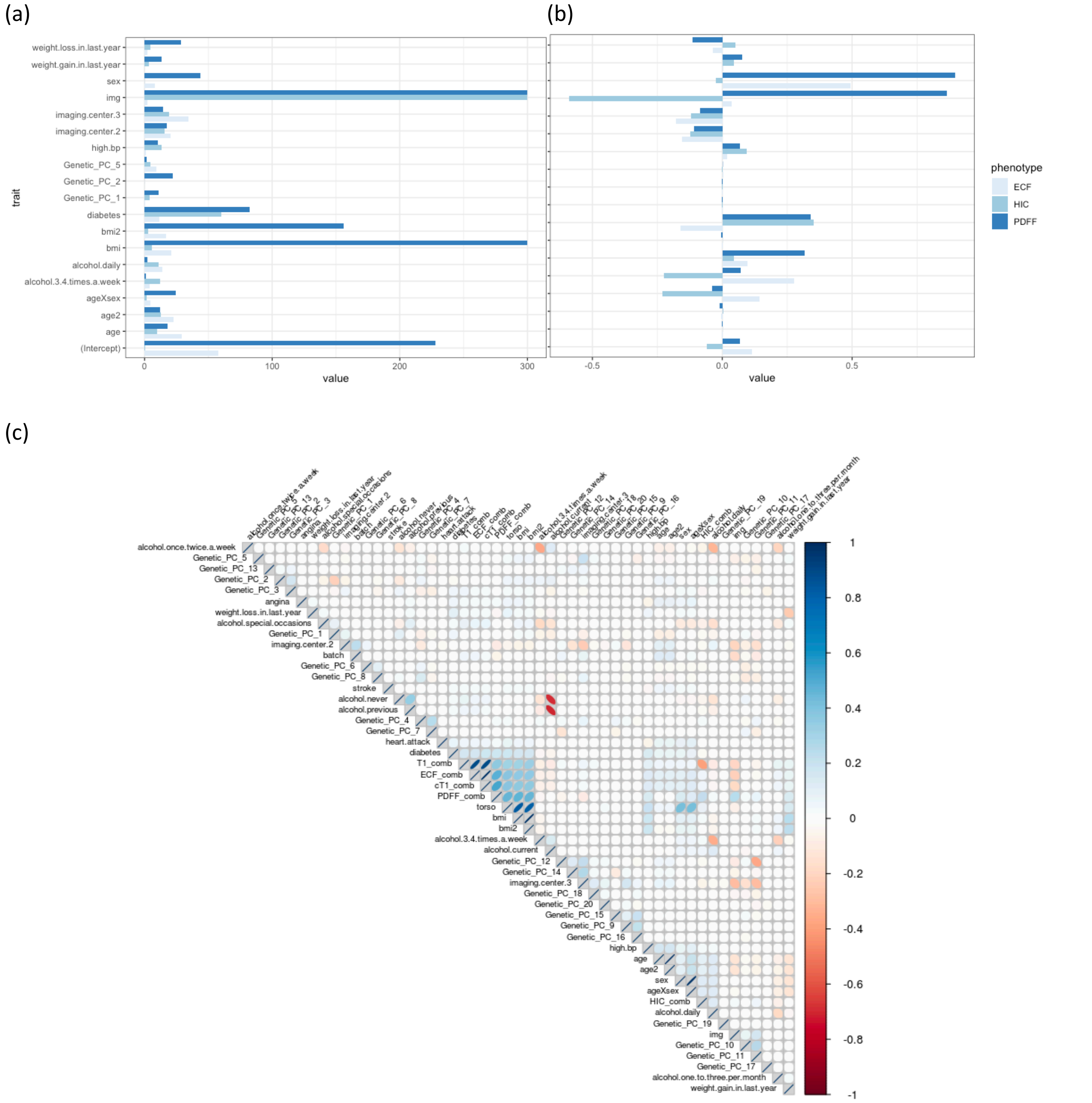

#### Supplementary Figure 12. Distribution of significance and estimates for covariate effects across ECF, HIC, PDFF.

(a) Boxplot of the distribution of -log10(P-value) for covariate effects from a linear regression of each trait against selected deconfounding variables, (b) boxplot of the distribution of estimates for covariates from the linear regression of each trait against selected deconfounding variables. For PDFF, model R-squared was 0.456, for ECF it was 0.208 and for HIC it was 0.056. (c) pairwise correlations between liver IDPs and covariates.

### Supplementary Tables

| | | | | | Haas et al. | | | | RGC | | | | | |
|---|---|---|---|---|---|---|---|---|---|---|---|---|---|---|
| Locus | Ref | Alt | Lead variant | Gene | Effect on liver fat | Effect on liver fat, P | Effect on liver fat % | Effect on liver fat %, P | Effect on liver fat (extra cov) | Effect on liver fat, P (extra cov) | Effect on liver fat | Effect on liver fat, P | Effect on liver % | Effect on liver %, P |
| 1:220796686 | A | G | rs2642438 | MARC1. | 0.05 | 2.00E-09 | 0.22 | 3.00E-09 | 0.04 | 3.93E-12 | 0.04 | 7.12E-09 | 0.21 | 6.67E-11 |
| 4:99318162:1 | T | C | rs1229984 | ADH1B | 0.16 | 7.00E-10 | 0.51 | 3.00E-06 | 0.10 | 1.06E-10 | 0.12 | 5.04E-10 | 0.50 | 1.23E-08 |
| 8:125494452 | G | A | rs112875651 | TRIB1 | -0.05 | 4.00E-10 | -0.19 | 2.00E-08 | -0.06 | 3.88E-24 | -0.05 | 1.66E-12 | -0.24 | 8.32E-16 |
| 10:11216159 | G | A | rs2250802 | GPAM | -0.05 | 1.00E-09 | -0.24 | 1.00E-10 | -0.05 | 6.25E-16 | -0.05 | 9.95E-11 | -0.27 | 5.70E-17 |
| 19:18118398 | G | T | rs56252442 | MAST3 | 0.05 | 3.00E-08 | 0.18 | 3.00E-06 | 0.03 | 1.71E-05 | 0.04 | 1.24E-07 | 0.13 | 1.89E-04 |
| Previously-identified variants | | | | | | | | | | | | | | |
| 19:19268740 | C | T | rs58542926 | TM6SF2 | 0.29 | 3.00E-85 | 1.37 | 1.00E-104 | 0.26 | 3.09E-141 | 0.26 | 4.20E-96 | 1.46 | 4.98E-148 |
| 19:44908684 | T | C | rs429358 | APOE | -0.12 | 2.00E-29 | -0.51 | 2.00E-28 | -0.08 | 4.17E-27 | -0.10 | 8.79E-30 | -0.46 | 6.97E-30 |
| 22:43928850 | C | G | rs738408 | PNPLA3 | 0.19 | 6.00E-96 | 0.88 | 1.00E-106 | 0.22 | 1.96E-253 | 0.21 | 1.41E-160 | 1.09 | 6.07E-206 |

Supplementary Table 1. Previously reported common DNA variants associated with liver fat indices. For both Haas and RGC datasets, effects are shown on transformed and original (% fat) scales.

| RSID | CPRA (GRCh38) | GENE | Effect | Pval | AAF | References |
|---|---|---|---|---|---|---|
| rs1260326 | 2:27508073:T:C | GCKR | -0.056 | 3.18E-25 | 0.606 | 1,2 |
| rs780094 | 2:27518370:T:C | GCKR | -0.053 | 4.22E-22 | 0.617 | 1,3 |
| rs2228603 | 19:19219115:C:T | NCAN | 0.193 | 5.92E-80 | 0.073 | 1,3 |
| rs12137855 | 1:219275036:C:T | LYPLAL1 | -0.008 | 0.222903 | 0.211 | 1,3 |
| rs4240624 | 8:9326721:G:A | PPP1R3B | 0.042 | 6.95E-06 | 0.910 | 1,3 |
| rs641738 | 19:54173068:T:C | MBOAT7-TMC4 | -0.033 | 8.67E-10 | 0.563 | 4,5 |
| rs62305723 | 4:87310277:G:A | HSD17B13 | 0.004 | 0.705456 | 0.066 | 6 |
| rs6834314 | 4:87292656:A:G | HSD17B13 | -0.004 | 0.486247 | 0.278 | 6,7,8 |
| rs72613567 | 4:87310240:T:TA | HSD17B13 | -0.003 | 0.639434 | 0.274 | 6,7,8 |
| rs11597086 | 10:100193948:A:C | CHUK | -0.028 | 1.83E-07 | 0.446 | 9 |
| rs10883451 | 10:100164661:T:C | ERLIN1 | -0.029 | 3.38E-08 | 0.500 | 9 |

Supplementary Table 2. Effects of variants previously associated with liver fat and NAFLD. 1=Palmer et al., 2013, 2=Parisinos et al., 2020, 3=Speliotes et al., 2011, 4=Buch et al., 2015, 5=Mancina et al., 2016, 6=Ma et al., 2019, 7=Abul-Husn et al., 2018, 8=Gellert-Kristensen et al., 2020, 9=Feitosa et al., 2013

| Variant | Gene | AF | PDFF P | PDFF Effect | ALT P | ALT Effect | AST P | AST Effect |
|---|---|---|---|---|---|---|---|---|
| 22:43928847:C:G | PNPLA3 | 0.21 | $2x10^{-127}$ | 0.22 | $6x10^{-16}$ | 0.07 | $6x10^{-18}$ | 0.07 |
| 19:19268740:C:T | TM6SF2 | 0.07 | $2x10^{-71}$ | 0.26 | $9x10^{-7}$ | 0.07 | 0.003 | 0.04 |
| 19:44908684:T:C | APOE | 0.15 | $2x10^{-24}$ | -0.11 | 0.04 | -0.02 | 0.99 | 1.00E-04 |
| 10:112150963:A:G | GPAM | 0.73 | $6x10^{-10}$ | -0.05 | $8x10^{-5}$ | -0.03 | 0.07 | -0.01 |
| 2:27508073:T:C | GCKR | 0.61 | $7x10^{-10}$ | -0.05 | 0.87 | 0.001 | 0.41 | -0.006 |
| 17:66214462:A:C | APOH | 0.03 | $1x10^{-8}$ | 0.13 | $1x10^{-6}$ | 0.1 | 0.0001 | 0.07 |
| 1:220796686:A:G | MTARC1 | 0.7 | $2x10^{-8}$ | 0.05 | 0.0008 | 0.026 | 0.08 | 0.01 |
| 22:36149089:A:T | APOL3 | 0.83 | 0.72 | 0.003 | $2x10^{-8}$ | 0.05 | 0.1 | 0.01 |

Supplementary Table 3. Top results for PDFF alongside results for ALT and AST

| | Liver Imaging | No liver imaging |
|---|---|---|
| **n** | 40,058 | 440,982 |
| **Female, n (%)** | 21,796 (54.4) | 240,382 (54.5) |
| **Age at enrollment in years, mean (SD)** | 55.6 (7.55) | 57.2 (8.14) |
| **Age at imaging in years, mean (SD)** | 64.5 (7.67) | |
| **Race, n (%)** | | |
| White | 38,765 (96.8) | 414,370 (93.9) |
| Black | 312 (0.78) | 8,260 (1.9) |
| South Asian | 614 (1.53) | 11,070 (2.51) |
| Multiple or other | 367 (0.92) | 7,282 (1.65) |
| **Had myocardial infarction, n (%)** | 887 (2.21) | 10,741 (2.44) |
| **Diabetes, n (%)** | 2,148 (5.36) | 24,811 (5.63) |
| **Obese or severely obese, n (%)** | 8,472 (21.1) | 110,406 (25.0) |
| **Hypertension, n (%)** | 6,573 (16.4) | 115,150 (26.1) |
| **Excessive alcohol intake, n (%)** | 11,178 (27.9) | 95,294 (21.6) |
| **Medications** | | |
| Anti-hypertensive, n (%) | 3,933 (9.82) | 44,391 (10.1) |
| Lipid-lowering therapy, n (%) | 3,472 (8.67) | 32,348 (7.34) |
| **Anthropometric measurements** | | |
| Body mass index, kg/m$^2$, mean (SD) | 26.3 (4.16) | 27.5 (4.83) |
| Body fat percentage, mean (SD) | 30.8 (8.08) | 31.6 (8.56) |
| **Systolic blood pressure, mean (SD)** | 140.9 (19.1) | 140.0 (19.7) |
| **Liver biomarkers** | | |
| Alanine aminotransferase (IU/L) , mean (SD) | 22.6 (13.1) | 23.6 (14.2) |
| Aspartate aminotransferase (IU/L, mean (SD) | 25.6 (9.78) | 26.3 (10.7) |
| Gamma glutamyltransferase (IU/L) , mean (SD) | 33.2 (32.6) | 37.8 (42.9) |
| **Lipid biomarkers** | | |
| Total cholesterol, mmol/L, mean (SD) | 5.73 (1.09) | 5.69 (1.15) |
| LDL cholesterol, mmol/L, mean (SD) | 3.58 (0.83) | 3.55 (0.87) |
| HDL cholesterol, mmol/L, mean (SD) | 1.48 (0.37) | 1.45 (0.38) |
| Triglycerides, mmol/L, mean (SD) | 1.63 (0.95) | 1.76 (1.03) |
| **Glycemic biomarkers** | | |
| Glycated hemoglobin HbA1c mmol/mol, mean (SD) | 35.0 (5.16) | 36.2 (6.91) |
| Glucose, mmol/L, mean (SD) | 4.99 (0.97) | 5.14 (1.27) |

Supplementary Table 4. Imaging cohort characteristics of 40,058 individuals used in this study.
Excessive alcohol is defined as any alcohol drinking below age 21 or having drinks every day or almost every day of the week.
